# Glucagon-like peptide-1 receptor agonist initiation and risk of clinically recorded Alzheimer’s disease–type dementia in older adults with type 2 diabetes: a target trial emulation using causal machine learning

**DOI:** 10.64898/2026.08.21.26361012

**Authors:** Jonas Schröder, Octavia-Andreea Ciora, Philip Heesen, Martin Bendszus, Johannes Levin, Robert Perneczky, Lia Bally, Stefan Feuerriegel

## Abstract

**Background:** Glucagon-like peptide-1 (GLP-1) receptor agonists and sodium–glucose cotransporter-2 (SGLT2) inhibitors are increasingly used for type 2 diabetes. Despite established metabolic, cardiovascular, and renal benefits, it remains uncertain whether GLP-1 receptor agonists are associated with longer clinically recorded Alzheimer’s disease (AD)-type dementia-free survival than sulfonylureas (SU) or SGLT2 inhibitors.

**Methods:** Using *All of Us* electronic health records, we emulated target trials among adults aged *≥* 55 years with type 2 diabetes, a 12-month washout, and no prior dementia. We compared GLP1 receptor agonists with SU and SGLT2 inhibitors. Propensity score weighting and doubly robust estimation addressed confounding. Causal survival forests estimated individualized treatment effects on 48-month RMST free from clinically recorded AD-type dementia.

**Findings:** In the GLP-1 receptor agonist versus SU comparison (6,328 individuals; 48-month NNT *≈* 202), initiation was associated with a small but statistically significant increase in AD-type dementia-free survival (ATE 0*·*21 months; 95% CI: 0*·*07–0*·*35). The highest-benefit stratum gained 0*·*45 months (95% CI: 0*·*28–0*·*62).

In the SGLT2 inhibitor comparison (3,070 individuals; 48-month NNT *≈* 245), the average effect was not statistically significant (ATE 0*·*06 months; 95% CI: *−*0*·*18–0*·*31), but treatment effects were heterogeneous. The highest-benefit stratum gained 0*·*83 months (95% CI: 0*·*49– 1*·*17). Predicted benefit was associated with older age, insulin use, lower HbA1c, and lower BMI.

**Interpretation:** GLP-1 receptor agonists may delay clinically recorded AD-type dementia compared with SU. Comparative effectiveness versus SGLT2 inhibitors may vary, supporting study. Given hypothesis-generating findings, diabetes treatment selection should remain guided by glycemic, cardiovascular, renal, and patient-centered considerations.

**Funding:** German Federal Ministry of Research, Technology and Space (03LWH0181B).

## Introduction

Alzheimer’s disease (AD) is the leading cause of dementia. In 2019, 57*·*4 million people lived with dementia worldwide, projected to reach 152*·*8 million by 2050.^1, 2^ Yet, prevention remains difficult because clinical AD-type dementia risk emerges from interacting genetic, metabolic, vascular, environmental, and lifestyle factors.^3, 4^ Among these risk factors, type 2 diabetes has consistently been associated with an increased risk of AD-type dementia, making glucose-lowering therapies an attractive target for dementia prevention.^1^ This multifactorial aetiology also complicates real-world evidence generation, because patients initiating glucose-lowering therapies may differ in cardiometabolic risk, diabetes treatment history, comorbidity burden, and healthcare trajectories before treatment initiation.

Glucagon-like peptide-1 (GLP-1) receptor agonists were developed for the treatment of type 2 diabetes and obesity,^5^ but their biological effects extend well beyond glycemic control.^6–8^ Experimental, mechanistic, and observational studies suggest that GLP-1 receptor agonists may modulate inflammatory, metabolic, vascular, and neuroprotective pathways implicated in clinical AD-type dementia pathophysiology,^9–12^ yet clinical evidence for long-term reduction in clinically recorded AD-type dementia risk remains inconclusive in people living with type 2 diabetes.^13–15^

In this study, we emulated two active-comparator, new-user target trials to evaluate the association between GLP-1 receptor agonist initiation and clinical AD-type dementia-free survival in individuals with type 2 diabetes (Figure 1). First, we compared GLP-1 receptor agonists with sulfonylureas (SU), to assess whether GLP-1 receptor agonist initiation was associated with longer clinical AD-type dementia-free survival. Second, we compared this association against sodium– glucose cotransporter-2 (SGLT2) inhibitors, a commonly used glucose-lowering drug class with overlapping clinical indications. For both analyses, we used causal machine learning^16, 17^ to quantify heterogeneity in treatment effects and identify clinical phenotypes associated with larger predicted benefit.

**Figure 1:**
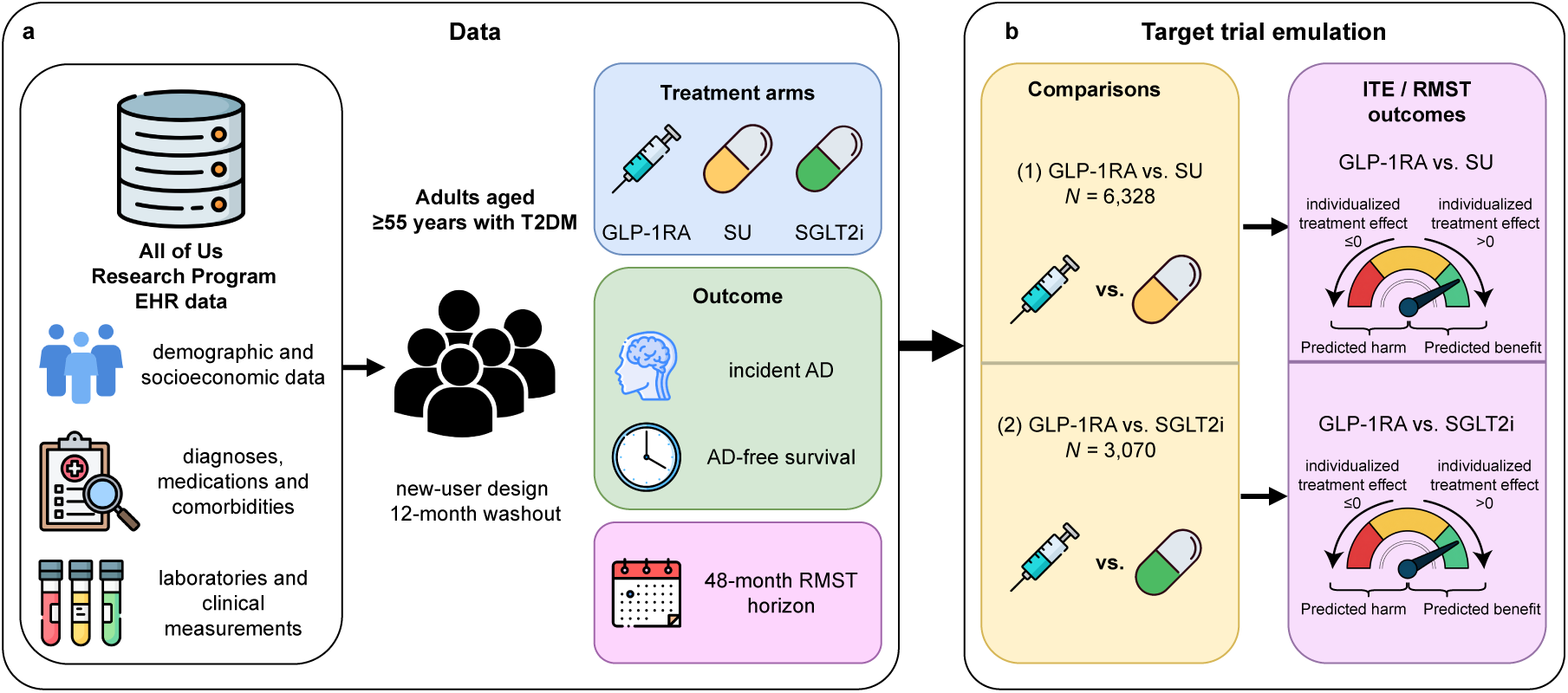
Target trial emulation using causal machine learning. (**a**) Electronic health records from the *All of Us Research Program* were used to identify adults aged *≥*55 years with type 2 diabetes using an active-comparator, new-user design with a 12-month washout period. Incident Alzheimer’s disease (AD) was evaluated over a 48-month restricted mean survival time (RMST) horizon in two emulated target trial comparisons: (1) glucagon-like peptide-1 receptor agonists (GLP-1RA) versus sulfonylureas (SU), and (2) GLP-1RA versus sodium-glucose cotransporter-2 inhibitors (SGLT2i). (**b**) For each emulated comparison, individualized treatment effects (ITEs) were estimated as differences in 48-month RMST using causal survival forests trained on demographic, socioeconomic, cardiometabolic, neurological, medication, and laboratory features. The resulting ITE distributions were used to characterize predicted benefit or harm of GLP-1RA relative to SU or SGLT2i.

## Methods

### Study design

Using the *All of Us Research Program* Controlled Tier Dataset v8 (data cutoff Oct 1, 2023),^18^ we emulated two active-comparator, new-user target trials comparing GLP-1 receptor agonists separately with SU and SGLT2 inhibitors.^19^ The first qualifying initiation defined time zero; follow-up ended at the outcome, death, loss to follow-up, the end of data availability, or the prespecified horizon. Full specifications and the temporal design are provided in Supplementary Table S1 and Supplementary Figure S15.

### Inclusion criteria

Individuals were eligible if they were aged *≥* 55 years at the index date, had a confirmed diagnosis of type 2 diabetes before baseline, and satisfied the active-comparator, new-user requirements. Eligibility was reassessed monthly using information available at time zero and a fixed 12-month look-back period, with re-entry permitted after a treatment-free washout when all criteria were again met. Full definitions, coding rules, and treatment-specific requirements are provided in the Appendix: Detailed eligibility criteria.

### Exclusion criteria

Individuals were excluded if they had prior exposure to GLP-1 receptor agonists, SU, DPP-4 inhibitors, or SGLT2 inhibitors during the 12-month washout period, or evidence of pre-existing clinical AD-type dementia or dementia. The primary analysis additionally excluded specified cardiovascular and cerebrovascular conditions recorded during the same look-back period; this restriction was removed in a complementary sensitivity analysis. Complete diagnosis codes, time windows, and reassessment rules are provided in the Appendix: Detailed eligibility criteria.

### Outcome definition

The primary outcome was incident clinically recorded AD-type dementia, defined as the first qualifying electronic health record diagnosis or anti-dementia medication dispensing after the index date; all participants were outcome-free during the 12-month baseline. This routine-care endpoint did not represent biomarker-confirmed AD. Full definitions are provided in the Appendix: Detailed outcome and covariate definitions.

### Covariates

Prespecified baseline covariates, selected based on clinical relevance and prior literature,^20^ were measured during the 12-month look-back period and covered demographic, socioeconomic, cardiometabolic, laboratory, diabetes-related, neurological, and medication factors. Full definitions and coding procedures are provided in Table S2 and the Appendix: Detailed outcome and covariate definitions.

### Causal machine learning analysis

The primary estimand was the 48-month difference in restricted mean clinical AD-type dementia-free survival, expressed as GLP-1 receptor agonist minus comparator; positive values favored GLP-1 receptor agonists. Individualized treatment effects for right-censored outcomes were estimated with causal survival forests.^21^ Propensity scores from generalized random forests^22^ were obtained with stratified five-fold cross-fitting,^23^ truncated to [0*·*05, 0*·*95], and used in doubly robust augmented inverse probability weighting (AIPW). Overlap weighting assessed balance, while IPTW-weighted Kaplan–Meier and Cox analyses served as robustness checks. Further details are provided in the Appendix: Detailed causal machine learning analysis.

### Treatment effect heterogeneity

Predicted individualized treatment effects served as effect scores, with higher values indicating greater predicted benefit from GLP-1 receptor agonists.^24^ Participants were ranked into four equally sized predicted-benefit strata, and AIPW estimated effects within each stratum. Prespecified subgroup analyses considered age, sex, BMI, body weight, obesity, HbA1c, hypertension, metformin use, and insulin use; definitions are provided in Supplementary Table S5 and the Appendix: Detailed causal machine learning analysis.

### Model validation

Following recommendations and prior applications,^16, 24, 25^ validation included covariate balance, propensity-score overlap and calibration, extreme weights, effective sample size, and an alternative Cox-based T-learner.^26^ A negative-control analysis^27^ used acute sinusitis defined from clinical guidance.^28^ Details are provided in the Appendix: Validation details.

### Implementation details

Analyses used `R` version 4.5.0, with `grf` version 2.5.0 for causal survival forests, `data.table` version 1.18.2.1 and `dplyr` version 1.2.1 for preprocessing, `arrow` version 23.0.0 for data storage and input/output, and `pROC` version 1.19.0.1 for model evaluation.

## Results

The overall eligible cohort comprised 8,812 initiators. The two primary comparisons included 7,294 individuals: 2,104 GLP-1 receptor agonist initiators, 4,224 SU initiators, and 966 SGLT2 inhibitor initiators. The remaining 1,518 DPP-4 inhibitor initiators contributed only to exploratory supplementary analyses. Pairwise cohorts included 6,328 individuals for GLP-1 receptor agonists versus SU and 3,070 for GLP-1 receptor agonists versus SGLT2 inhibitors (Figure 2).

**Figure 2:**
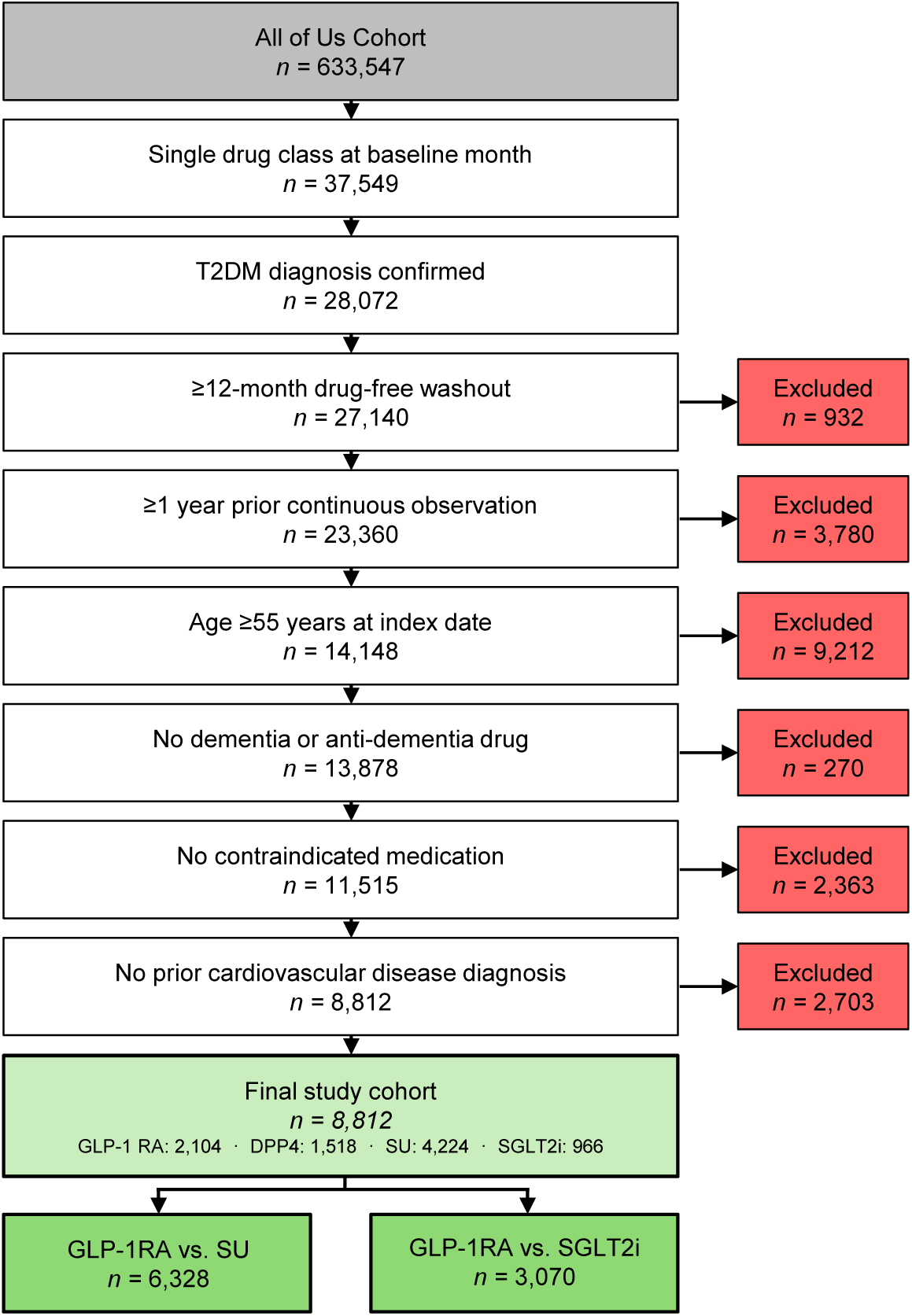
Flow chart of the inclusion criteria. Cohort construction and eligibility criteria. The overall eligible cohort included DPP-4 inhibitor initiators used only in exploratory supplementary analyses; the two primary active-comparator analyses were GLP-1 receptor agonists versus SU and versus SGLT2 inhibitors.

Selected major baseline characteristics of the treatment groups are summarized in Table 1; complete distributions and balance diagnostics for all prespecified baseline covariates are provided in Supplementary Table S3. Overall, participants were predominantly older adults with type 2 diabetes and a high burden of cardiometabolic comorbidity. GLP-1 receptor agonist initiators were more often female (67*·*9%) than SU initiators (47*·*5%) or SGLT2 inhibitor initiators (50*·*5%). Compared with SU initiators, they had higher body mass index, were more often obese or hypertensive, had lower HbA1c, were more likely to have used insulin, and were less likely to have used metformin or statins. SGLT2 inhibitor initiators had similar prevalences of hypertension and dyslipidemia but lower body mass index and obesity prevalence than GLP-1 receptor agonist initiators.

**Table 1:** Selected major baseline characteristics of patients initiating GLP-1RAs, SU, or SGLT2i. Complete baseline characteristics and pairwise standardized mean differences before and after overlap weighting are reported in Supplementary Table S3.

| Characteristic | GLP-1RA<br>(n=2104) | SU<br>(n=4224) | SGLT2i<br>(n=966) |
| --- | --- | --- | --- |
| *Demographic and socioeconomic characteristics* |  |  |  |
| Age at baseline, mean (SD), years | 63.0 (6.0) | 63.2 (6.3) | 64.9 (6.8) |
| Sex at birth, female | 1429 (67.9%) | 2005 (47.5%) | 488 (50.5%) |
| Sex at birth, male | 650 (30.9%) | 2141 (50.7%) | 470 (48.7%) |
| Sex at birth, unknown | 25 (1.2%) | 78 (1.8%) | 8 (0.8%) |
| Race, American Indian or Alaska Native | 16 (0.8%) | 39 (0.9%) | 12 (1.2%) |
| Race, Asian | 31 (1.5%) | 50 (1.2%) | 19 (2.0%) |
| Race, Black or African American | 446 (21.2%) | 865 (20.5%) | 156 (16.1%) |
| Race, more than one population | 91 (4.3%) | 169 (4.0%) | 35 (3.6%) |
| Race, none of these | 26 (1.2%) | 53 (1.3%) | 14 (1.4%) |
| Race, unknown | 349 (16.6%) | 723 (17.1%) | 187 (19.4%) |
| Race, White | 1145 (54.4%) | 2325 (55.0%) | 543 (56.2%) |
| Ethnicity, Hispanic or Latino | 344 (16.3%) | 665 (15.7%) | 173 (17.9%) |
| Ethnicity, not Hispanic or Latino | 1695 (80.6%) | 3409 (80.7%) | 752 (77.8%) |
| Ethnicity, none of these | 26 (1.2%) | 53 (1.3%) | 14 (1.4%) |
| Ethnicity, unknown | 39 (1.9%) | 97 (2.3%) | 27 (2.8%) |
| Education, college graduate or higher | 424 (20.2%) | 136 (3.2%) | 206 (21.3%) |
| Education, some college | 346 (16.4%) | 140 (3.3%) | 152 (15.7%) |
| Education, high school or less | 314 (14.9%) | 205 (4.9%) | 162 (16.8%) |
| Education, unknown | 1020 (48.5%) | 3743 (88.6%) | 446 (46.2%) |
| *Cardiometabolic and laboratory characteristics* |  |  |  |
| BMI, mean (SD), kg/m<sup>2</sup> | 37.2 (7.7) | 33.6 (7.1) | 33.1 (6.6) |
| Obesity | 885 (42.1%) | 433 (10.3%) | 221 (22.9%) |
| Hypertension | 1151 (54.7%) | 1178 (27.9%) | 550 (56.9%) |
| Dyslipidemia | 1043 (49.6%) | 1010 (23.9%) | 511 (52.9%) |
| *Continued on next page* |  |  |  |

| *Table 1 continued from previous page* |  |  |  |
| --- | --- | --- | --- |
| **Characteristic** | **GLP-1RA**<br>(*n*=2104) | **SU**<br>(*n*=4224) | **SGLT2i**<br>(*n*=966) |
| Smoking-related conditions | 106 (5.0%) | 151 (3.6%) | 48 (5.0%) |
| Alcohol-related conditions | 32 (1.5%) | 39 (0.9%) | 15 (1.6%) |
| HbA1c, mean (SD), % | 7.6 (1.8) | 8.1 (1.8) | 7.9 (1.6) |
| eGFR, mean (SD), mL/min/1.73m<sup>2</sup> | 72.1 (20.2) | 73.0 (22.1) | 73.7 (19.2) |
| *Diabetes-related and neuropsychiatric characteristics* |  |  |  |
| Diabetes severity score $\geq 1$ | 778 (37.0%) | 810 (19.2%) | 400 (41.4%) |
| Hypoglycemia history | 636 (30.2%) | 1108 (26.2%) | 236 (24.4%) |
| Psychiatric disorders | 676 (32.1%) | 591 (14.0%) | 252 (26.1%) |
| Sleep disorders | 589 (28.0%) | 355 (8.4%) | 209 (21.6%) |
| Migraine | 99 (4.7%) | 48 (1.1%) | 24 (2.5%) |
| Hearing loss | 111 (5.3%) | 89 (2.1%) | 54 (5.6%) |
| Cancer excluding non-melanoma skin cancer | 141 (6.7%) | 185 (4.4%) | 88 (9.1%) |
| *Selected prior medication use* |  |  |  |
| Antihypertensive agents (C02) | 375 (17.8%) | 883 (20.9%) | 130 (13.5%) |
| RAS agents (C09) | 1036 (49.2%) | 2501 (59.2%) | 481 (49.8%) |
| Beta-blocking agents (C07) | 496 (23.6%) | 1468 (34.8%) | 234 (24.2%) |
| Calcium channel blockers (C08) | 557 (26.5%) | 1301 (30.8%) | 243 (25.2%) |
| Diuretics (C03) | 698 (33.2%) | 1603 (37.9%) | 265 (27.4%) |
| Antiplatelet agents | 89 (4.2%) | 401 (9.5%) | 32 (3.3%) |
| Anticoagulants | 209 (9.9%) | 537 (12.7%) | 84 (8.7%) |
| Statins | 1106 (52.6%) | 2445 (57.9%) | 576 (59.6%) |
| Other lipid-lowering agents | 407 (19.3%) | 1054 (25.0%) | 149 (15.4%) |
| Antidepressants | 698 (33.2%) | 1087 (25.7%) | 262 (27.1%) |
| Metformin | 1060 (50.4%) | 2682 (63.5%) | 535 (55.4%) |
| Insulin | 688 (32.7%) | 863 (20.4%) | 241 (24.9%) |
| Oral antidiabetic combinations | 207 (9.8%) | 890 (21.1%) | 128 (13.3%) |

Values are *n* (%) unless otherwise indicated. College graduate or higher combines the “college graduate” and “advanced degree” categories; high school or less combines the “twelve or GED”, “nine through eleven”, “five through eight”, “one through four”, and “never attended” categories; diabetes severity score $\geq 1$ combines scores 1–4. BMI, body mass index; eGFR, estimated glomerular filtration rate; GLP-1RA, glucagon-like peptide-1 receptor agonist; HbA1c, glycated hemoglobin; RAS, renin–angiotensin system; SD, standard deviation; SGLT2i, sodium–glucose cotransporter-2 inhibitor; SU, sulfonylurea.

Initiation of GLP-1 receptor agonists compared with SU was associated with longer clinical AD-type dementia-free survival over 48 months, ATE 0*·*21 months (95% CI: 0*·*07–0*·*35), indicating a statistically significant population-level benefit. The distribution of individualized treatment effects shows that, even though the average benefit was small at the population level, the predicted treatment effects varied across individuals (Figure 3).

**Figure 3:**
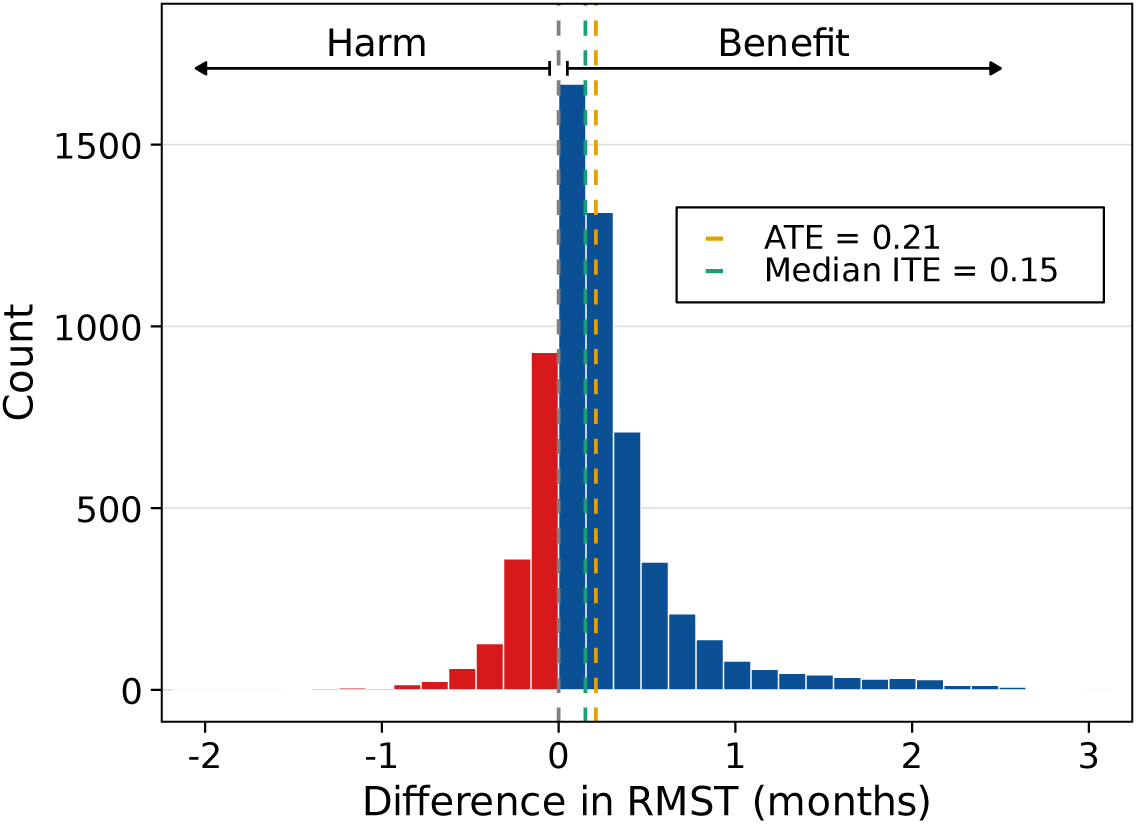
Distribution of individualized treatment effects for GLP-1 receptor agonists versus SU. Individualized treatment effects (ITEs) are expressed as differences in 48-month restricted mean survival time (RMST), in months, comparing initiation of GLP-1 receptor agonists with initiation of SU. Positive values indicate longer predicted clinical AD-type dementia-free survival under GLP-1 receptor agonists, whereas negative values indicate longer predicted clinical ADtype dementia-free survival under SU. The dashed vertical line indicates the average treatment effect (ATE), and the solid vertical line indicates the median ITE.

To further characterize heterogeneity, individuals were ranked according to the individualized treatment effects predicted by the final causal survival forest and divided into four equally sized strata. Within each stratum, the ATE was estimated via AIPW (Supplementary Figure S1b). Estimated clinical AD-type dementia-free survival benefits were 0*·*04 months in Q1 (95% CI: *−*0*·*15– 0*·*22), 0*·*22 months in Q2 (95% CI: 0*·*13–0*·*32), 0*·*13 months in Q3 (95% CI: 0*·*01–0*·*24), and 0*·*45 months in Q4 (95% CI: 0*·*28–0*·*62).

We repeated the stratum-specific analysis using a 60-month RMST horizon to assess whether the predicted benefit ranking remained informative. The measured clinical AD-type dementia-free survival differences were 0*·*14 months in Q1 (95% CI: *−*0*·*08 to 0*·*36), *−*0*·*09 months in Q2 (95% CI: *−*0*·*26 to 0*·*08), 0*·*35 months in Q3 (95% CI: 0*·*17 to 0*·*52), and 0*·*51 months in Q4 (95% CI: 0*·*28 to 0*·*75; Supplementary Figure S14b).

Subgroup analyses showed positive RMST differences across various clinically defined strata, although the magnitude of the estimated benefit varied substantially (Figure 4). Statistically significant positive effects were observed in participants aged <65 years (0*·*17 months, 95% CI: 0*·*04–0*·*30), women (0*·*24 months, 95% CI: 0*·*04–0*·*45), participants without obesity (0*·*28 months, 95% CI: 0*·*11–0*·*44), participants without hypertension (0*·*24 months, 95% CI: 0*·*10–0*·*38), and participants not receiving insulin (0*·*28 months, 95% CI: 0*·*05–0*·*50). The estimated effect sizes were larger in smaller subgroups, including participants aged *≥* 80 years (2*·*61 months, 95% CI: *−*0*·*03–5*·*25), participants with BMI <25 kg/m^2^ (2*·*36 months, 95% CI: 0*·*15–4*·*58), and participants with lower body weight (0*·*50 months, 95% CI: 0*·*11–0*·*89). Treatment effects differed across cardiometabolic strata. Participants without obesity showed a positive RMST difference (0*·*27 months, 95% CI: 0*·*11–0*·*44), whereas the estimate among participants with obesity was close to zero (*−*0*·*05 months, 95% CI: *−*0*·*34–0*·*25). Similarly, participants without hypertension showed a positive RMST difference (0*·*24 months, 95% CI: 0*·*10–0*·*38). Variation was also observed by insulin use and baseline glycemic control. Participants not receiving insulin showed a positive RMST difference (0*·*28 months, 95% CI: 0*·*05–0*·*50). Across HbA1c strata, the largest estimate was observed among participants with HbA1c 7–8% (0*·*33 months, 95% CI: 0*·*03–0*·*63), while estimates were smaller in participants with HbA1c <7% (0*·*07 months, 95% CI: *−*0*·*09–0*·*24) and HbA1c *≥* 9% (0*·*09 months, 95% CI: *−*0*·*40–0*·*57). No subgroup had a 95% CI entirely below zero, indicating that none of the defined subgroups consistently favored SU over GLP-1 receptor agonists.

GLP-1 receptor agonist initiation showed an ATE of 0*·*06 months (95% CI: *−*0*·*18–0*·*31) compared with SGLT2 inhibitors. The distribution of individualized treatment effects was centered around zero, but predicted RMST differences spanned both negative and positive values, suggesting bidirectional heterogeneity rather than a consistent advantage of either drug class across the cohort (Figure 5).

**Figure 4:**
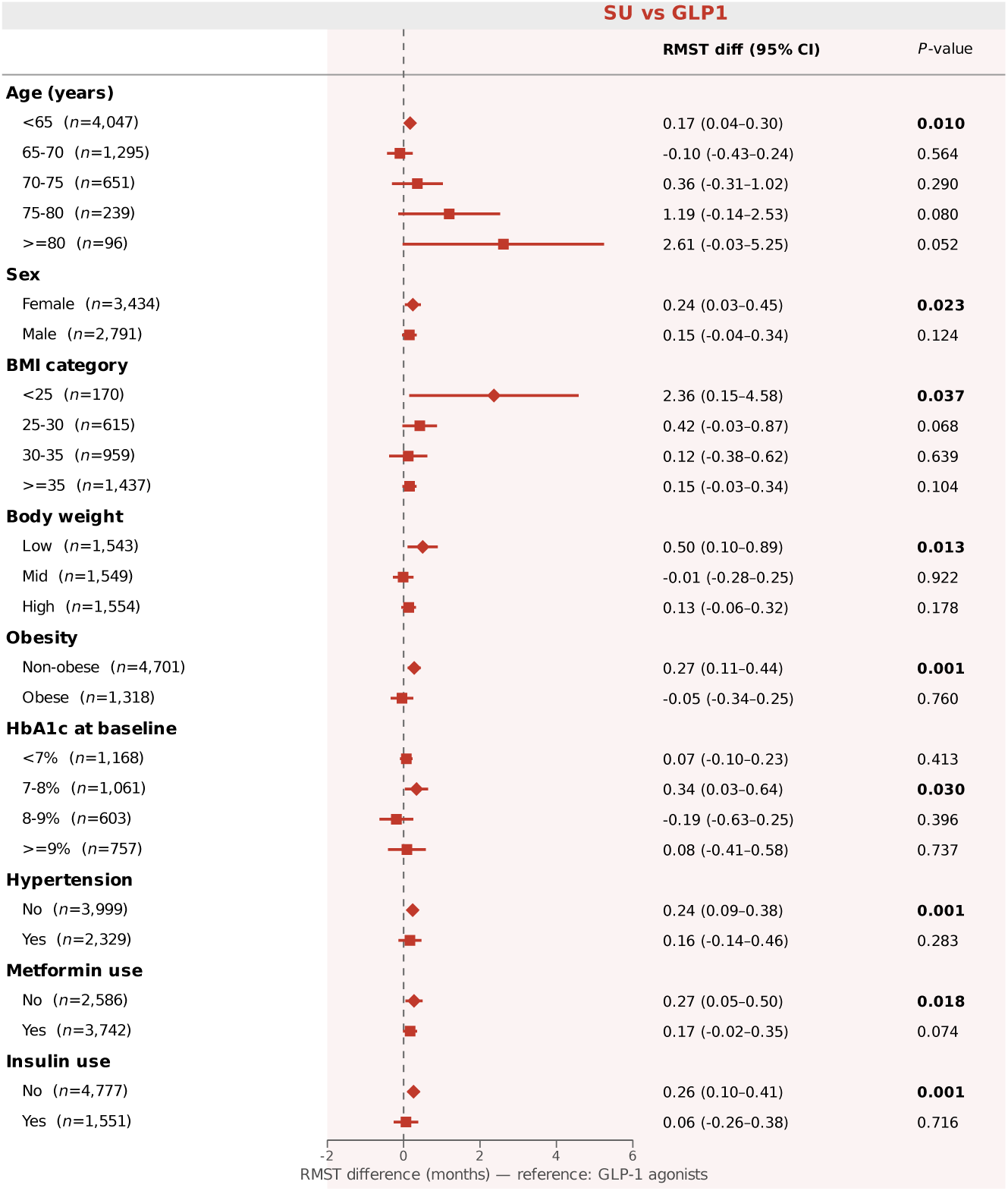
Comparison of treatment effects across patient subgroups for GLP-1 versus SU. Subgroup-specific treatment effects are shown as differences in 48-month restricted mean survival time (RMST), in months. Estimates are reported with 95% confidence intervals, shown by horizontal whiskers. Positive values indicate longer clinical AD-type dementia-free survival under GLP-1 receptor agonist initiation, whereas negative values indicate longer clinical AD-type dementia-free survival under SU initiation. The vertical dashed line indicates no difference between treatment groups. Subgroup-specific sample sizes may not sum to the full analytic cohort because individuals with missing values for the subgroup-defining variable were excluded from that specific subgroup analysis. *P* -values correspond to subgroup-specific treatment effect estimates. Abbreviations: AD, Alzheimer’s disease; BMI, body mass index; CI, confidence interval; GLP-1, glucagon-like peptide-1; HbA1c, glycated hemoglobin; RMST, restricted mean survival time; SU, sulfonylurea.

**Figure 5:**
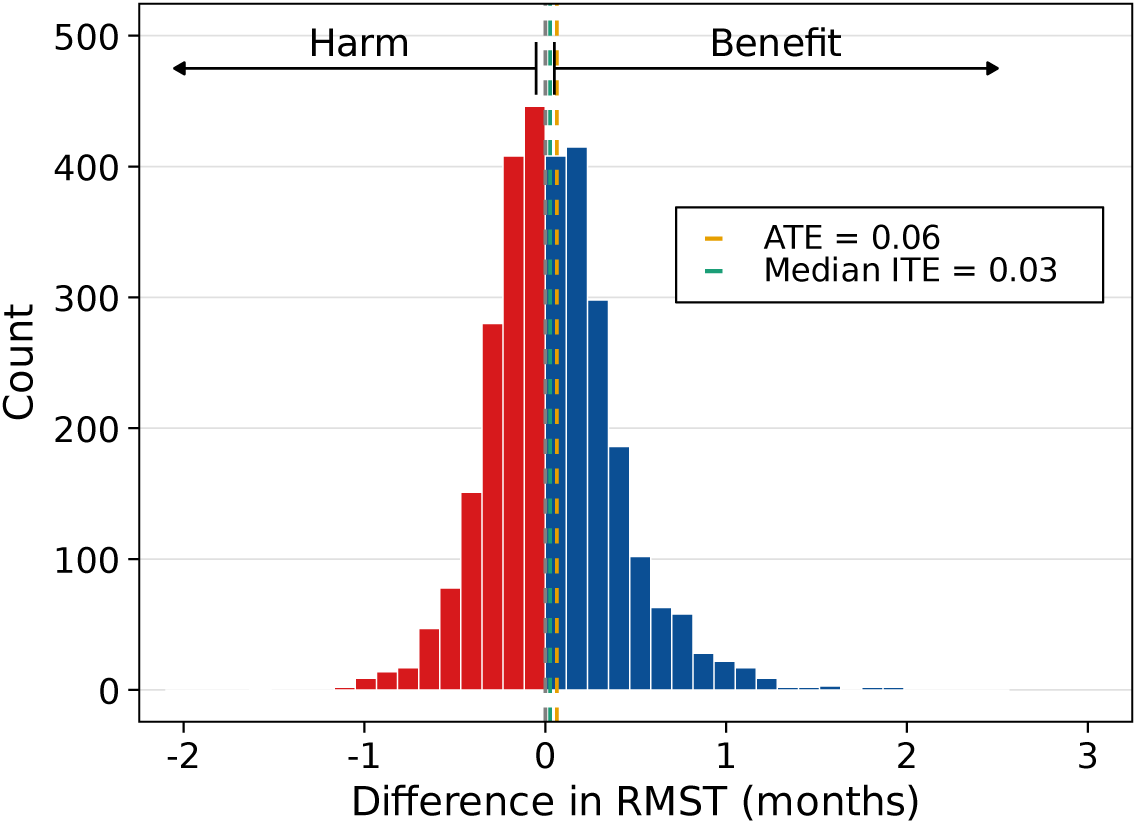
Distribution of individualized treatment effects for GLP-1 receptor agonists versus SGLT2 inhibitors. Individualized treatment effects (ITEs) are expressed as differences in 48month restricted mean survival time (RMST), in months, comparing initiation of GLP-1 receptor agonists with initiation of SGLT2 inhibitors. Positive values indicate longer predicted clinical AD-type dementia-free survival under GLP-1 receptor agonists, whereas negative values indicate longer predicted clinical AD-type dementia-free survival under SGLT2 inhibitors. The dashed vertical line indicates the average treatment effect (ATE), and the solid vertical line indicates the median ITE.

The effect score analysis compared clinical AD-type dementia-free survival differences across quartiles of predicted individual benefit from GLP-1 receptor agonist initiation (see Supplementary Figure S1a). In the GLP-1 receptor agonist versus SGLT2 inhibitor comparison, estimates increased across quartiles, from *−*0*·*25 months in Q1 (95% CI: *−*0*·*52 to 0*·*03) and *−*0*·*03 months in Q2 (95% CI: *−*0*·*18 to 0*·*12) to 0*·*17 months in Q3 (95% CI: 0*·*01 to 0*·*34) and 0*·*83 months in Q4 (95% CI: 0*·*49 to 1*·*17).

Beyond the primary 48-month endpoint, we also examined whether patterns of predicted treatment effect heterogeneity persisted over a longer follow-up horizon (see Figure S14a). In the GLP1 receptor agonist versus SGLT2 inhibitor comparison, the extended 60-month analysis yielded *−*0*·*46 months in Q1 (95% CI: *−*0*·*88 to *−*0*·*04), *−*0*·*41 months in Q2 (95% CI: *−*0*·*72 to *−*0*·*10), 0*·*25 months in Q3 (95% CI: 0*·*03 to 0*·*47), and 0*·*91 months in Q4 (95% CI: 0*·*46 to 1*·*36).

Subgroup analyses demonstrated variation in RMST differences across clinically defined strata (Figure 6). A negative RMST difference was observed in participants aged 65–70 years (*−*0*·*57 months, 95% CI: *−*1*·*08–*−*0*·*07), whereas estimates in older age groups were positive, including participants aged *≥* 80 years (2*·*91 months, 95% CI: *−*2*·*80–8*·*60). Across BMI categories, participants with BMI <25 kg/m^2^ showed a negative RMST difference (*−*0*·*15 months, 95% CI: *−*0*·*28–*−*0*·*02), while estimates in higher BMI categories were variable. Stratification by cardiometabolic factors showed no consistent directional pattern. Estimates were 0*·*13 months (95% CI: *−*0*·*21– 0*·*47) among participants without obesity and *−*0*·*06 months (95% CI: *−*0*·*48–0*·*36) among participants with obesity. Similarly, estimates were 0*·*12 months (95% CI: *−*0*·*19–0*·*42) in participants without hypertension and 0*·*02 months (95% CI: *−*0*·*35–0*·*39) in participants with hypertension. Treatment effect estimates also varied by baseline glycemic control and insulin use. Across HbA1c, estimates included 0*·*47 months (95% CI: *−*0*·*07–1*·*02) for HbA1c <7%, *−*0*·*04 months (95% CI: *−*0*·*11–0*·*03) for HbA1c 7–8%, and near-zero or negative estimates in higher HbA1c categories. The estimated RMST difference was 0*·*32 months (95% CI: *−*0*·*34–0*·*98) among insulin users and *−*0*·*05 months (95% CI: *−*0*·*25–0*·*16) among non-insulin users.

**Figure 6:**
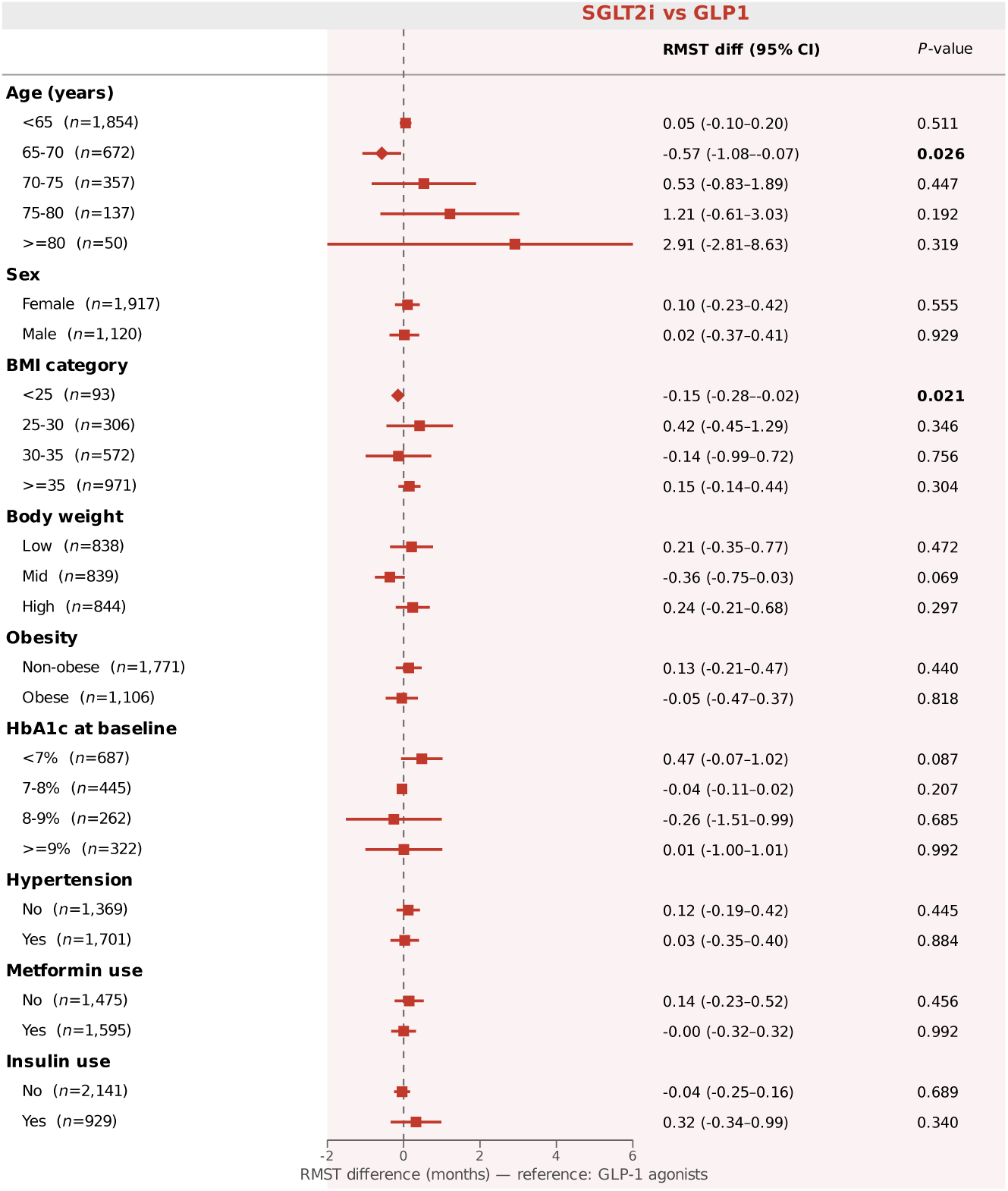
Comparison of treatment effects across patient subgroups for GLP-1 versus SGLT2 inhibitors. Subgroup-specific treatment effects are shown as differences in 48-month restricted mean survival time (RMST), in months. Estimates are reported with 95% confidence intervals, shown by horizontal whiskers. Positive values indicate longer clinical AD-type dementia-free survival under GLP-1 receptor agonist initiation, whereas negative values indicate shorter clinical AD-type dementia-free survival under SGLT2 inhibitors initiation. The vertical dashed line indicates no difference between treatment groups. Subgroup-specific sample sizes may not sum to the full analytic cohort because individuals with missing values for the subgroup-defining variable were excluded from that specific subgroup analysis. *P* -values correspond to subgroup-specific treatment effect estimates. Abbreviations: AD, Alzheimer’s disease; BMI, body mass index; CI, confidence interval; GLP-1, glucagon-like peptide-1; HbA1c, glycated hemoglobin; RMST, restricted mean survival time; SGLT2 inhibitor, sodium–glucose cotransporter-2 inhibitor.

To analyze whether the primary dementia-related predicted-benefit strata were accompanied by differences in cardiometabolic follow-up measures, we summarized secondary outcomes across the same primary individualized treatment effect quartiles using IPW. These analyses were descriptive; quartiles were defined using the predicted individualized treatment effects for dementia-free survival from the primary model, and secondary outcomes were not used to define the strata. IPW-adjusted means for changes in BMI, body weight, and HbA1c are shown in Table S4.

In the SU comparison, GLP-1 receptor agonist initiators showed larger weight reductions across all four primary predicted-benefit quartiles, with IPW-adjusted differences ranging from *−*2*·*28 kg in Q1 to *−*2*·*81 kg in Q2. BMI differences were smaller in Q1 but larger in Q2–Q4, ranging from *−*0*·*89 to *−*1*·*20 BMI units in these strata. In contrast, HbA1c differences did not show a consistent pattern across quartiles; the IPW-adjusted differences ranged from *−*0*·*26 percentage points in Q1 to 0*·*01 percentage points in Q2, and all 95% CIs included zero.

In the SGLT2 inhibitor comparison, secondary cardiometabolic profiles were less clearly separated across the primary predicted-benefit quartiles. For BMI, body weight, and HbA1c, the IPW-adjusted differences were smaller than in the SU comparison, and all 95% CIs included zero across Q1–Q4. BMI differences ranged from *−*0*·*64 to 0*·*19 BMI units, weight differences from *−*0*·*96 to 0*·*13 kg, and HbA1c differences from *−*0*·*39 to *−*0*·*02 percentage points.

The secondary outcome profiles did not indicate that the primary predicted dementia-related benefit strata were uniformly aligned with larger improvements in conventional cardiometabolic markers. In the SU comparison, GLP-1 receptor agonist initiators had consistently larger weight reductions, but HbA1c differences were variable. In the SGLT2 inhibitor comparison, cardiometabolic differences were small across strata.

Robustness checks were consistent with the primary analysis. First, in the negative-control outcome analysis using acute sinusitis, the primary analysis did not show a consistent treatment benefit associated with GLP-1 receptor agonist initiation across predicted-benefit strata (Supplementary Figure S6). Second, IPTW-weighted Kaplan–Meier curves and Cox proportional hazards models yielded marginal hazard-ratio estimates close to the null: 0*·*99 for SU versus GLP-1 receptor agonists (95% CI: 0*·*53–1*·*86; *P* = 0*·*980) and 1*·*53 for SGLT2 inhibitors versus GLP-1 receptor agonists (95% CI: 0*·*73–3*·*19; *P* = 0*·*257; Supplementary Figure S11). Third, the Tlearner results showed the same direction of ranking performance as the primary causal survival forest, with larger clinical AD-type dementia-free time lost among discordant than concordant patient pairs: 0*·*13 months in the SU comparison and 0*·*25 months in the SGLT2 inhibitor comparison (Supplementary Figure S12). Fourth, RATE curves showed larger estimated treatment effects among participants ranked highest by predicted benefit, particularly in the SGLT2 inhibitor comparison, indicating that the predicted-benefit scores enriched for individuals with larger expected time gained under GLP-1 receptor agonists (Supplementary Figure S3). Together, these analyses supported the robustness of the estimated heterogeneity patterns.

## Discussion

In this target trial emulation of older adults with type 2 diabetes, initiation of GLP-1 receptor agonists was associated with a small increase in clinically recorded AD-type dementia-free survival compared with SU over 48 months. The estimated population-level difference was 0*·*21 months (95% CI 0*·*07–0*·*35), with a larger average difference of 0*·*45 months (95% CI 0*·*28–0*·*62) in the stratum ranked as having the greatest predicted benefit. By contrast, GLP-1 receptor agonists showed no clear average advantage over SGLT2 inhibitors (0*·*06 months, 95% CI *−*0*·*18 to 0*·*31), although the model ranked groups with estimated effects ranging from benefit under SGLT2 inhibitors in the lowest quartile to benefit under GLP-1 receptor agonists in the highest quartile. The 60-month sensitivity analysis showed broadly similar ranking patterns. These results suggest a favorable but small comparative association against SU and possible treatment effect heterogeneity against SGLT2 inhibitors, but they do not establish a clinically actionable benefit for an individual patient.

Our findings should be interpreted within an evolving and partly discordant evidence base. Pooled cardiovascular outcome trials and Danish registry analyses reported lower dementia incidence among patients exposed to GLP-1 receptor agonists, although dementia was not the primary endpoint and broad dementia definitions were used.^29^ A Swedish emulated trial similarly reported lower dementia risk for GLP-1 receptor agonists than for SU and DPP-4 inhibitors in older adults with type 2 diabetes.^20^ A meta-analysis of randomized trials also suggested a class-specific reduction in broad dementia or cognitive-impairment outcomes for GLP-1 receptor agonists, but did not show a clear association for AD dementia specifically; moreover, dementia events were uncommon and generally secondary outcomes.^15^ More recently, the evoke and evoke+ phase 3 trials did not show that oral semaglutide slowed clinical progression in early symptomatic AD despite changes in some disease-related biomarkers.^14^ These trials address symptomatic treatment rather than prevention before clinically recorded dementia, but they temper any disease-modifying interpretation of our observational findings. Our favorable comparison with SU is therefore broadly concordant with evidence derived from diabetes populations and older glucose-lowering comparators. However, it should not be interpreted as evidence that GLP-1 receptor agonists prevent biomarker-confirmed AD, because our endpoint reflects clinically recorded diagnoses and anti-dementia medication use rather than adjudicated neuropathology.

Evidence comparing GLP-1 receptor agonists with SGLT2 inhibitors is less consistent. Our near-null average estimate indicates little separation between the two contemporary drug classes over 48 months. This differs from a large comparative-effectiveness study that reported modestly lower risks of some dementia outcomes with SGLT2 inhibitors than with GLP-1 receptor agonists.^30^ The apparent contrast between the favorable result against SU and the absence of a clear advantage against SGLT2 inhibitors may therefore be explained partly by comparator choice. SU is an older glucose-lowering alternative, whereas GLP-1 receptor agonists and SGLT2 inhibitors have overlapping indications and both modify metabolic and vascular pathways that may be relevant to dementia risk. Accordingly, the SU comparison addresses whether GLP-1 receptor agonists are associated with more favorable dementia-related outcomes than an older active comparator, whereas the SGLT2 inhibitor comparison asks whether one modern treatment class is preferable to another. These are distinct causal contrasts, and concordance is not necessarily expected.

The secondary cardiometabolic analyses provide insight into possible mechanisms. In the SU comparison, GLP-1 receptor agonist initiators lost more weight in every predicted dementia-benefit quartile, but weight loss did not increase with predicted dementia benefit. Adjusted differences in body weight ranged from approximately *−*2*·*3 to *−*2*·*8 kg across Q1–Q4, whereas dementia-related RMST differences increased from approximately zero in Q1 to 0*·*45 months in Q4. HbA1c changes were variable and showed no corresponding gradient. In the SGLT2 inhibitor comparison, differences in body weight, BMI, and HbA1c were small and did not systematically align with the dementia-benefit ranking. Thus, participants ranked as deriving larger dementia-related benefit were not consistently those with the greatest weight loss or glycemic improvement. This discordance argues against short-term weight loss as the primary mechanism underlying the predicted strata and makes it unlikely that weight loss alone explains the observed associations. These analyses were descriptive, restricted to participants with available follow-up measurements, and did not formally estimate mediation. They therefore cannot exclude longer-term metabolic effects or indirect pathways involving vascular risk, inflammation, physical activity, or treatment adherence, but suggest that the dementia-related ranking captures information beyond conventional changes in weight and HbA1c.

Several limitations are important. Most fundamentally, treatment was not randomized, and residual confounding may remain despite adjustment for measured baseline characteristics. Frailty and functional status were not adequately captured. Clinicians may avoid GLP-1 receptor agonists in frail patients because of gastrointestinal adverse effects, treatment burden, or concern about further weight loss, while healthier and more functionally independent patients may be more likely to receive a newer injectable therapy. Such selection could create an apparent dementia-related advantage for GLP-1 receptor agonists, particularly compared with SU, even in the absence of a direct neuroprotective effect. Conversely, contraindications and clinical indications may channel patients with different cardiometabolic risks toward SGLT2 inhibitors, making the direction of confounding in the modern-comparator analysis difficult to predict.

The marked sex imbalance between treatment groups also warrants consideration. GLP-1 receptor agonist initiators were more often female (67*·*9%) than SU initiators (47*·*5%) or SGLT2 inhibitor initiators (50*·*5%), which may reflect sex-related differences in treatment selection, obesity burden, treatment preferences, access, or other measured and unmeasured clinical and social factors. Although sex at birth was included in the propensity and outcome models and post-weighting balance was assessed, residual confounding or effect modification related to sex cannot be excluded.

The cardiovascular-disease exclusion may also have selected a healthier cohort and removed patients with high baseline risk and potentially larger absolute benefit. However, the sensitivity analysis retaining cardiovascular and cerebrovascular disease yielded attenuated, statistically nonsignificant average estimates (0*·*13 months [95% CI *−*0*·*01 to 0*·*28] versus SU and 0*·*03 months [*−*0*·*21 to 0*·*27] versus SGLT2 inhibitors). Thus, this analysis did not suggest that the primary restriction concealed a larger average benefit, although the changed cohort and limited precision preclude firm conclusions.

Further clinical context, strengths, subgroup interpretation, and methodological and mechanistic limitations are provided in the Supplementary Discussion.

In summary, GLP-1 receptor agonist initiation was associated with a small increase in clinically recorded AD-type dementia-free survival compared with SU, in concordance with several observational studies using older glucose-lowering comparators. No clear average advantage was observed compared with SGLT2 inhibitors, and direct comparative evidence between these modern classes remains mixed. Weight loss and glycemic improvement did not align with the predicted dementia-benefit strata, arguing against weight loss as the primary mechanism for the observed heterogeneity. Although causal survival forests identified groups enriched for larger estimated effects, the results support group-level ranking rather than precise individual treatment effect prediction, and the inconsistent age, BMI, and HbA1c patterns should not be given a post hoc biological interpretation. Residual confounding by frailty, cognitive reserve, education, healthcare engagement, and healthy-user behavior remains plausible. The findings are therefore hypothesis-generating and should not alter current diabetes treatment selection, but they motivate longer-term validation in independent, biomarker-supported cohorts.

## Protocol availability

A retrospectively documented protocol recording the study design and analyses described in this report is provided as a separate submission file. It should not be interpreted as evidence of prospective registration.

## Data sharing

This study used deidentified demographic, electronic health record, medication, laboratory, survey, and physical-measurement data from the *All of Us Research Program* Controlled Tier Dataset v8 (CDR v8; data cutoff Oct 1, 2023), available to authorised users through the Researcher Workbench (https://workbench.researchallofus.org/). Individual participant data and the study-specific analytic dataset cannot be distributed by the authors because *All of Us* prohibits copying, downloading, or redistributing participant-level data outside the secure Researcher Workbench. Researchers wishing to undertake biomedical or health research permitted by the programme’s Data User Code of Conduct may apply independently at https://www.researchallofus.org/register/. Access is determined by *All of Us*, not by the authors, and requires an institutional Data Use and Registration Agreement for the Controlled Tier, account and identity verification, completion of Responsible Conduct of Research and applicable tier-specific training, and acceptance of the Data User Code of Conduct. Access begins when programme authorisation is granted and continues only while the researcher remains authorised and CDR v8 is retained by *All of Us*; the programme retains its three most recent curated data releases. Official data dictionaries and release information are available at https://support.researchallofus.org/hc/en-us/articles/360033200232-Data-Dictionaries. Authorised researchers using CDR v8 can reproduce the final analytic cohorts from the cohort definitions, OMOP concept mappings, preprocessing specifications, and analysis code. The retrospectively documented study protocol accompanies this submission as a separate file; the completed STROBE and RECORD checklists are appended to this compiled manuscript and can also be supplied as separate submission files. Code for cohort construction, preprocessing, causal survival forest estimation, validation analyses, and figure generation will be deposited in a permanent public repository upon publication; until a repository URL is assigned, it is available from the corresponding author on reasonable request, subject to institutional and *All of Us* disclosure and data-use restrictions.

## Data Availability

The individual-level data analysed in this study were obtained from the NIH All of Us Research Program Controlled Tier Dataset v8 (data cutoff Oct 1, 2023) and are available to authorised researchers only through the secure Researcher Workbench (https://workbench.researchallofus.org/). Access requires an institutional Data Use and Registration Agreement, researcher registration and identity verification, completion of the required training, and acceptance of the Data User Code of Conduct. The authors are not permitted to download, redistribute, or provide the participant-level data or the study-specific analytic dataset. Eligible researchers can apply for access at https://www.researchallofus.org/register/. All aggregate data supporting the findings are contained in the manuscript and its appendix.

## Acknowledgements

This work has been supported by the German Federal Ministry of Research, Technology and Space (Grant: 03LWH0181B).

## Author contributions

All authors contributed to conceptualization, manuscript writing, and approved the manuscript.

## Supplementary Materials

### Detailed study design

We emulated two target trials following established principles for target trial emulation.^1^ For each comparison, we defined the eligibility criteria, treatment strategies, treatment assignment, time zero, follow-up, outcome definition, and censoring rules before analysis. The emulated target trial design aligned the observational analysis with the protocol of a hypothetical randomized trial to reduce biases related to immortal time, prevalent use, and post-baseline treatment decisions.

Both target trials followed an active-comparator, new-user design among individuals with type 2 diabetes. In the first target trial, initiation of GLP-1 receptor agonists was compared with initiation of SU; in the second target trial, initiation of GLP-1 receptor agonists was compared with initiation of SGLT2 inhibitors. Unless stated otherwise, the same eligibility, follow-up, outcome, and censoring definitions were applied to both comparisons. Treatment groups were assigned according to the first observed initiation of the respective drug class at baseline, and measured baseline confounding was controlled for using propensity scores. Follow-up started at treatment initiation and continued until incident clinically recorded AD-type dementia, death, loss to follow-up, or the end of the study period, whichever occurred first.

Analyses involving DPP-4 inhibitor initiators are presented only as exploratory supplementary analyses and were not one of the two primary target-trial comparisons.

We used data from the *All of Us Research Program*,^2^ a US-wide prospective cohort designed to investigate biological, environmental, and behavioral determinants of health and disease. The program links longitudinal electronic health records with participant surveys, physical measurements, and biospecimens, which allowed us to reconstruct real-world treatment and outcome trajectories. All participant-facing materials are reviewed by a central Institutional Review Board.

**Supplementary Table S1:** Specification of the two primary target trial emulations using data from the *All of Us Research Program*.

| Protocol component | Target trial specification | Target trial emulation |
| --- | --- | --- |
| **Eligibility criteria** | Adults aged $\geq 55$ years.<br>Established type 2 diabetes.<br>Eligible for initiation of GLP-1 receptor agonist therapy at baseline.<br>New-user design: no use of GLP-1 receptor agonists, SGLT2 inhibitors, DPP-4 inhibitors, or sulfonylureas in the previous 12 months.<br>At least 1 year of observable history before baseline.<br>No prior dementia, AD, or anti-dementia drug use during the one-year look-back.<br>No major contraindications during the one-year look-back.<br>No recorded major cardiovascular or cerebrovascular disease during the fixed 12-month look-back period immediately before baseline.<br>Baseline is defined as the date of first qualifying GLP-1 receptor agonist, SGLT2 inhibitor, or sulfonylurea exposure. | Adults aged $\geq 55$ years.<br>Established type 2 diabetes, defined as having records of a type 2 diabetes diagnosis (ICD-10: E11), thereby excluding individuals with type 1 diabetes. All study participants additionally fulfil the criterion of receiving non-insulin antidiabetic drugs (ATC: A10B) by study design.<br>Without exposure to GLP-1 receptor agonists (ATC: A10BJ), SGLT2 inhibitors (ATC: A10BK), DPP-4 inhibitors (ATC: A10BH), or sulfonylureas (ATC: A10BB) in the 12 months prior to baseline (new-user design).<br>No records of dementia (ICD-10: F00–F03, G30, G31) and no use of anti-dementia drugs including donepezil (N06DA02), rivastigmine (N06DA03), galantamine (N06DA04), or memantine (N06DX01) in the one year prior to baseline.<br>No records of major contraindications during the one year prior to baseline, including inflammatory bowel disease (ICD-10: K50, K51, K52), thyroid cancer (ICD-10: C73), pancreatitis (ICD-10: K85, K86.1), pancreatic cancer (ICD-10: C25), renal dysfunction (ICD-10: N17, N18.3, N18.4, N18.5, N18.9, N19, N00, N01, N03, N04, N05, I12.0, I13.1, I13.2, Y84.1, Z49, Z94.0, Z99.2), hepatic failure (ICD-10: K72), bariatric surgery (procedure codes), and intestinal bypass (ICD-10: Z98.0).<br>No records of major cardiovascular or cerebrovascular disease during the fixed 12-month look-back period immediately before baseline, including ischemic heart disease (ICD-10: I20–I25), heart failure (ICD-10: I50), atrial fibrillation, aortic aneurysm or dissection (ICD-10: I71), or cerebrovascular disease (ICD-10: G45, G46, I60–I63, I65–I69). Diagnoses recorded only before this 12-month window did not constitute an exclusion. This exclusion was removed in a complementary sensitivity analysis.<br>Continuous observation for at least one year prior to baseline.<br>Baseline is defined as the date of first qualifying GLP-1 receptor agonist, SGLT2 inhibitor, or sulfonylurea exposure. |
| **Treatment strategies** | Assignment at baseline to initiation of GLP-1 receptor agonist therapy versus SGLT2 inhibitors or sulfonylureas as active comparators. | Same as the target trial. |
| **Treatment assignment** | Individuals would be assigned at baseline to initiate GLP-1 receptor agonist therapy or to receive SGLT2 inhibitors or sulfonylureas. | No randomization is performed. Individuals are classified as treated if they initiate GLP-1 receptor agonists, and as controls if they initiate SGLT2 inhibitors or sulfonylureas at baseline. |
| **Outcomes** | Time to clinically recorded Alzheimer's disease/dementia after treatment assignment. | Same as the target trial. |
| **Follow-up** | Starts at baseline and continues until clinically AD/dementia, death, loss to follow-up, or administrative end of follow-up. | Same as the target trial. |
| **Causal contrast** | Effect of assignment to initiate GLP-1 receptor agonist therapy versus SGLT2 inhibitors or sulfonylureas at baseline, under the specified eligibility and follow-up rules, regardless of post-baseline discontinuation, switching, treatment intensification, or addition of other glucose-lowering therapies. | Same as the target trial. |
| **Statistical analysis** | Estimate adjusted treatment effects comparing clinically recorded AD-type dementia-free survival between GLP-1 receptor agonist initiators and initiators of SGLT2 inhibitors or sulfonylureas. | Propensity scores are estimated using baseline covariates. The analysis reports balance diagnostics and outcome summaries including numbers at risk, cumulative events, and cumulative censoring over time. |

The study used data from the *All of Us Research Program*, a nationwide EHR-linked cohort mapped to the OMOP Common Data Model. Diagnoses were identified using standardized clinical vocabularies, and medication exposures were captured from prescription records, harmonized to RxNorm ingredient concepts, and linked to ATC drug classes. A fixed one-year look-back period prior to cohort entry was applied to assess baseline characteristics.

### Detailed eligibility criteria

#### Inclusion criteria

Eligibility for both active-comparator target trial emulations was assessed repeatedly within a target trial framework.^1, 3^ At each monthly time point, the risk set included individuals who remained free of the study outcome, were alive and under observation (i.e., had not been censored because of death, loss to follow-up, or the end of data availability), satisfied all eligibility criteria using information available up to that time point, and had not initiated either treatment class during the preceding washout period. Individuals were eligible if they were aged *≥* 55 years at the index date and had a confirmed diagnosis of type 2 diabetes before baseline, defined by at least one ICD-10-CM E11 diagnosis code recorded in the OMOP condition table. To avoid selection bias, all eligibility criteria were assessed at baseline (time zero) using only information available up to the index date. Baseline medical history was ascertained during a fixed one year look-back period preceding each index date.^4^ Individuals could re-enter the risk set at later monthly time points after a 12-month treatment-free washout period, provided that all eligibility criteria were again satisfied. In the first target trial, patients initiating GLP-1 receptor agonists were compared with patients initiating SU. In the second target trial, patients initiating GLP-1 receptor agonists were compared with patients initiating SGLT2 inhibitors. In both target trials, initiation was defined as the first observed prescription of the respective drug class after at least 12 months without prior exposure to GLP-1 receptor agonists, SU, DPP-4 inhibitors, or SGLT2 inhibitors. Exposure was identified using standardized RxNorm ingredient mappings. Among 2,104 GLP-1 receptor agonist initiators, the most common index molecules were semaglutide (32*·*3%), dulaglutide (32*·*2%), and liraglutide (25*·*6%). Comparator groups were defined analogously as new initiators of SU or SGLT2 inhibitors.

#### Exclusion criteria

At each index date, individuals were excluded if they had any recorded exposure to GLP-1 receptor agonists, SU, DPP-4 inhibitors, or SGLT2 inhibitors during the preceding 12-month washout period, ensuring a new-user design in both treatment and comparator groups. We further excluded individuals with evidence of pre-existing clinical AD-type dementia or dementia during the one year look-back period, defined by ICD-10 codes F00–F03, G30, or G31. For the primary analysis, we also excluded individuals with recorded ischemic heart disease, heart failure, atrial fibrillation, aortic aneurysm or dissection, or cerebrovascular disease during the same fixed 12-month look-back period immediately before each index date; cardiovascular or cerebrovascular diagnoses recorded only before that window did not constitute an exclusion. In a complementary sensitivity analysis, we removed this cardiovascular and cerebrovascular exclusion and adjusted for the five component indicators (Supplementary sensitivity analysis). All exclusion criteria for the primary analysis were reassessed at each eligible monthly time point using the same fixed 12-month look-back window.

### Detailed outcome and covariate definitions

#### Outcome definition

The primary outcome was incident clinical AD-type dementia, defined using electronic health record diagnoses or anti-dementia medication dispensing. This endpoint reflects a routine care-based clinical definition rather than biomarker-confirmed AD pathology. Diagnostic evidence compatible with clinical AD-type dementia was identified using ICD-10 codes F00 and G30 (78*·*86% of included AD-type dementia cases). Because AD-type dementia diagnoses may be incompletely captured in routine care, we additionally considered dispensing of anti-dementia medications as evidence of disease onset. These included donepezil, rivastigmine, galantamine, and memantine (21*·*14% of included AD-type dementia cases). The event date was assigned as the earliest occurrence of a qualifying diagnosis or medication dispensing record. Incident clinically recorded AD-type dementia was defined as an outcome occurring after the index date during follow-up, among individuals without a qualifying clinical AD-type dementia diagnosis or anti-dementia medication dispensing during the one year baseline look-back period.

#### Covariate definitions

We included a prespecified set of baseline covariates to adjust for measured confounding and to characterize heterogeneity in AD-related treatment effects. Covariates were selected a priori based on clinical relevance and prior literature^4^ and were measured during the fixed one year look-back period before each index date. Covariates were organized into five domains: (i) demographic and socioeconomic characteristics, (ii) cardiometabolic factors and laboratory measures, (iii) diabetes-related characteristics, (iv) neurological and related clinical factors, and (v) medication history (the full list can be found in Table S2). Diagnoses recorded in the electronic health records were identified using standardized clinical vocabularies, including ICD-10 and SNOMED CT. Medication use was captured from prescription records, harmonized to RxNorm ingredient concepts, and linked to ATC codes to summarize into broader drug classes.

### Detailed causal machine learning analysis

#### Rationale, estimand, and causal survival forest

We used causal machine learning to estimate heterogeneous treatment effects of GLP-1 receptor agonist initiation on clinical AD-type dementia-free survival.^5^ Causal machine learning has several strengths in our setting.^6, 7^ First, it is well-suited to estimate treatment effects from electronic health record data, where treatment assignment is non-random and is driven by clinical indication, disease severity, comorbidity burden, and medication history. Second, it can incorporate highdimensional baseline covariates without requiring strong parametric assumptions. Third, unlike predictive machine learning, causal machine learning explicitly targets treatment effect estimation and can adjust for measured confounding through propensity or doubly robust estimation.

The estimand of interest was the difference in 48-month restricted mean clinical AD-type dementia-free survival time between initiation of GLP-1 receptor agonists and initiation of an active comparator therapy among eligible individuals with type 2 diabetes. Treatment effects were expressed as the difference in RMST between administration of (1) GLP-1 receptor agonists and SU or (2) GLP-1 receptor agonists and SGLT2 inhibitors, expressed in months. Therefore, positive values indicate longer clinical AD-type dementia-free survival under GLP-1 receptor agonists. The first comparison assesses the general benefit of GLP-1 receptor agonists relative to SU as an established reference therapy, while the second uses SGLT2 inhibitors as a modern active comparator to assess whether heterogeneous effects persisted against a glucose-lowering alternative used in overlapping clinical contexts.

Individualized treatment effects for time-to-event outcomes were estimated using causal survival forests (CSurvFs),^8^ a non-parametric causal machine learning model for estimating heterogeneous treatment effects in observational survival data. CSurvFs extend the generalized random forest model to right-censored outcomes, thereby enabling estimation of CATEs without imposing a prespecified functional form on the treatment–outcome relationship.^9, 10^ For estimation, the CSurvF recursively partitions the covariate space to identify regions with differential treatment response. Candidate splits are selected to maximize heterogeneity in estimated CATEs rather than overall outcome prediction accuracy. Individual trees are grown using subsampling and random feature selection, and forest-level estimates were obtained by aggregating predictions across trees. This ensemble procedure stabilizes individualized treatment effects and reduces variance across the covariate distribution.

#### Propensity scores, cross-fitting, and weighting

Confounding due to baseline covariates was addressed using propensity score weighting.^11^ For each active-comparator analysis, propensity scores were estimated using a generalized random forest,^10^ based on prespecified baseline demographic, socioeconomic, cardiometabolic, neurological, and medication covariates. Out-of-fold propensity scores were estimated using generalized random forests and then truncated to the interval [0*·*05, 0*·*95] before construction of inverse probability weights and downstream causal effect estimation, to reduce the influence of extreme treatment probabilities.

To reduce overfitting and improve generalizability, we used stratified five-fold cross-fitting^12^ to estimate out-of-fold propensity scores for each individual. In each fold, the generalized random forest was trained on 80% of the sample and used to predict treatment probabilities in the heldout 20%. This procedure ensured that propensity scores used for downstream weighting were generated from fold-specific models trained on separate subsets of the data.

Different weighting schemes were used as follows. Table 1 reports selected major unweighted baseline characteristics, while Supplementary Table S3 reports pairwise standardized mean differences before and after overlap weighting for the full prespecified covariate set to assess balance in the region of common support. For the primary treatment effect analyses, treatment effects were estimated using doubly robust augmented inverse probability weighting (AIPW) within the causal survival forest framework. IPTW-weighted Kaplan–Meier curves and Cox proportional hazards models were used only as supplementary robustness checks. Thus, overlap weights were used for balance diagnostics, AIPW for the primary causal estimates, and IPTW for sensitivity analyses.

#### Treatment effect heterogeneity

To characterize heterogeneity in treatment response, we used the ITEs predicted by the causal survival forest as effect scores, following Wang et al.^13^ Each effect score represented the predicted individual difference in 48-month restricted mean clinical AD-type dementia-free survival time for GLP-1 receptor agonist initiation versus comparator initiation. Higher scores indicated larger predicted benefit from GLP-1 receptor agonists.

We used these scores to summarize the distribution of predicted benefit and to rank individuals into four equally sized predicted benefit strata. Within each stratum, we estimated the average treatment effect using augmented inverse probability weighting (AIPW), allowing us to compare the model-based ranking with doubly robust stratum-level treatment effects. The overall population-level ATE was estimated analogously using the doubly robust estimator implemented for causal survival forests with cross-fitted propensity scores (see Figures 3 and 5).

Subgroup-level treatment effects were estimated for prespecified clinically relevant strata. Subgroups were defined by age, sex, BMI category, baseline body weight, obesity status, baseline HbA1c, hypertension, metformin use, and insulin use. Age was categorized as <65, 65–70, 70– 75, 75–80, and *≥*80 years. BMI was categorized as <25, 25–30, 30–35, and *≥*35 kg/m^2^. HbA1c was categorized as <7%, 7–8%, 8–9%, and *≥*9%. Baseline body weight was categorized into comparison-specific tertiles among individuals with non-missing baseline body-weight measurements. The exact tertile cut points are reported in Supplementary Table S5. Subgroup-level treatment effects were estimated using doubly robust estimation.^14^ For each subgroup, we reported RMST differences with 95% confidence intervals (CIs) and two-sided *P* -values. Positive values indicate longer clinical AD-type dementia-free survival under GLP-1 receptor agonist initiation relative to the comparator.

### Validation details

#### Model validation

Model validation drew on methodological recommendations and prior applications of causal machine learning and individualized treatment-effect estimation.^6, 13, 15^ First, covariate balance between treatment groups was assessed before and after pairwise overlap weighting using standardized mean differences (SMDs). These weights were used for balance diagnostics and were different from the AIPW estimator used for primary treatment effect estimation. Balance diagnostics were used to evaluate whether propensity score weighting reduced observed baseline differences across demographic, socioeconomic, cardiometabolic, neurological, and medication covariates. Second, we examined propensity score distributions to assess treatment group overlap and identify potential violations of the positivity assumption. Extreme weights were inspected, and the effective sample size after weighting was estimated to quantify the influence of weighting on the information available for treatment effect estimation. Third, propensity score model calibration was assessed using calibration curves and summary metrics. Fourth, following the negative-control method,^16^ we used acute sinusitis, defined from clinical guidance,^17^ as an outcome with no expected effect of GLP-1 receptor agonist initiation. The absence of an estimated treatment effect on this outcome was used as a refutation check to assess whether residual confounding or systematic bias could explain the primary findings. Fifth, to evaluate robustness across alternative modeling strategies, we additionally assessed the consistency of treatment effect patterns using a T-learner instantiated with Cox regression models.^18^ These analyses were conducted as supplementary validation checks (see Supplementary Figures S11 and S12).

### Baseline Covariates

**Supplementary Table S2:** Baseline covariates, exclusion variables, and negative-control outcome definitions with corresponding diagnostic, medication, or measurement codes.

| Domain | Covariate or exclusion variable | Codes or definition |
| --- | --- | --- |
| Demographic and socioeconomic characteristics | Age at baseline | Age in years at index date |
| Demographic and socioeconomic characteristics | Sex at birth | All of Us survey / person-level variable |
| Demographic and socioeconomic characteristics | Race | All of Us survey / person-level variable |
| Demographic and socioeconomic characteristics | Ethnicity | All of Us survey / person-level variable |

Table S2 continued
| Domain | Covariate or exclusion variable | Codes or definition |
| --- | --- | --- |
| Demographic and socioeconomic characteristics | Enrollment year | Year of cohort entry |
| Demographic and socioeconomic characteristics | Income | All of Us survey variable |
| Demographic and socioeconomic characteristics | Education level | All of Us survey variable |
| Demographic and socioeconomic characteristics | Marital status | All of Us survey variable |
| Cardiometabolic factors | Diabetes severity | ICD-10: E11.0–E11.8; treatment proxies |
| Cardiometabolic factors | Hypertension | ICD-10: I10–I15 |
| Cardiometabolic factors | Dyslipidemia | ICD-10: E78 |
| Cardiometabolic factors | Obesity | ICD-10: E65–E66 |
| Cardiometabolic factors | Hypoglycemia | OMOP condition concept IDs: 443727, 4008576, 4185932, 45757458, 4059290, 4230254, 4099651, 36713918 |
| Cardiometabolic factors | Lifestyle-related conditions | ICD-10: Z72 |
| Cardiometabolic factors | Smoking-related conditions | ICD-10: F17 |
| Cardiometabolic factors | Alcohol-related conditions | ICD-10: F10, Y90–Y91 |

Table S2 continued
| Domain | Covariate or exclusion variable | Codes or definition |
| --- | --- | --- |
| Exclusion variables | Prior exposure to GLP-1 receptor agonists, SGLT2 inhibitors, DPP-4 inhibitors, or sulfonylureas during the 12-month washout period | ATC: A10BJ, A10BK, A10BH, A10BB |
| Exclusion variables | Pre-existing dementia or Alzheimer's disease | ICD-10: F00–F03, G30, G31 |
| Exclusion variables | Anti-dementia drug use | ATC: N06DA02, N06DA03, N06DA04, N06DX01 |
| Exclusion variables | Ischemic heart disease | ICD-10: I20–I25 |
| Exclusion variables | Heart failure | ICD-10: I50 |
| Exclusion variables | Atrial fibrillation | ICD-10: I48 |
| Exclusion variables | Aortic aneurysm or dissection | ICD-10: I71 |
| Exclusion variables | Cerebrovascular disease | ICD-10: G45, G46, I60–I63, I65–I69 |
| Exclusion variables | Inflammatory bowel disease | ICD-10: K50, K51, K52 |
| Exclusion variables | Thyroid cancer | ICD-10: C73 |
| Exclusion variables | Pancreatitis | ICD-10: K85, K86.1 |

Table S2 continued
| Domain | Covariate or exclusion variable | Codes or definition |
| --- | --- | --- |
| Exclusion variables | Pancreatic cancer | ICD-10: C25 |
| Exclusion variables | Renal dysfunction | ICD-10: N17, N18.3, N18.4, N18.5, N18.9, N19, N00, N01, N03, N04, N05, I12.0, I13.1, I13.2, Y84.1, Z49, Z94.0, Z99.2 |
| Exclusion variables | Hepatic failure | ICD-10: K72 |
| Exclusion variables | Bariatric surgery | Procedure codes |
| Exclusion variables | Intestinal bypass | ICD-10: Z98.0 |
| *Note:* Exclusion variables were used to define eligibility and were not retained as baseline covariates in the final analytic models. |  |  |
| Negative-control outcome | Acute sinusitis | ICD-10: J01.0–J01.9 |
| *Note:* Acute sinusitis was used only as a negative-control outcome in robustness analyses and was not included as a baseline covariate or exclusion variable. |  |  |
| Laboratory and measurement co-variates | Renal function | eGFR; OMOP measurement concept IDs: 3049187, 3053283, 46236952, 3030354 |
| Laboratory and measurement co-variates | Body mass index | BMI; OMOP measurement concept IDs: 3038553, 4245997 |
| Laboratory and measurement co-variates | Glycated hemoglobin | HbA1c; OMOP measurement concept IDs: 3004410, 3005673, 40789263 |
| Neurological and related clinical factors | Parkinson's disease | ICD-10: G20 |
| Neurological and related clinical factors | Epilepsy | ICD-10: G40 |
| Neurological and related clinical factors | Migraine | ICD-10: G43 |

Table S2 continued
| Domain | Covariate or exclusion variable | Codes or definition |
| --- | --- | --- |
| Neurological and related clinical factors | Intracranial injury | ICD-10: S06 |
| Neurological and related clinical factors | Multiple sclerosis | ICD-10: G35 |
| Neurological and related clinical factors | Huntington's disease | ICD-10: G10 |
| Neurological and related clinical factors | Brain and meningeal tumors | ICD-10: C71, D32–D33, D42–D43 |
| Neurological and related clinical factors | Psychiatric disorders | ICD-10: F04–F99 |
| Neurological and related clinical factors | Hearing loss | ICD-10: H90–H91 |
| Neurological and related clinical factors | Sleep disorders | ICD-10: F51, G47 |
| Neurological and related clinical factors | Cancer excluding non-melanoma skin cancer | ICD-10: C00–C97 excluding C44 |
| Medication history | Antihypertensive agents | ATC: C02 |
| Medication history | Diuretics | ATC: C03 |
| Medication history | Peripheral vasodilators | ATC: C04 |
| Medication history | Vasoprotectives | ATC: C05 |
| Medication history | Beta-blocking agents | ATC: C07 |

Table S2 continued
| Domain | Covariate or exclusion variable | Codes or definition |
| --- | --- | --- |
| Medication history | Calcium channel blockers | ATC: C08 |
| Medication history | Renin-angiotensin system agents | ATC: C09 |
| Medication history | Statins | ATC: C10AA |
| Medication history | Other lipid-lowering agents | ATC: C10AB, C10AC, C10AD, C10AX |
| Medication history | Cardiac therapy | ATC: C01 |
| Medication history | Antiplatelet agents | ATC: B01AC06, B01AC04, B01AC24, B01AC22 |
| Medication history | Anticoagulants | ATC: B01AA03, B01AB01, B01AB05, B01AE07, B01AF01, B01AF02, B01AF03 |
| Medication history | Thrombolytics | ATC: B01AD02, B01AD04, B01AD11 |
| Medication history | Metformin | ATC: A10BA02 |
| Medication history | Insulin | ATC: A10A |
| Medication history | SGLT2 inhibitors | ATC: A10BK |
| Medication history | Thiazolidinediones | ATC: A10BG |
| Medication history | Oral antidiabetic combinations | ATC: A10BD |
| Medication history | Other antidiabetic drugs | ATC: A10BC, A10BF, A10BX |
| Medication history | Antidepressants | ATC: N06A |
| Medication history | Antipsychotics | ATC: N05A |

Table S2 continued
| Domain | Covariate or exclusion variable | Codes or definition |
| --- | --- | --- |
| Medication history | Antiparkinsonian agents | ATC: N04 |
| Medication history | Antiepileptics | ATC: N03A |
| Medication history | Antimigraine drugs | ATC: N02C |
| Medication history | Drugs for amyotrophic lateral sclerosis | ATC: N07XX02, N07XX59 |
| Medication history | Drugs for Huntington's disease | ATC: N07XX06, N07XX10 |

### Full baseline characteristics

Complete baseline distributions and pairwise standardized mean differences before and after overlap weighting are reported below for all prespecified covariates. The corresponding selected major characteristics are shown in Table 1 in the main text.

**Supplementary Table S3:** Full baseline characteristics and pairwise balance diagnostics by treatment group.

| Characteristic | GLP-1RA<br>(n=2104) | SU<br>(n=4224) | SGLT2i<br>(n=966) | SMD (unweighted) |  | SMD (weighted)<sup>†</sup> |  |
| --- | --- | --- | --- | --- | --- | --- | --- |
|  |  |  |  | vs SU | vs SGLT2i | vs SU | vs SGLT2i |
| Demographic and socioeconomic covariates |  |  |  |  |  |  |  |
| Age at baseline, mean (SD) | 63.0 (6.0) | 63.2 (6.3) | 64.9 (6.8) | 0.039 | 0.297 | 0.097 | 0.187 |
| Sex at birth |  |  |  | 0.423 | 0.369 | 0.211 | 0.220 |
| Female | 1429 (67.9%) | 2005 (47.5%) | 488 (50.5%) |  |  |  |  |
| Male | 650 (30.9%) | 2141 (50.7%) | 470 (48.7%) |  |  |  |  |
| Unknown | 25 (1.2%) | 78 (1.8%) | 8 (0.8%) |  |  |  |  |
| Race |  |  |  | 0.025 | 0.130 | 0.072 | 0.077 |
| Amer. Indian/AK Native | 16 (0.8%) | 39 (0.9%) | 12 (1.2%) |  |  |  |  |
| Asian | 31 (1.5%) | 50 (1.2%) | 19 (2.0%) |  |  |  |  |
| Black or African American | 446 (21.2%) | 865 (20.5%) | 156 (16.1%) |  |  |  |  |
| More than one population | 91 (4.3%) | 169 (4.0%) | 35 (3.6%) |  |  |  |  |
| None of these | 26 (1.2%) | 53 (1.3%) | 14 (1.4%) |  |  |  |  |
| Unknown | 349 (16.6%) | 723 (17.1%) | 187 (19.4%) |  |  |  |  |
| White | 1145 (54.4%) | 2325 (55.0%) | 543 (56.2%) |  |  |  |  |
| Ethnicity |  |  |  | 0.031 | 0.067 | 0.024 | 0.057 |
| Hispanic or Latino | 344 (16.3%) | 665 (15.7%) | 173 (17.9%) |  |  |  |  |
| Not Hispanic or Latino | 1695 (80.6%) | 3409 (80.7%) | 752 (77.8%) |  |  |  |  |
| None of these | 26 (1.2%) | 53 (1.3%) | 14 (1.4%) |  |  |  |  |
| Unknown | 39 (1.9%) | 97 (2.3%) | 27 (2.8%) |  |  |  |  |
| Enrollment year, mean (SD) | 2007.1 (7.9) | 2003.8 (8.2) | 2007.7 (8.1) | 0.411 | 0.070 | 0.166 | 0.038 |
| Income |  |  |  | 0.855 | 0.126 | 0.104 | 0.136 |
| Annual income: <\$10k | 161 (7.7%) | 88 (2.1%) | 64 (6.6%) | | | | |
| Annual income: \$10k–25k | 188 (8.9%) | 95 (2.2%) | 72 (7.5%) | | | | |
| Annual income: \$25k–35k | 76 (3.6%) | 43 (1.0%) | 44 (4.6%) | | | | |
| Annual income: \$35k–50k | 88 (4.2%) | 33 (0.8%) | 41 (4.2%) | | | | |
| Annual income: \$50k–75k | 119 (5.7%) | 39 (0.9%) | 63 (6.5%) | | | | |
| Annual income: \$75k–100k | 93 (4.4%) | 27 (0.6%) | 52 (5.4%) | | | | |
| Annual income: \$100k–150k | 95 (4.5%) | 36 (0.9%) | 51 (5.3%) | | | | |
| Annual income: \$150k–200k | 51 (2.4%) | 13 (0.3%) | 8 (0.8%) | | | | |
| Annual income: >\$200k | 47 (2.2%) | 6 (0.1%) | 25 (2.6%) | | | | |
| Unknown | 1186 (56.4%) | 3844 (91.0%) | 546 (56.5%) |  |  |  |  |
| Education |  |  |  | 0.958 | 0.076 | 0.106 | 0.080 |
| Advanced degree | 197 (9.4%) | 61 (1.4%) | 96 (9.9%) |  |  |  |  |
| Continued on next page |  |  |  |  |  |  |  |

Supplementary Table S3 continued from previous page
| Characteristic | GLP-1RA<br>(n=2104) | SU<br>(n=4224) | SGLT2i<br>(n=966) | SMD (unweighted) |  | SMD (weighted)<sup>†</sup> |  |
| --- | --- | --- | --- | --- | --- | --- | --- |
|  |  |  |  | vs SU | vs SGLT2i | vs SU | vs SGLT2i |
| College graduate | 227 (10.8%) | 75 (1.8%) | 110 (11.4%) |  |  |  |  |
| College one to three years | 346 (16.4%) | 140 (3.3%) | 152 (15.7%) |  |  |  |  |
| Twelve or GED | 209 (9.9%) | 122 (2.9%) | 103 (10.7%) |  |  |  |  |
| Nine through eleven | 51 (2.4%) | 42 (1.0%) | 29 (3.0%) |  |  |  |  |
| Five through eight | 35 (1.7%) | 25 (0.6%) | 23 (2.4%) |  |  |  |  |
| One through four | 13 (0.6%) | 14 (0.3%) | 7 (0.7%) |  |  |  |  |
| Never attended | 6 (0.3%) | 2 (0.0%) | 0 (0.0%) |  |  |  |  |
| Unknown | 1020 (48.5%) | 3743 (88.6%) | 446 (46.2%) |  |  |  |  |
| Marital status |  |  |  | 0.946 | 0.094 | 0.084 | 0.077 |
| Divorced | 238 (11.3%) | 109 (2.6%) | 112 (11.6%) |  |  |  |  |
| Living with partner | 43 (2.0%) | 14 (0.3%) | 19 (2.0%) |  |  |  |  |
| Married | 458 (21.8%) | 193 (4.6%) | 249 (25.8%) |  |  |  |  |
| Never married | 185 (8.8%) | 75 (1.8%) | 89 (9.2%) |  |  |  |  |
| Separated | 61 (2.9%) | 32 (0.8%) | 26 (2.7%) |  |  |  |  |
| Widowed | 89 (4.2%) | 59 (1.4%) | 37 (3.8%) |  |  |  |  |
| Unknown | 1030 (49.0%) | 3742 (88.6%) | 434 (44.9%) |  |  |  |  |
| *Cardiometabolic risk factors* |  |  |  |  |  |  |  |
| Obesity | 885 (42.1%) | 433 (10.3%) | 221 (22.9%) | 0.809 | 0.408 | 0.342 | 0.241 |
| BMI, mean (SD) | 37.2 (7.7) | 33.6 (7.1) | 33.1 (6.6) | 0.492 | 0.568 | 0.362 | 0.361 |
| Hypertension | 1151 (54.7%) | 1178 (27.9%) | 550 (56.9%) | 0.566 | 0.045 | 0.072 | 0.088 |
| Dyslipidemia | 1043 (49.6%) | 1010 (23.9%) | 511 (52.9%) | 0.552 | 0.067 | 0.115 | 0.094 |
| Smoking/substance abuse | 106 (5.0%) | 151 (3.6%) | 48 (5.0%) | 0.074 | 0.008 | 0.082 | 0.028 |
| Alcoholism | 32 (1.5%) | 39 (0.9%) | 15 (1.6%) | 0.054 | 0.003 | 0.033 | 0.013 |
| Lifestyle-related conditions | 10 (0.5%) | 8 (0.2%) | 11 (1.1%) | 0.051 | 0.081 | 0.013 | 0.083 |
| *Laboratory values* |  |  |  |  |  |  |  |
| HbA1c (%), mean (SD) | 7.6 (1.8) | 8.1 (1.8) | 7.9 (1.6) | 0.271 | 0.179 | 0.194 | 0.100 |
| eGFR (mL/min/1.73m<sup>2</sup>), mean (SD) | 72.1 (20.2) | 73.0 (22.1) | 73.7 (19.2) | 0.042 | 0.079 | 0.112 | 0.048 |
| *Diabetes-related characteristics* |  |  |  |  |  |  |  |
| Diabetes severity score (0–4) |  |  |  | 0.404 | 0.091 | 0.071 | 0.109 |
| Score 0 | 1326 (63.0%) | 3414 (80.8%) | 566 (58.6%) |  |  |  |  |
| Score 1 | 513 (24.4%) | 573 (13.6%) | 268 (27.7%) |  |  |  |  |
| Score 2 | 184 (8.7%) | 180 (4.3%) | 96 (9.9%) |  |  |  |  |
| Score 3 | 63 (3.0%) | 45 (1.1%) | 27 (2.8%) |  |  |  |  |
| Score 4 | 18 (0.9%) | 12 (0.3%) | 9 (0.9%) |  |  |  |  |
| Hypoglycemia history | 636 (30.2%) | 1108 (26.2%) | 236 (24.4%) | 0.089 | 0.130 | 0.057 | 0.085 |
| *Neurological and psychiatric conditions* |  |  |  |  |  |  |  |
| Psychiatric disorders | 676 (32.1%) | 591 (14.0%) | 252 (26.1%) | 0.441 | 0.133 | 0.085 | 0.051 |
| Sleep disorders | 589 (28.0%) | 355 (8.4%) | 209 (21.6%) | 0.543 | 0.127 | 0.199 | 0.061 |
| Migraine | 99 (4.7%) | 48 (1.1%) | 24 (2.5%) | 0.213 | 0.120 | 0.095 | 0.076 |
| Parkinson's disease | 7 (0.3%) | 5 (0.1%) | 0 (0.0%) | 0.046 | 0.084 | 0.016 | 0.088 |
| Epilepsy | 15 (0.7%) | 10 (0.2%) | 4 (0.4%) | 0.069 | 0.040 | 0.023 | 0.028 |
| Multiple sclerosis | 10 (0.5%) | 6 (0.1%) | 4 (0.4%) | 0.062 | 0.006 | 0.033 | 0.003 |
| Continued on next page |  |  |  |  |  |  |  |

Supplementary Table S3 continued from previous page
| Characteristic | GLP-1RA<br>(n=2104) | SU<br>(n=4224) | SGLT2i<br>(n=966) | SMD (unweighted) |  | SMD (weighted)<sup>†</sup> |  |
| --- | --- | --- | --- | --- | --- | --- | --- |
|  |  |  |  | vs SU | vs SGLT2i | vs SU | vs SGLT2i |
| Intracranial injury | 10 (0.5%) | 7 (0.2%) | 3 (0.3%) | 0.056 | 0.024 | 0.000 | 0.009 |
| Brain and meningeal tumors | 10 (0.5%) | 8 (0.2%) | 7 (0.7%) | 0.051 | 0.037 | 0.006 | 0.044 |
| *Other comorbidities* |  |  |  |  |  |  |  |
| Hearing loss | 111 (5.3%) | 89 (2.1%) | 54 (5.6%) | 0.173 | 0.026 | 0.078 | 0.029 |
| Cancer excl. non-melanoma skin | 141 (6.7%) | 185 (4.4%) | 88 (9.1%) | 0.105 | 0.107 | 0.021 | 0.097 |
| Huntington's disease | 1 (0.0%) | 0 (0.0%) | 0 (0.0%) | 0.032 | 0.032 | 0.034 | 0.033 |
| *Concomitant medications* |  |  |  |  |  |  |  |
| Antihypertensive agents (C02) | 375 (17.8%) | 883 (20.9%) | 130 (13.5%) | 0.078 | 0.120 | 0.018 | 0.069 |
| RAS agents (C09) | 1036 (49.2%) | 2501 (59.2%) | 481 (49.8%) | 0.201 | 0.011 | 0.117 | 0.008 |
| Beta-blocking agents (C07) | 496 (23.6%) | 1468 (34.8%) | 234 (24.2%) | 0.248 | 0.015 | 0.125 | 0.044 |
| Calcium channel blockers (C08) | 557 (26.5%) | 1301 (30.8%) | 243 (25.2%) | 0.096 | 0.030 | 0.056 | 0.003 |
| Diuretics (C03) | 698 (33.2%) | 1603 (37.9%) | 265 (27.4%) | 0.100 | 0.125 | 0.043 | 0.075 |
| Peripheral vasodilators (C04) | 53 (2.5%) | 217 (5.1%) | 14 (1.4%) | 0.137 | 0.077 | 0.069 | 0.053 |
| Vasoprotectives (C05) | 760 (36.1%) | 1452 (34.4%) | 303 (31.4%) | 0.037 | 0.101 | 0.005 | 0.031 |
| Cardiac therapies (C01) | 605 (28.8%) | 1215 (28.8%) | 209 (21.6%) | 0.000 | 0.165 | 0.021 | 0.089 |
| Antiplatelet agents | 89 (4.2%) | 401 (9.5%) | 32 (3.3%) | 0.209 | 0.048 | 0.100 | 0.030 |
| Anticoagulants | 209 (9.9%) | 537 (12.7%) | 84 (8.7%) | 0.088 | 0.043 | 0.108 | 0.002 |
| Statins (C10AA) | 1106 (52.6%) | 2445 (57.9%) | 576 (59.6%) | 0.107 | 0.143 | 0.056 | 0.122 |
| Other lipid-lowering agents | 407 (19.3%) | 1054 (25.0%) | 149 (15.4%) | 0.135 | 0.104 | 0.028 | 0.058 |
| Antidepressants (N06A) | 698 (33.2%) | 1087 (25.7%) | 262 (27.1%) | 0.164 | 0.132 | 0.119 | 0.074 |
| Antipsychotics (N05A) | 177 (8.4%) | 294 (7.0%) | 48 (5.0%) | 0.055 | 0.138 | 0.015 | 0.089 |
| Antiparkinsonian agents (N04) | 170 (8.1%) | 283 (6.7%) | 50 (5.2%) | 0.053 | 0.117 | 0.010 | 0.078 |
| Antiepileptics (N03A) | 589 (28.0%) | 938 (22.2%) | 210 (21.7%) | 0.134 | 0.145 | 0.080 | 0.092 |
| Antimigraine drugs (N02C) | 339 (16.1%) | 663 (15.7%) | 111 (11.5%) | 0.011 | 0.134 | 0.037 | 0.073 |
| Thiazolidinediones | 53 (2.5%) | 251 (5.9%) | 20 (2.1%) | 0.171 | 0.030 | 0.082 | 0.029 |
| Drugs for ALS (N07XX) | 217 (10.3%) | 610 (14.4%) | 74 (7.7%) | 0.126 | 0.093 | 0.072 | 0.049 |
| Thrombolytics (acute stroke) | 4 (0.2%) | 19 (0.4%) | 2 (0.2%) | 0.046 | 0.004 | 0.042 | 0.010 |
| *Prior glucose-lowering therapy (one year look-back)* |  |  |  |  |  |  |  |
| Metformin | 1060 (50.4%) | 2682 (63.5%) | 535 (55.4%) | 0.267 | 0.100 | 0.222 | 0.082 |
| Insulin | 688 (32.7%) | 863 (20.4%) | 241 (24.9%) | 0.280 | 0.172 | 0.231 | 0.156 |
| Oral antidiabetic combinations. | 207 (9.8%) | 890 (21.1%) | 128 (13.3%) | 0.315 | 0.107 | 0.197 | 0.109 |
| Other antidiabetic agents | 21 (1.0%) | 40 (0.9%) | 6 (0.6%) | 0.005 | 0.042 | 0.025 | 0.039 |
| SMD, standardized mean difference; SD, standard deviation. <sup>†</sup> Weighted SMDs were computed using pairwise overlap weights derived from cross-fitted propensity forests and are reported for covariate balance diagnostics only. Primary treatment-effect estimates were obtained using augmented inverse probability weighting (AIPW); IPTW-weighted Kaplan–Meier and Cox models were used as supplementary robustness checks. |  |  |  |  |  |  |  |

### Causal machine learning estimates

**Supplementary Figure S1:**
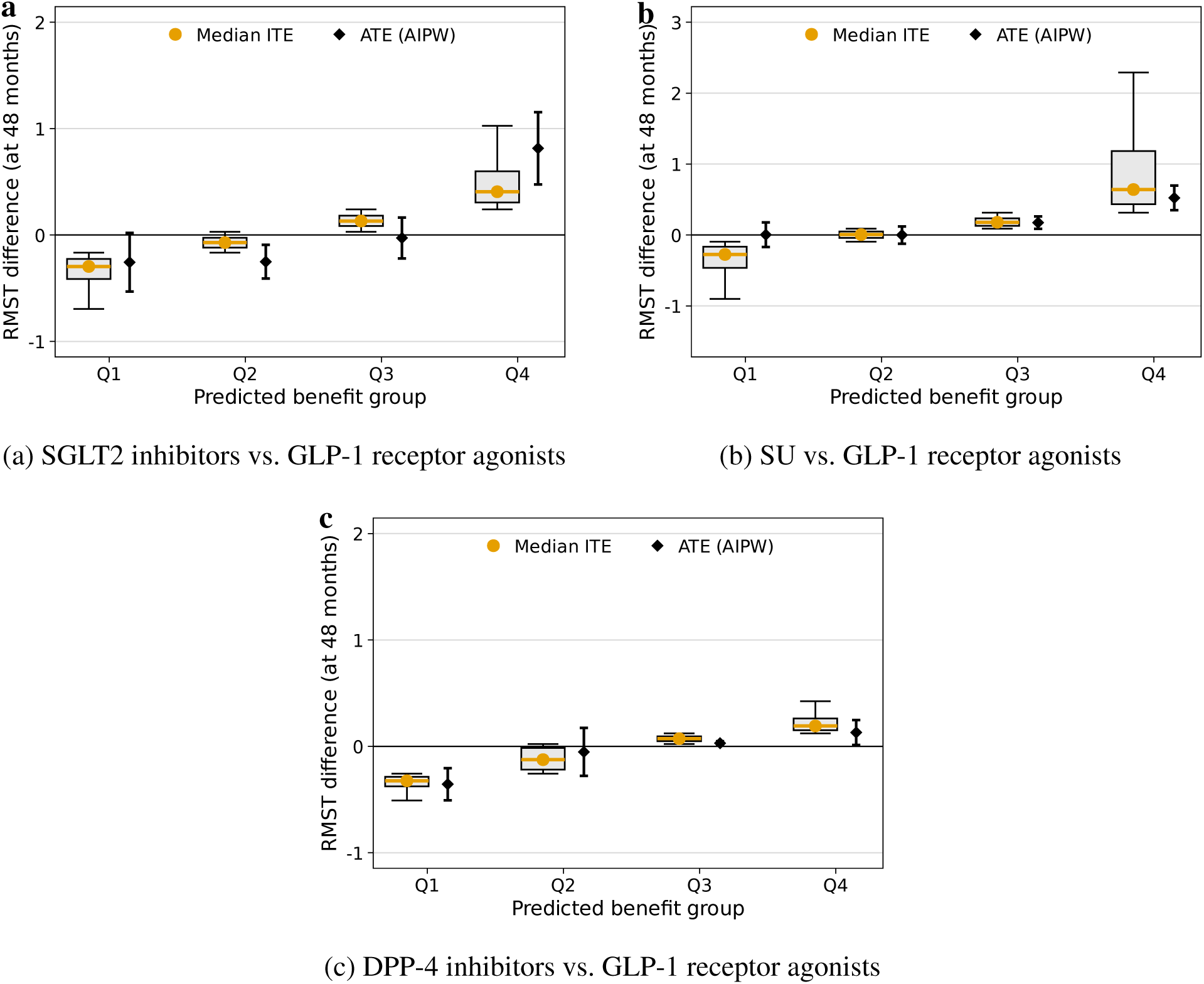
Causal machine learning estimates of treatment effect heterogeneity. RMST differences at 48 months are shown across predicted benefit groups (Q1–Q4) for **(a)** GLP-1 receptor agonists versus SGLT2 inhibitors, **(b)** GLP-1 receptor agonists versus SU, and **(c)** GLP-1 receptor agonists versus DPP-4 inhibitors. Values are expressed as GLP-1 receptor agonist initiation minus comparator initiation. Positive values therefore indicate longer predicted clinical AD-type dementia-free survival under GLP-1 receptor agonists, whereas negative values indicate longer predicted clinical AD-type dementia-free survival under the comparator. Points represent median individualized treatment effects, and diamonds represent average treatment effects estimated using augmented inverse probability weighting (AIPW).

### Cumulative hazard of AD

**Supplementary Figure S2:**
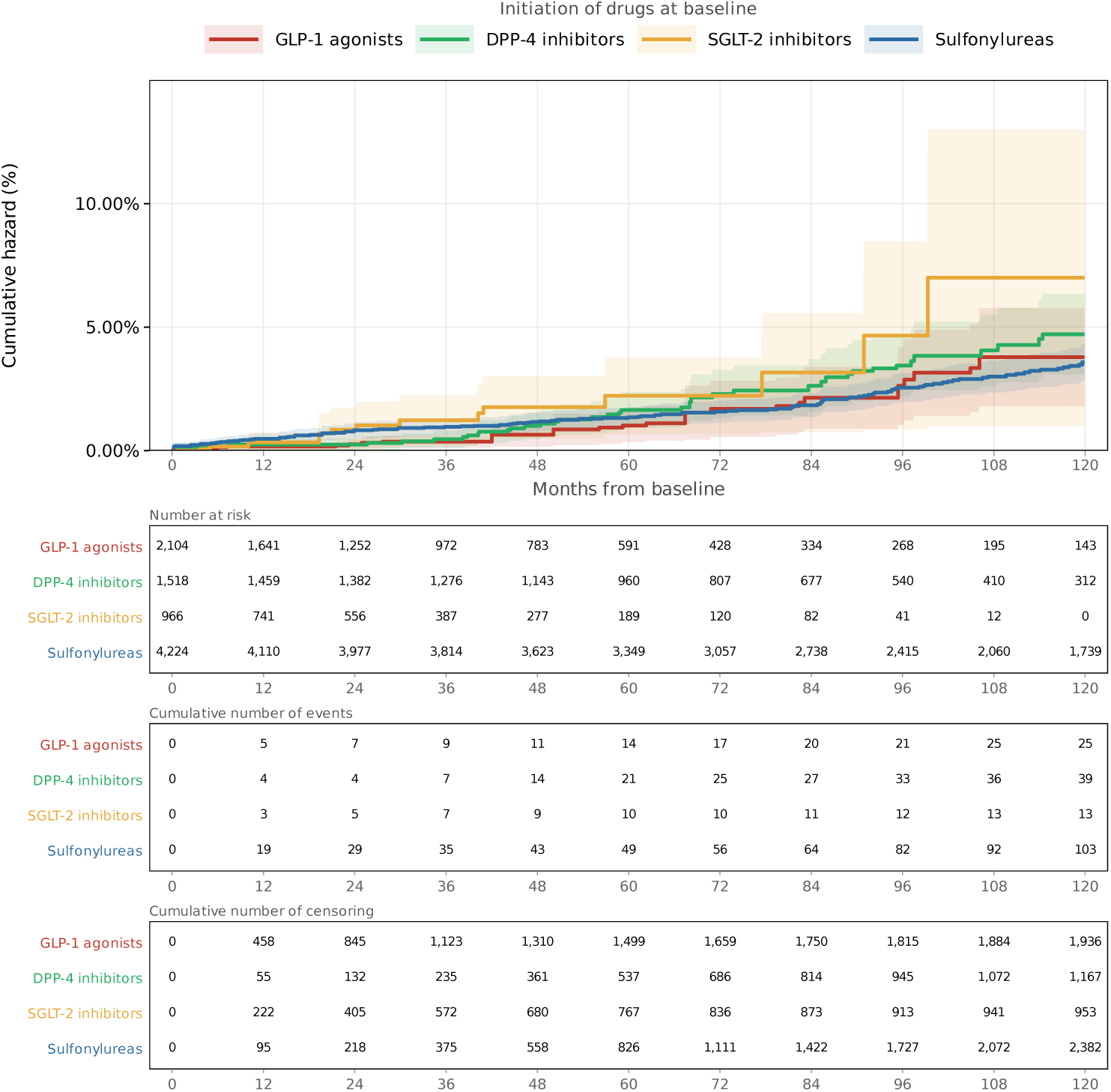
Cumulative hazard of clinical AD-type dementia by antidiabetic drug class. Nelson–Aalen cumulative hazard estimates for incident clinically recorded AD-type dementia are shown for GLP-1 receptor agonist (*n* = 2,104), DPP-4 inhibitor (*n* = 1,518), SGLT2 inhibitor (*n* = 966), and SU (*n* = 4,224) initiators. Shaded areas represent 95% confidence intervals. Stabilized inverse probability of treatment weighting (IPTW) was applied to adjust for baseline differences between groups. The number at risk, cumulative events, and cumulative censoring are shown below the main panel at 12-month intervals. Abbreviations: AD, Alzheimer’s disease; DPP4, dipeptidyl peptidase-4; GLP-1, glucagon-like peptide-1; IPTW, inverse probability of treatment weighting; SGLT2, sodium–glucose cotransporter-2; SU, sulfonylurea.

### RATE

**Supplementary Figure S3:**
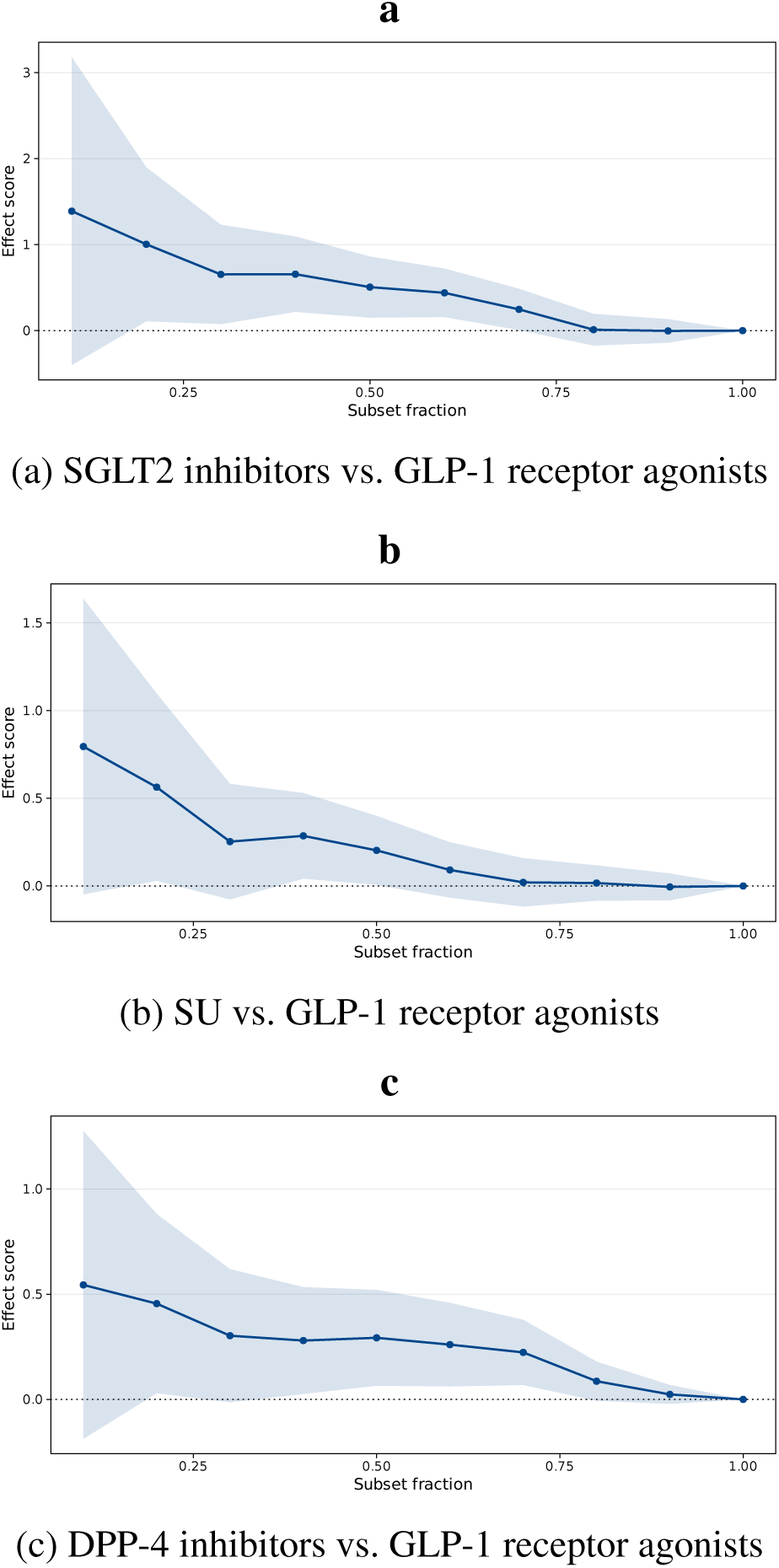
Rank-weighted average treatment effect curves. Targeting operator characteristic (TOC) curves for **(a)** SGLT2 inhibitors versus GLP-1 receptor agonists, **(b)** SU versus GLP-1 receptor agonists, and **(c)** DPP-4 inhibitors versus GLP-1 receptor agonists. The effect score represents the average treatment effect among the top fraction of patients ranked by predicted ITE. A monotonically decreasing curve above zero indicates that patients ranked highest by the model derive the greatest benefit, consistent with clinically meaningful heterogeneity in treatment response. Shaded bands represent *±*1 standard error. The dotted horizontal line denotes the null of no differential benefit.

### C for benefit

To assess the ability of the causal survival forest (CSurvF) to identify individuals with differential treatment benefit, we computed the C-for-benefit statistic, which quantifies the concordance between predicted ITEs and observed pairwise treatment differences in outcomes.

For the comparison of SU versus GLP-1 receptor agonists, the C-for-benefit was 0*·*578 (*n* = 6,328; NNT *≈* 202 events), indicating discriminative ability of the model to stratify patients by predicted treatment benefit. For the comparison of SGLT2 inhibitors versus GLP-1 receptor agonists, the C-for-benefit was 0*·*637 (*n* = 3,070; NNT *≈* 245), the highest across all three comparisons, suggesting that the CSurvF captures meaningful variation in the relative benefit of GLP-1 receptor agonists over SGLT2 inhibitors despite the limited event count. While this estimate should be interpreted cautiously given the small number of events, which may introduce instability in the concordance estimate, the value above 0*·*5 provides preliminary evidence of genuine heterogeneity. For the comparison of DPP-4 inhibitors versus GLP-1 receptor agonists, the C-for-benefit was 0*·*521 (*n* = 3,622; 78 events), approaching the chance level of 0*·*5 and suggesting limited discriminative ability in this comparison, consistent with the near-null average treatment effect observed.

### Clinical characteristics by predicted benefit direction

**Supplementary Figure S4:**
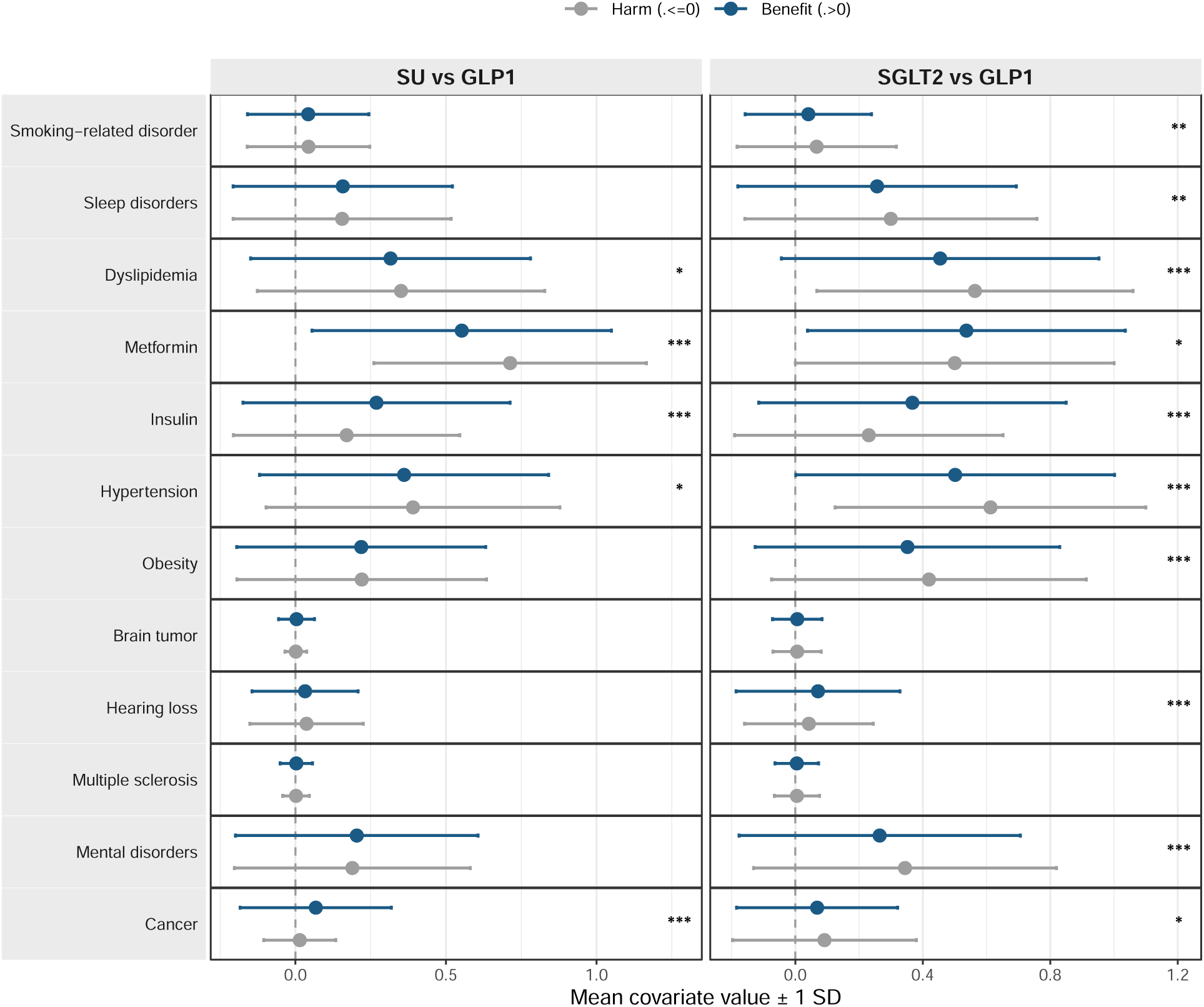
Clinical characteristics associated with predicted treatment benefit versus harm. Patients were stratified according to the direction of the predicted treatment effect (*τ >* 0 vs. *τ ≤* 0). Points represent mean covariate values, with error bars indicating *±*1 SD. In the comparison of SU versus GLP-1 receptor agonists, most patients were predicted to benefit, whereas in the comparison with SGLT2 inhibitors, benefit and harm were more evenly distributed. Stars indicate nominal two-sided Wilcoxon rank-sum tests comparing covariate values between patients with predicted benefit (*τ >* 0) and predicted harm or no benefit (*τ ≤* 0) within each treatment comparison: \**P <* 0*·*05, \*\**P <* 0*·*01, and \*\*\**P <* 0*·*001. These tests were exploratory and not adjusted for multiplicity.

### Secondary Outcomes

**Supplementary Table S4:**
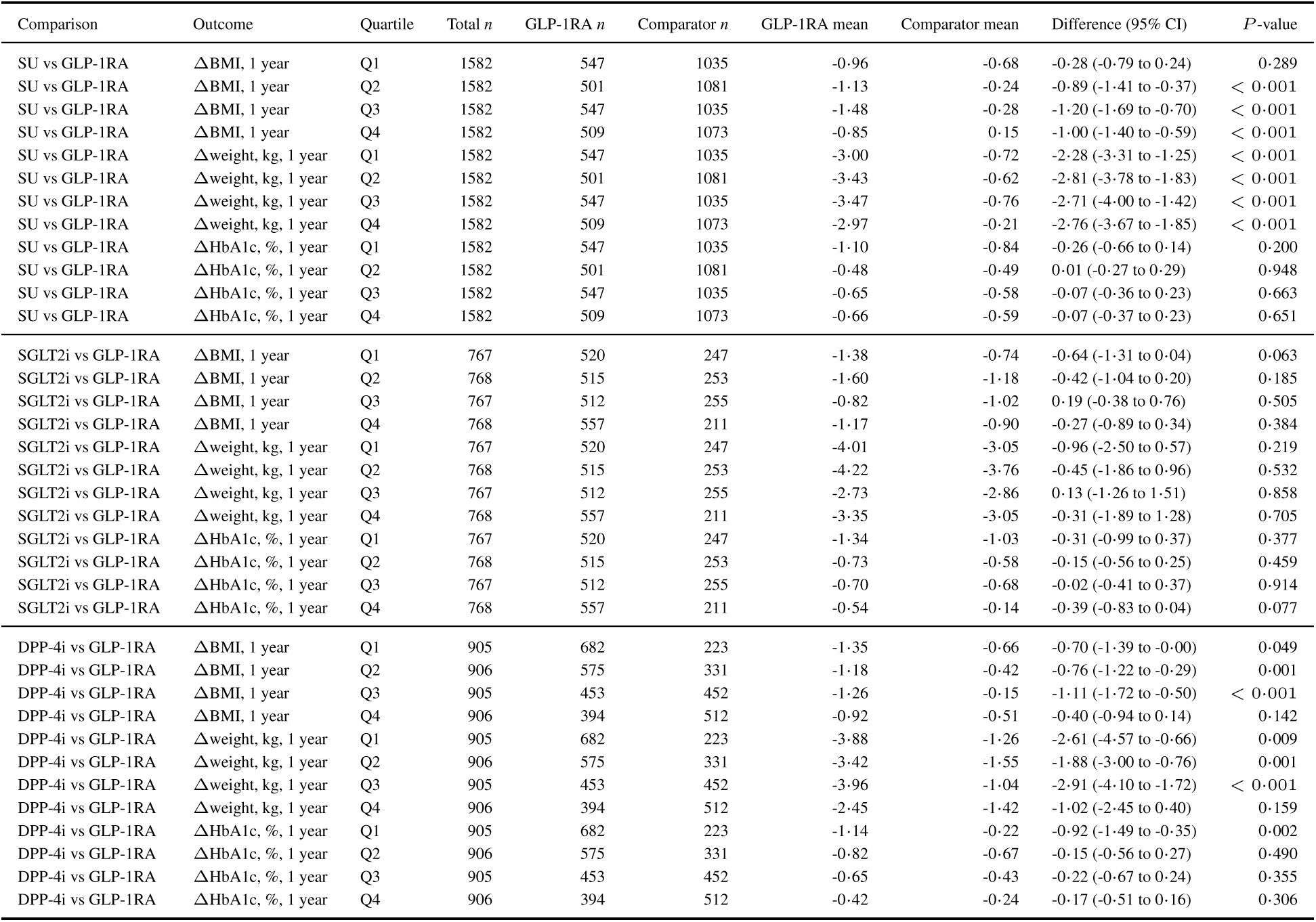
Secondary continuous outcomes across primary AD-CATE quartiles. Quartiles were defined according to predicted ITEs for clinical AD-type dementia-free survival in the primary model. Secondary outcomes were defined as changes from baseline to the one year follow-up measurement. Values are inverse probability weighted means among individuals with observed secondary outcome measurements. Differences are GLP-1 receptor agonists minus comparator. Total *n* denotes the number of individuals in the corresponding primary AD-CATE quartile. GLP-1RA *n* and comparator *n* denote the observed numbers contributing to each quartile-specific comparison.

| Comparison | Outcome | Quartile | Total $n$ | GLP-1RA $n$ | Comparator $n$ | GLP-1RA mean | Comparator mean | Difference (95% CI) | $P$-value |
| --- | --- | --- | --- | --- | --- | --- | --- | --- | --- |
| SU vs GLP-1RA | ΔBMI, 1 year | Q1 | 1582 | 547 | 1035 | -0.96 | -0.68 | -0.28 (-0.79 to 0.24) | 0.289 |
| SU vs GLP-1RA | ΔBMI, 1 year | Q2 | 1582 | 501 | 1081 | -1.13 | -0.24 | -0.89 (-1.41 to -0.37) | < 0.001 |
| SU vs GLP-1RA | ΔBMI, 1 year | Q3 | 1582 | 547 | 1035 | -1.48 | -0.28 | -1.20 (-1.69 to -0.70) | < 0.001 |
| SU vs GLP-1RA | ΔBMI, 1 year | Q4 | 1582 | 509 | 1073 | -0.85 | 0.15 | -1.00 (-1.40 to -0.59) | < 0.001 |
| SU vs GLP-1RA | Δweight, kg, 1 year | Q1 | 1582 | 547 | 1035 | -3.00 | -0.72 | -2.28 (-3.31 to -1.25) | < 0.001 |
| SU vs GLP-1RA | Δweight, kg, 1 year | Q2 | 1582 | 501 | 1081 | -3.43 | -0.62 | -2.81 (-3.78 to -1.83) | < 0.001 |
| SU vs GLP-1RA | Δweight, kg, 1 year | Q3 | 1582 | 547 | 1035 | -3.47 | -0.76 | -2.71 (-4.00 to -1.42) | < 0.001 |
| SU vs GLP-1RA | Δweight, kg, 1 year | Q4 | 1582 | 509 | 1073 | -2.97 | -0.21 | -2.76 (-3.67 to -1.85) | < 0.001 |
| SU vs GLP-1RA | ΔHbA1c, %, 1 year | Q1 | 1582 | 547 | 1035 | -1.10 | -0.84 | -0.26 (-0.66 to 0.14) | 0.200 |
| SU vs GLP-1RA | ΔHbA1c, %, 1 year | Q2 | 1582 | 501 | 1081 | -0.48 | -0.49 | 0.01 (-0.27 to 0.29) | 0.948 |
| SU vs GLP-1RA | ΔHbA1c, %, 1 year | Q3 | 1582 | 547 | 1035 | -0.65 | -0.58 | -0.07 (-0.36 to 0.23) | 0.663 |
| SU vs GLP-1RA | ΔHbA1c, %, 1 year | Q4 | 1582 | 509 | 1073 | -0.66 | -0.59 | -0.07 (-0.37 to 0.23) | 0.651 |
| SGLT2i vs GLP-1RA | ΔBMI, 1 year | Q1 | 767 | 520 | 247 | -1.38 | -0.74 | -0.64 (-1.31 to 0.04) | 0.063 |
| SGLT2i vs GLP-1RA | ΔBMI, 1 year | Q2 | 768 | 515 | 253 | -1.60 | -1.18 | -0.42 (-1.04 to 0.20) | 0.185 |
| SGLT2i vs GLP-1RA | ΔBMI, 1 year | Q3 | 767 | 512 | 255 | -0.82 | -1.02 | 0.19 (-0.38 to 0.76) | 0.505 |
| SGLT2i vs GLP-1RA | ΔBMI, 1 year | Q4 | 768 | 557 | 211 | -1.17 | -0.90 | -0.27 (-0.89 to 0.34) | 0.384 |
| SGLT2i vs GLP-1RA | Δweight, kg, 1 year | Q1 | 767 | 520 | 247 | -4.01 | -3.05 | -0.96 (-2.50 to 0.57) | 0.219 |
| SGLT2i vs GLP-1RA | Δweight, kg, 1 year | Q2 | 768 | 515 | 253 | -4.22 | -3.76 | -0.45 (-1.86 to 0.96) | 0.532 |
| SGLT2i vs GLP-1RA | Δweight, kg, 1 year | Q3 | 767 | 512 | 255 | -2.73 | -2.86 | 0.13 (-1.26 to 1.51) | 0.858 |
| SGLT2i vs GLP-1RA | Δweight, kg, 1 year | Q4 | 768 | 557 | 211 | -3.35 | -3.05 | -0.31 (-1.89 to 1.28) | 0.705 |
| SGLT2i vs GLP-1RA | ΔHbA1c, %, 1 year | Q1 | 767 | 520 | 247 | -1.34 | -1.03 | -0.31 (-0.99 to 0.37) | 0.377 |
| SGLT2i vs GLP-1RA | ΔHbA1c, %, 1 year | Q2 | 768 | 515 | 253 | -0.73 | -0.58 | -0.15 (-0.56 to 0.25) | 0.459 |
| SGLT2i vs GLP-1RA | ΔHbA1c, %, 1 year | Q3 | 767 | 512 | 255 | -0.70 | -0.68 | -0.02 (-0.41 to 0.37) | 0.914 |
| SGLT2i vs GLP-1RA | ΔHbA1c, %, 1 year | Q4 | 768 | 557 | 211 | -0.54 | -0.14 | -0.39 (-0.83 to 0.04) | 0.077 |
| DPP-4i vs GLP-1RA | ΔBMI, 1 year | Q1 | 905 | 682 | 223 | -1.35 | -0.66 | -0.70 (-1.39 to -0.00) | 0.049 |
| DPP-4i vs GLP-1RA | ΔBMI, 1 year | Q2 | 906 | 575 | 331 | -1.18 | -0.42 | -0.76 (-1.22 to -0.29) | 0.001 |
| DPP-4i vs GLP-1RA | ΔBMI, 1 year | Q3 | 905 | 453 | 452 | -1.26 | -0.15 | -1.11 (-1.72 to -0.50) | < 0.001 |
| DPP-4i vs GLP-1RA | ΔBMI, 1 year | Q4 | 906 | 394 | 512 | -0.92 | -0.51 | -0.40 (-0.94 to 0.14) | 0.142 |
| DPP-4i vs GLP-1RA | Δweight, kg, 1 year | Q1 | 905 | 682 | 223 | -3.88 | -1.26 | -2.61 (-4.57 to -0.66) | 0.009 |
| DPP-4i vs GLP-1RA | Δweight, kg, 1 year | Q2 | 906 | 575 | 331 | -3.42 | -1.55 | -1.88 (-3.00 to -0.76) | 0.001 |
| DPP-4i vs GLP-1RA | Δweight, kg, 1 year | Q3 | 905 | 453 | 452 | -3.96 | -1.04 | -2.91 (-4.10 to -1.72) | < 0.001 |
| DPP-4i vs GLP-1RA | Δweight, kg, 1 year | Q4 | 906 | 394 | 512 | -2.45 | -1.42 | -1.02 (-2.45 to 0.40) | 0.159 |
| DPP-4i vs GLP-1RA | ΔHbA1c, %, 1 year | Q1 | 905 | 682 | 223 | -1.14 | -0.22 | -0.92 (-1.49 to -0.35) | 0.002 |
| DPP-4i vs GLP-1RA | ΔHbA1c, %, 1 year | Q2 | 906 | 575 | 331 | -0.82 | -0.67 | -0.15 (-0.56 to 0.27) | 0.490 |
| DPP-4i vs GLP-1RA | ΔHbA1c, %, 1 year | Q3 | 905 | 453 | 452 | -0.65 | -0.43 | -0.22 (-0.67 to 0.24) | 0.355 |
| DPP-4i vs GLP-1RA | ΔHbA1c, %, 1 year | Q4 | 906 | 394 | 512 | -0.42 | -0.24 | -0.17 (-0.51 to 0.16) | 0.306 |

### Concordance-Discordance

**Supplementary Figure S5:**
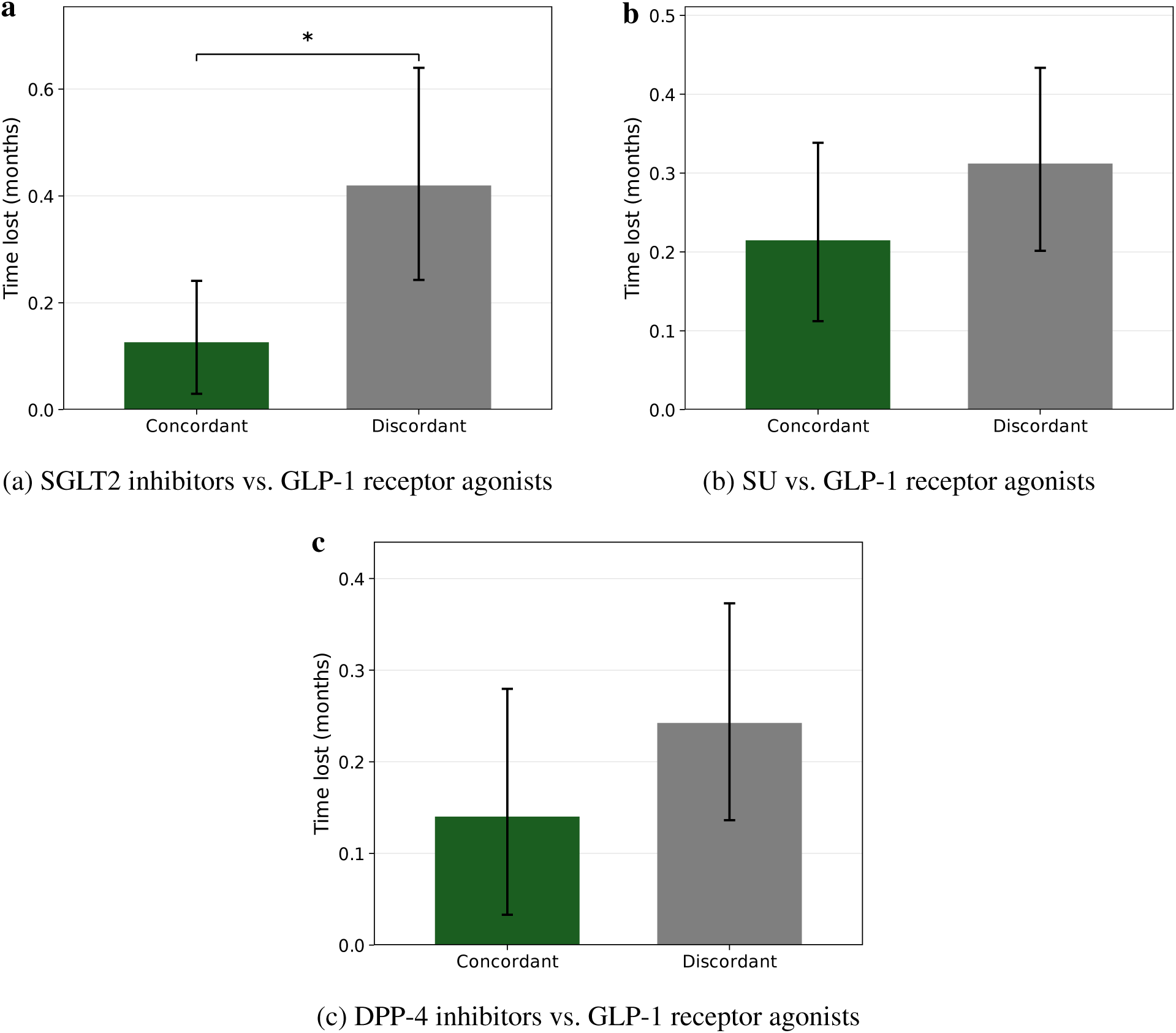
Time lost to clinical AD-type dementia in concordant and discordant patient pairs. Distribution of clinical AD-type dementia-free time lost in months, stratified by concordance status, for **(a)** GLP-1 receptor agonists versus SGLT2 inhibitors, **(b)** GLP-1 receptor agonists versus SU, and **(c)** GLP-1 receptor agonists versus DPP-4 inhibitors. Individualized treatment effects were estimated using the causal survival forest. Concordant pairs were defined as pairs in which the individual with the higher predicted benefit from GLP-1 receptor agonist initiation experienced the longer clinical AD-type dementia-free survival time. Discordant pairs were defined as pairs in which the individual with the lower predicted benefit experienced the longer clinical AD-type dementia-free survival time. Boxes show the interquartile range and median; violin shapes represent the full distribution. Differences between groups were assessed using the Wilcoxon rank-sum test.

### Negative Control Outcome Analysis

**Supplementary Figure S6:**
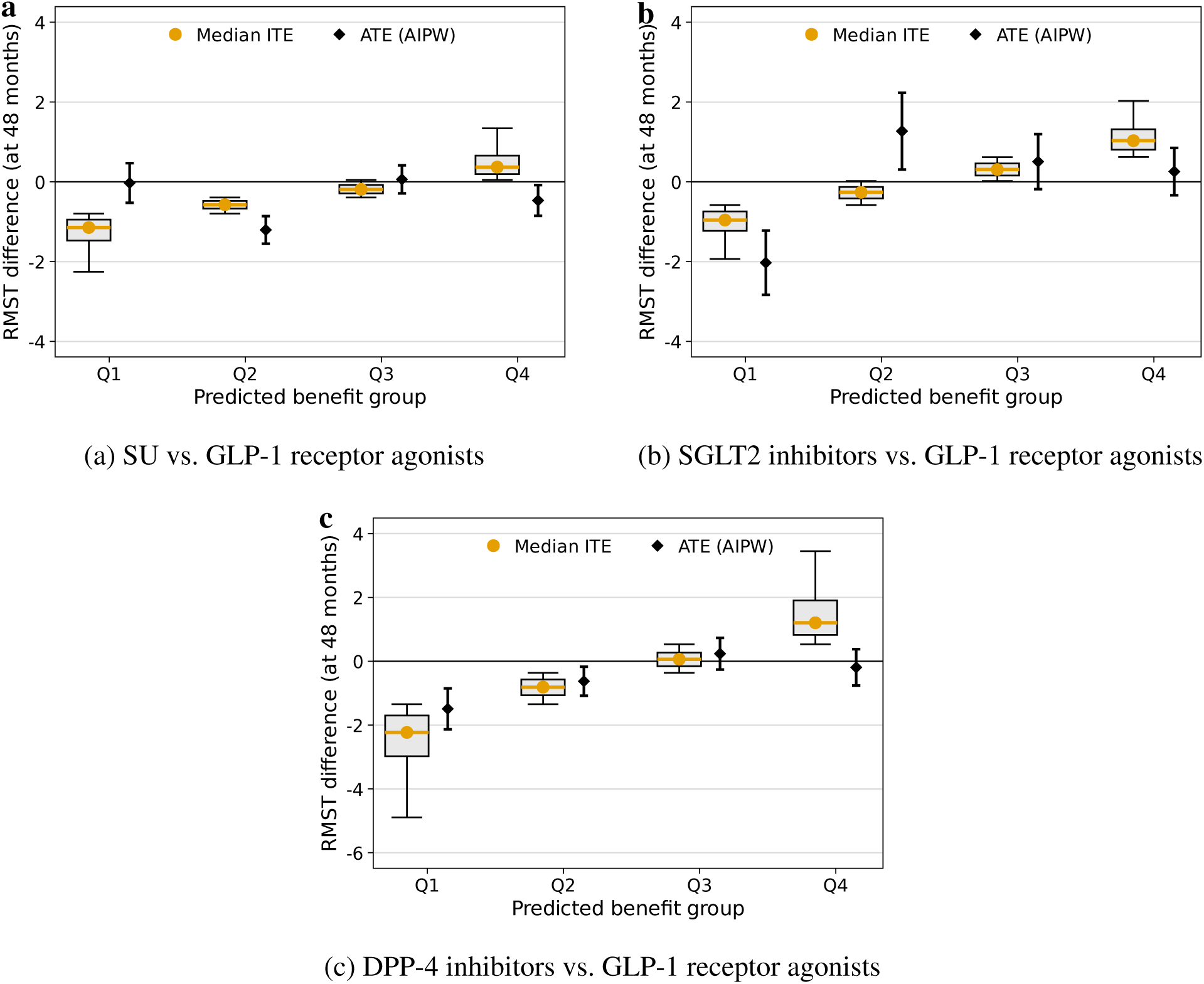
Negative control analysis: acute sinusitis as outcome. Predicted benefit quartiles and augmented inverse probability-weighted (AIPW) average treatment effect (ATE) estimates for acute sinusitis incidence across treatment comparisons. Acute sinusitis has no plausible biological relationship to the antidiabetic drug classes under study and was used as a negative control outcome to assess residual confounding. Absence of a monotone quartile gradient and ATEs close to zero across all three comparisons did not provide evidence of a similar systematic association for this negative-control outcome. However, this analysis cannot rule out residual confounding in the primary analysis.^19^

### CATE Histogram

**Supplementary Figure S7:**
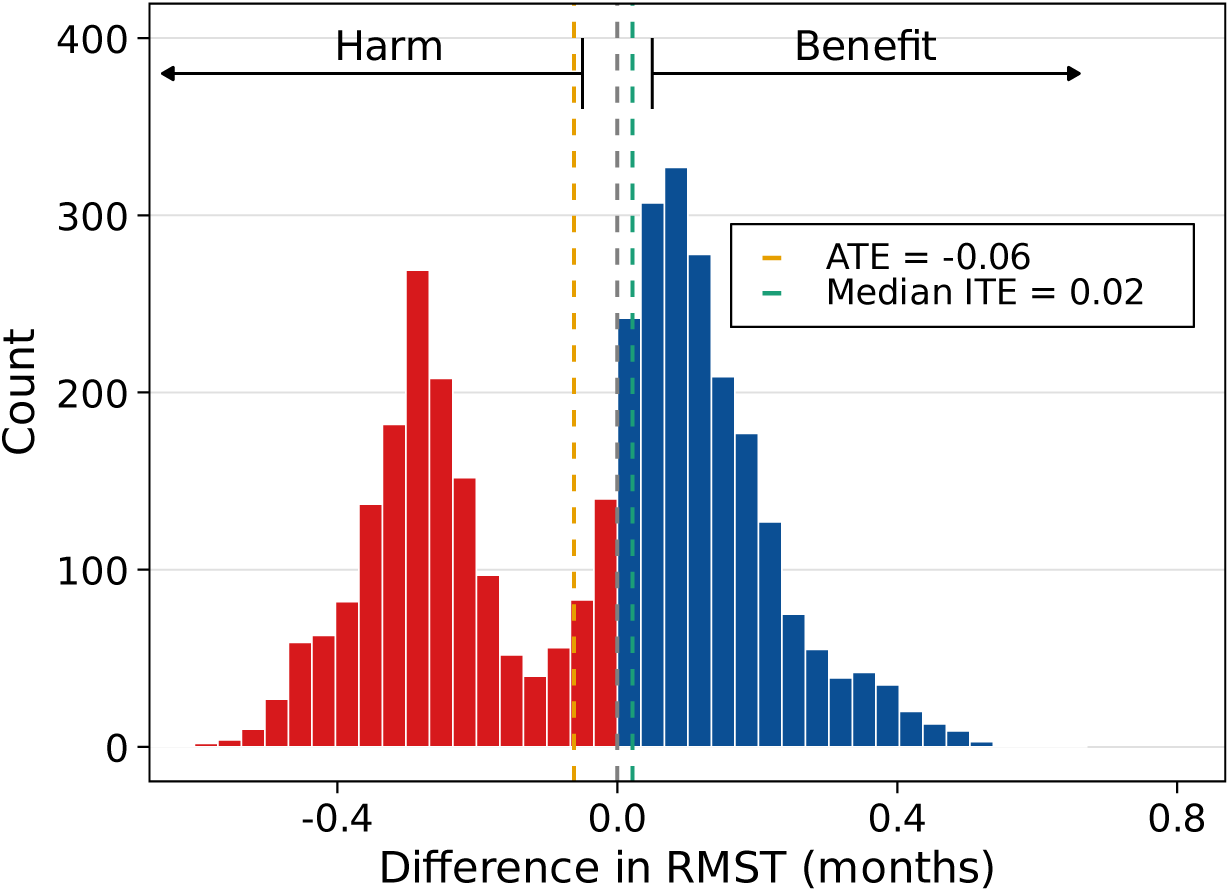
Distribution of individualized treatment effects for DPP-4 inhibitors versus GLP-1 receptor agonists. Individualized treatment effects (ITEs) were estimated by the causal survival forest and are expressed as differences in RMST at 48 months. Each bar represents the number of patients with a predicted RMST difference falling within the corresponding interval. Negative values indicate longer predicted clinical AD-type dementia-free survival under DPP-4 inhibitors, whereas positive values indicate longer predicted clinical AD-type dementia-free survival under GLP-1 receptor agonists. The dashed vertical lines indicate the augmented inverse probability-weighted (AIPW) average treatment effect and the median ITE.

### SHAP

**Supplementary Figure S8:**
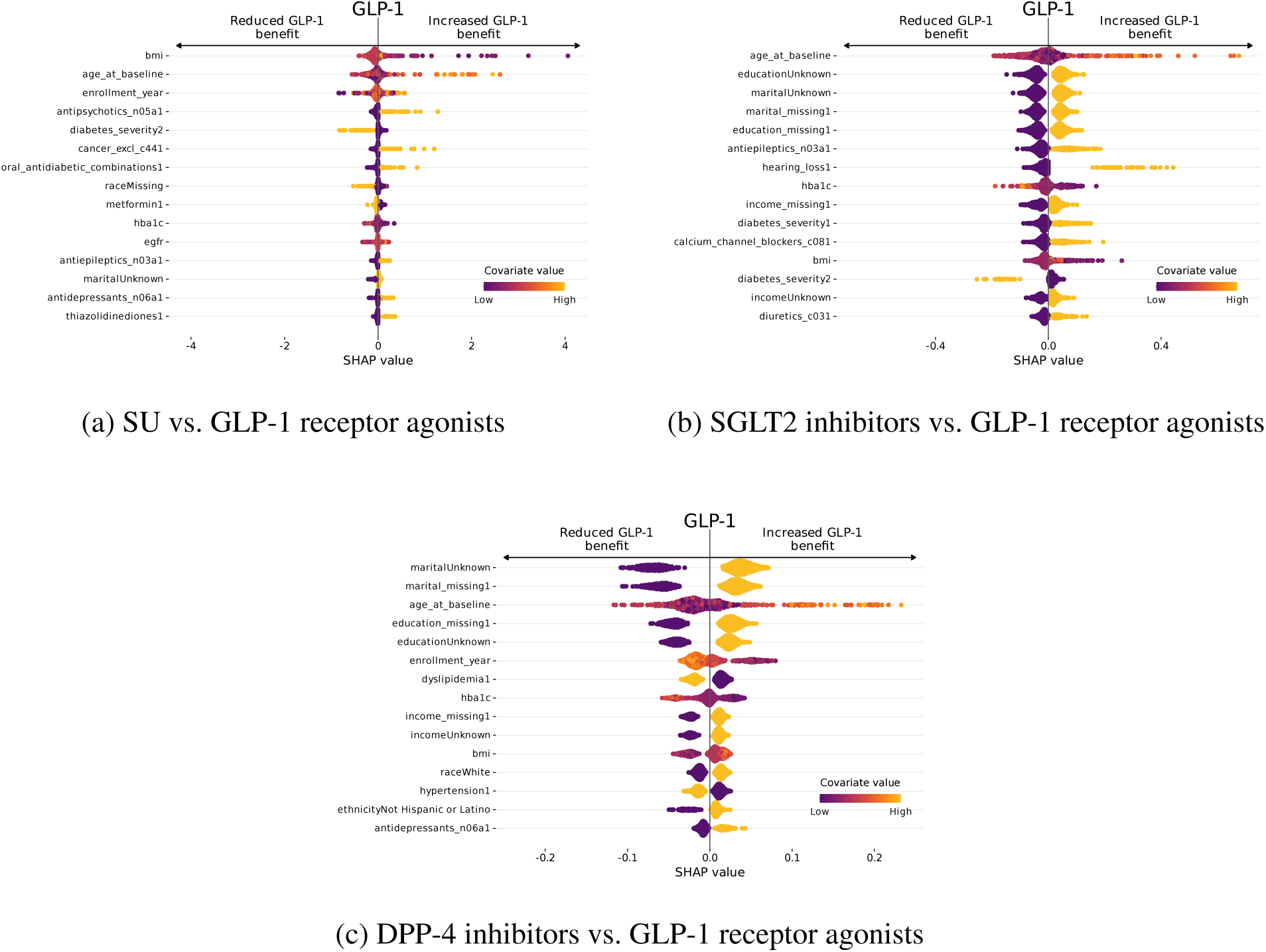
SHAP beeswarm plots for heterogeneous treatment effect modifiers. Each point represents one patient; the x-axis shows the SHAP value (contribution to the predicted individual treatment effect on RMST), and color indicates the covariate value (low = purple, high = yellow). Features are ranked by mean absolute SHAP value. Positive values indicate increased GLP-1 receptor agonist benefit relative to the comparator.^20^

### Propensity Score Diagnostics

**Supplementary Figure S9:**
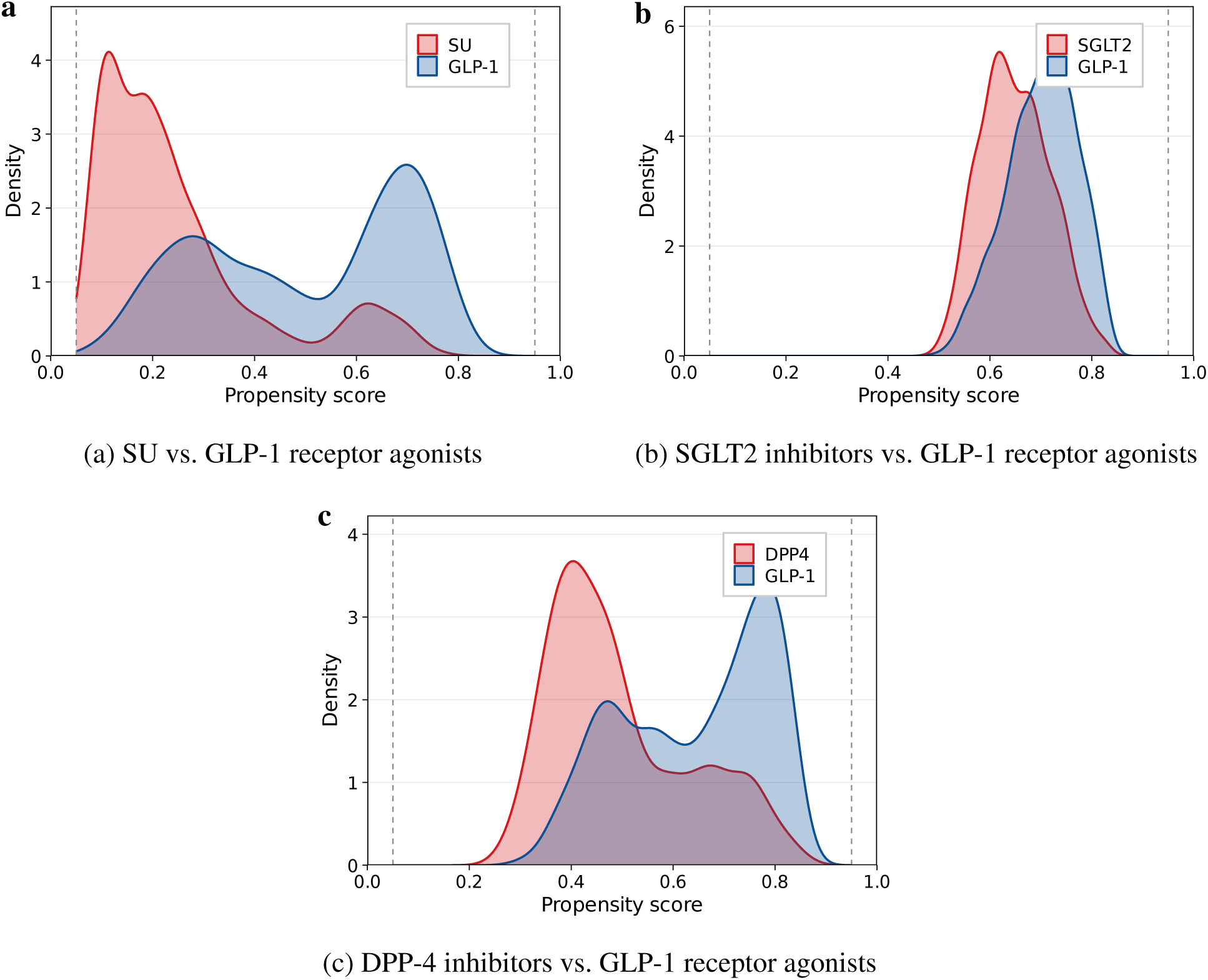
Propensity score overlap before IPTW weighting. Kernel density estimates of the estimated propensity score (probability of receiving GLP-1 receptor agonist treatment) for **(a)** SU versus GLP-1 receptor agonists, **(b)** SGLT2 inhibitors versus GLP-1 receptor agonists, and **(c)** DPP-4 inhibitors versus GLP-1 receptor agonists. Propensity scores were estimated using generalized random forests. Dashed vertical lines indicate the trimming thresholds at *P* = 0*·*05 and *P* = 0*·*95. Substantial overlap between treatment groups supports the positivity assumption required for valid causal inference.

### Propensity Score Calibration

**Supplementary Figure S10:**
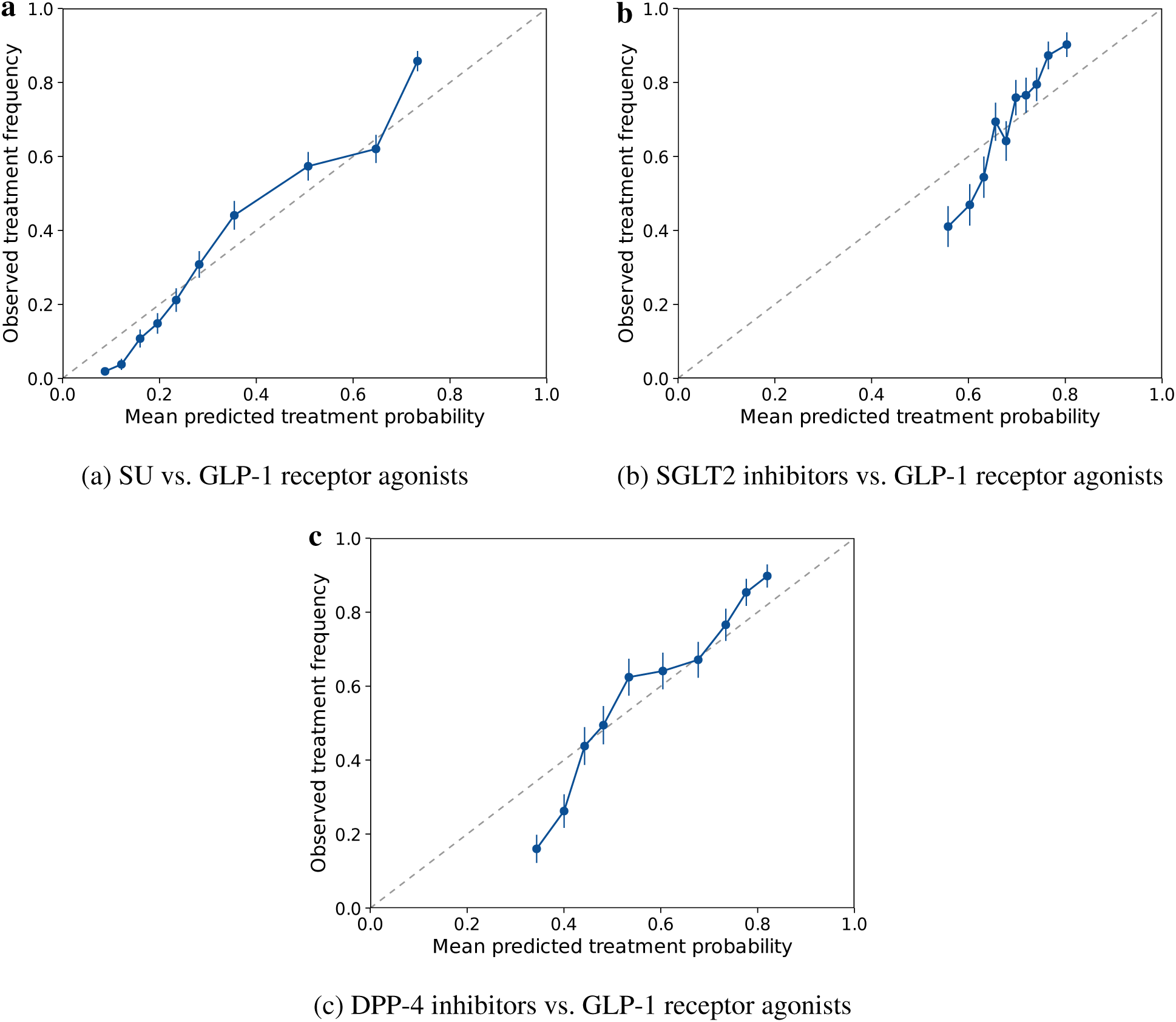
Propensity score calibration. Calibration plots of the estimated propensity scores for **(a)** SU versus GLP-1 receptor agonists, **(b)** SGLT2 inhibitors versus GLP-1 receptor agonists, and **(c)** DPP-4 inhibitors versus GLP-1 receptor agonists. Each point represents the mean predicted treatment probability against the observed treatment frequency within decile bins. Error bars denote 95% confidence intervals based on the binomial standard error. The dashed diagonal represents perfect calibration. Propensity scores were estimated using generalized random forests with 5-fold cross-fitting.

### Sensitivity Analyses with Alternative Models – Cox Regression

**Supplementary Figure S11:**
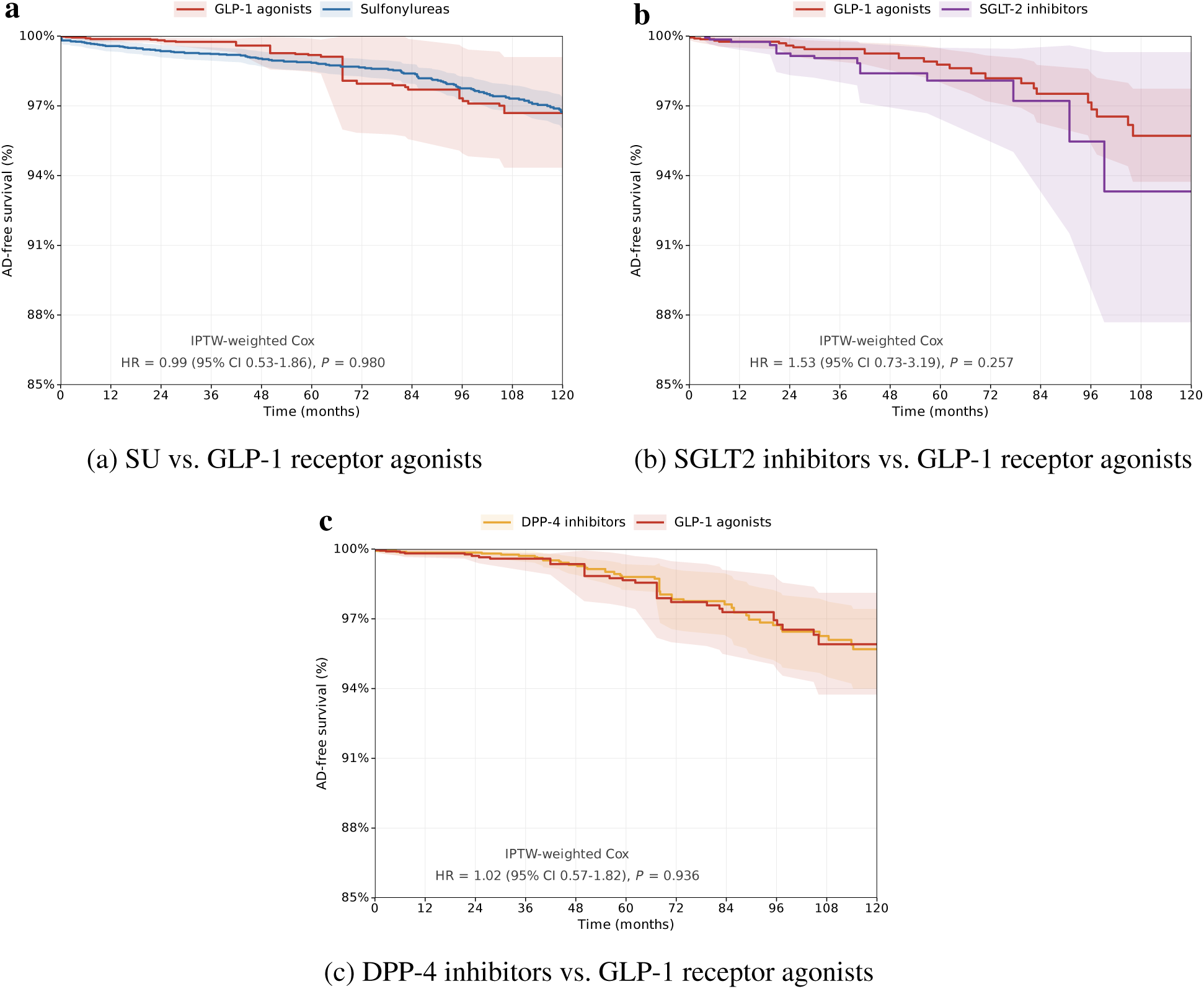
IPTW-weighted Kaplan–Meier survival curves for clinical AD-type dementia-free survival. Curves show the probability of remaining clinical AD-type dementia-free over 120 months of follow-up for **(a)** SU versus GLP-1 receptor agonists, **(b)** SGLT2 inhibitors versus GLP-1 receptor agonists, and **(c)** DPP-4 inhibitors versus GLP-1 receptor agonists. Shaded bands represent 95% confidence intervals. Weights were derived from stabilized inverse probability of treatment weighting (IPTW) based on propensity scores estimated via generalized random forests. Hazard ratios (HRs) with 95% CIs and Wald *P* values were obtained from IPTW-weighted Cox proportional hazards models with robust standard errors.

### Sensitivity Analyses with Alternative Models – T-Learner

**Supplementary Figure S12:**
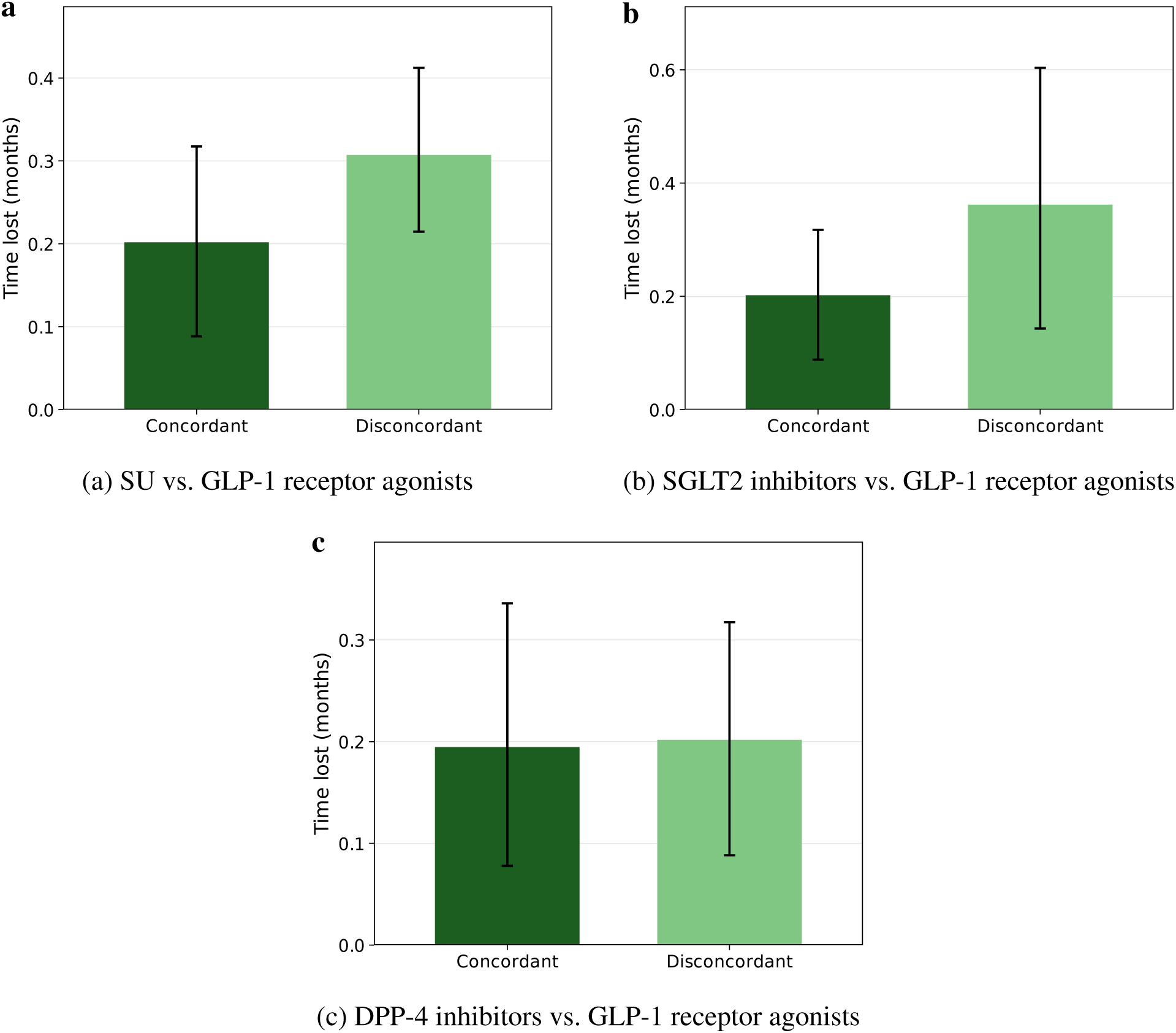
Time lost to clinical AD-type dementia in concordant and discordant patient pairs in the T-learner sensitivity analysis. Distribution of clinical AD-type dementia-free time lost in months, stratified by concordance status, for **(a)** SU versus GLP-1 receptor agonists, **(b)** SGLT2 inhibitors versus GLP-1 receptor agonists, and **(c)** DPP-4 inhibitors versus GLP-1 receptor agonists. Individualized treatment effects were estimated using the Tlearner model. Concordant pairs were defined as pairs in which the individual with the higher predicted benefit from GLP-1 receptor agonist initiation experienced the longer clinical AD-type dementia-free survival time. Discordant pairs were defined as pairs in which the individual with the lower predicted benefit experienced the longer clinical AD-type dementia-free survival time. Boxes show the interquartile range and median; violin shapes represent the full distribution. Differences between groups were assessed using the Wilcoxon rank-sum test. Results were consistent with the primary causal survival forest estimates, supporting the robustness of heterogeneous treatment effect findings.

### Subgroup definitions

**Supplementary Table S5:** Definitions of subgroup categories used in subgroup treatment effect analyses.

| Subgroup variable | Categories | Definition |
| --- | --- | --- |
| Age | <65, 65–70, 70–75, 75–80, $\geq 80$ years | Based on age at baseline |
| Sex | Female, Male | Based on recorded sex at birth |
| BMI category | <25, 25–30, 30–35, $\geq 35$ kg/m<sup>2</sup> | Based on baseline BMI |
| Body weight, SU vs GLP-1RA | Low, Mid, High | Low: <88.0 kg; Mid: 88.0–<106.6 kg; High: $\geq 106.6$ kg |
| Body weight, SGLT2i vs GLP-1RA | Low, Mid, High | Low: <88.9 kg; Mid: 88.9–<107.0 kg; High: $\geq 107.0$ kg |
| Body weight, DPP-4i vs GLP-1RA | Low, Mid, High | Low: <86.4 kg; Mid: 86.4–<104.8 kg; High: $\geq 104.8$ kg |
| Obesity | Non-obese, obese | Based on recorded obesity diagnosis or obesity indicator |
| HbA1c at baseline | <7%, 7–8%, 8–9%, $\geq 9\%$ | Based on baseline HbA1c |
| Hypertension | No, Yes | Based on recorded hypertension diagnosis |
| Metformin use | No, Yes | Based on baseline medication history |
| Insulin use | No, Yes | Based on baseline medication history |

### Co-Medication: Concomitant Drug Use During Follow-up

**Supplementary Figure S13:**
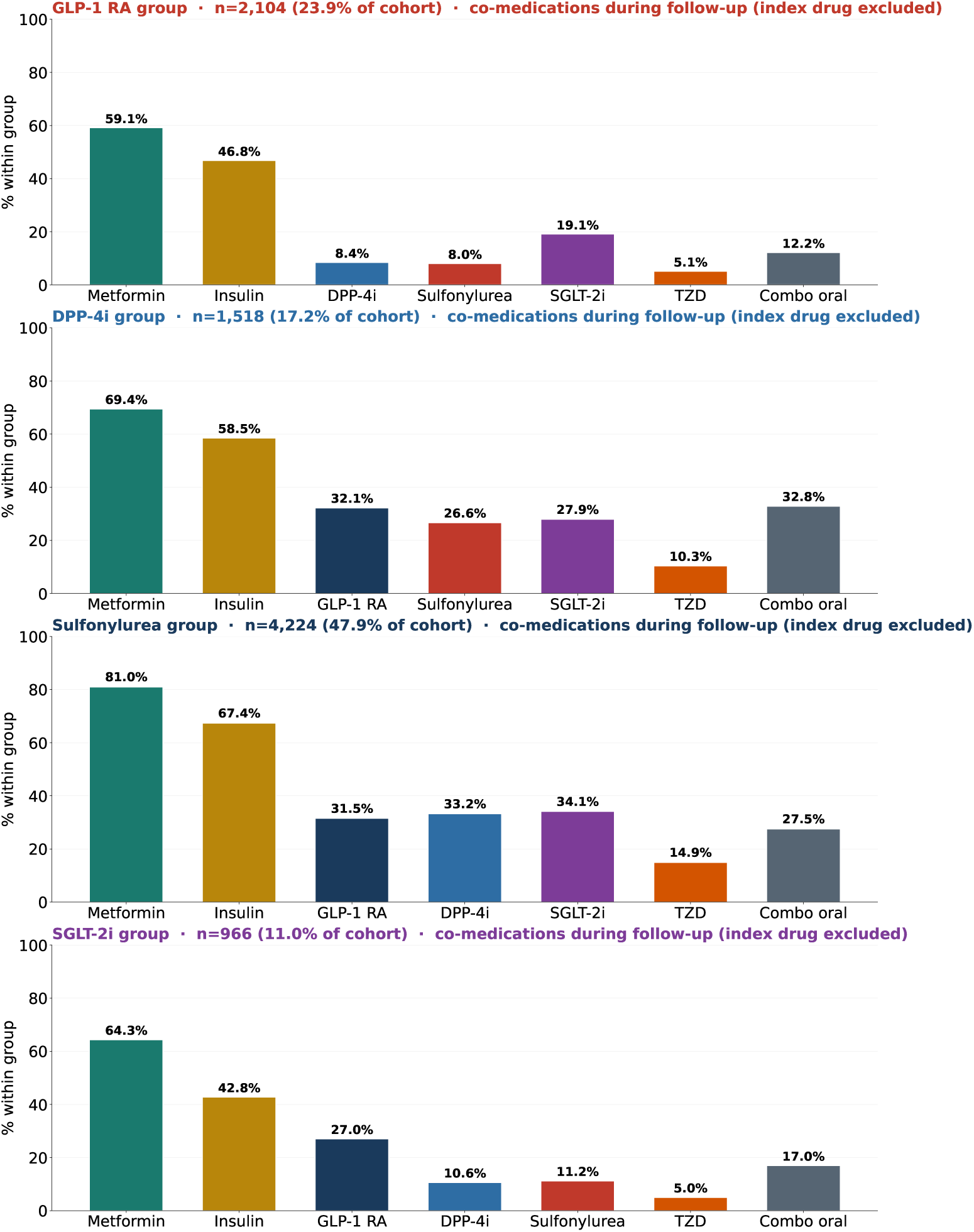
Concomitant antidiabetic drug use during follow-up by treatment group. Bars show the proportion of patients within each treatment group receiving concomitant antidiabetic drug classes at any point during follow-up. The index drug class is excluded from each panel to reflect co-medication rather than continuation. Proportions are calculated within treatment group. “Other oral” antidiabetics are excluded due to heterogeneous ATC mapping. GLP-1 RA: GLP-1 receptor agonists; DPP-4 inhibitors: dipeptidyl peptidase-4 inhibitors; SU: sulfonylureas; SGLT2i: SGLT2 inhibitors; TZD: thiazolidinediones. *n* = 8,812.

### 5-year causal machine learning estimates

**Supplementary Figure S14:**
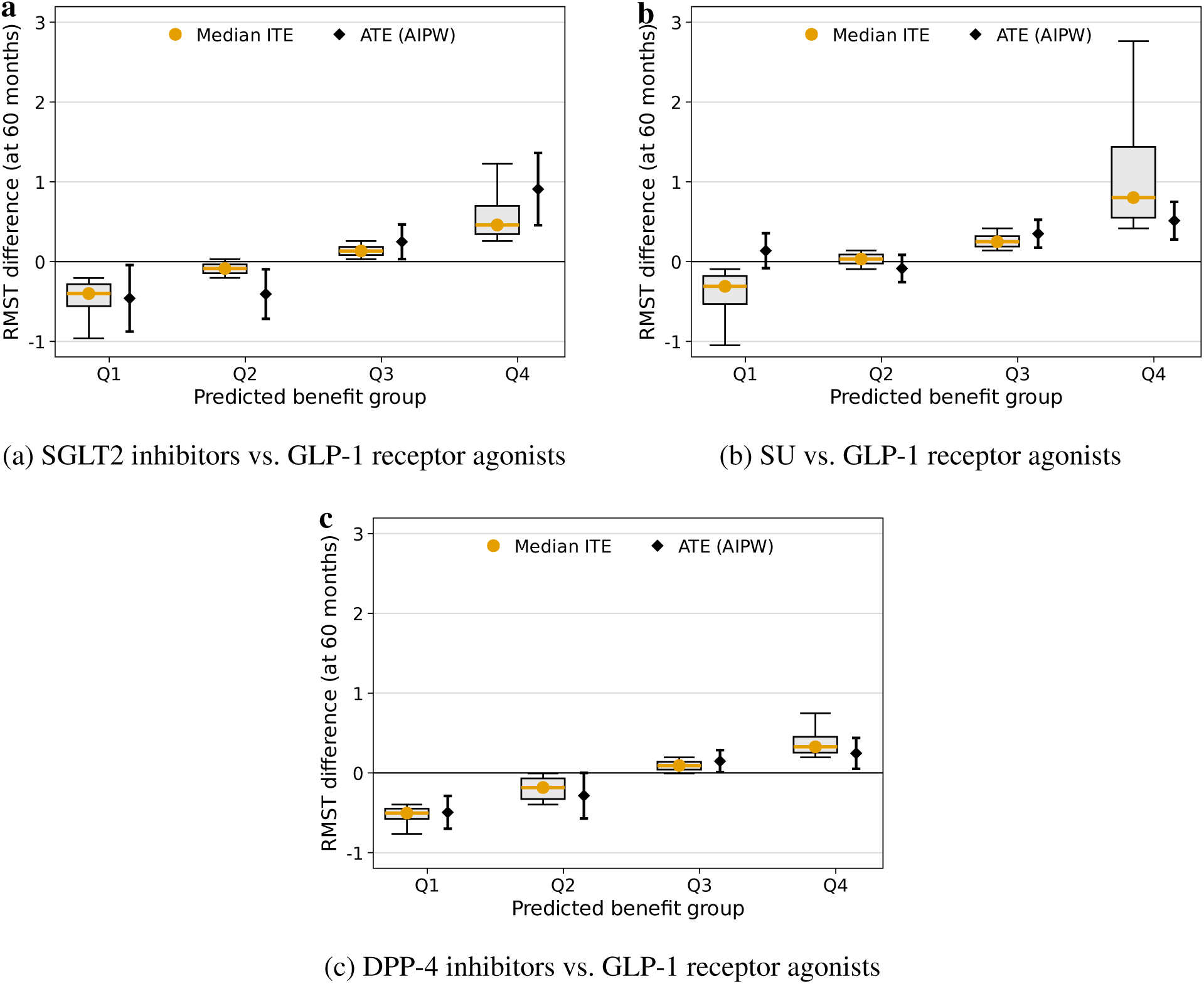
Causal machine learning estimates of treatment effect heterogeneity at 5 years. RMST differences at 60 months are shown across predicted benefit groups (Q1–Q4) for **(a)** GLP-1 receptor agonists versus SGLT2 inhibitors, **(b)** GLP-1 receptor agonists versus SU, and **(c)** GLP-1 receptor agonists versus DPP-4 inhibitors. Values are expressed as GLP-1 receptor agonist initiation minus comparator initiation. Positive values therefore indicate longer predicted clinical AD-type dementia-free survival under GLP-1 receptor agonists, whereas negative values indicate longer predicted clinical AD-type dementia-free survival under the comparator. Points represent median individualized treatment effects, and diamonds represent average treatment effects estimated using augmented inverse probability weighting (AIPW).

### Target trial timing

**Supplementary Figure S15:**
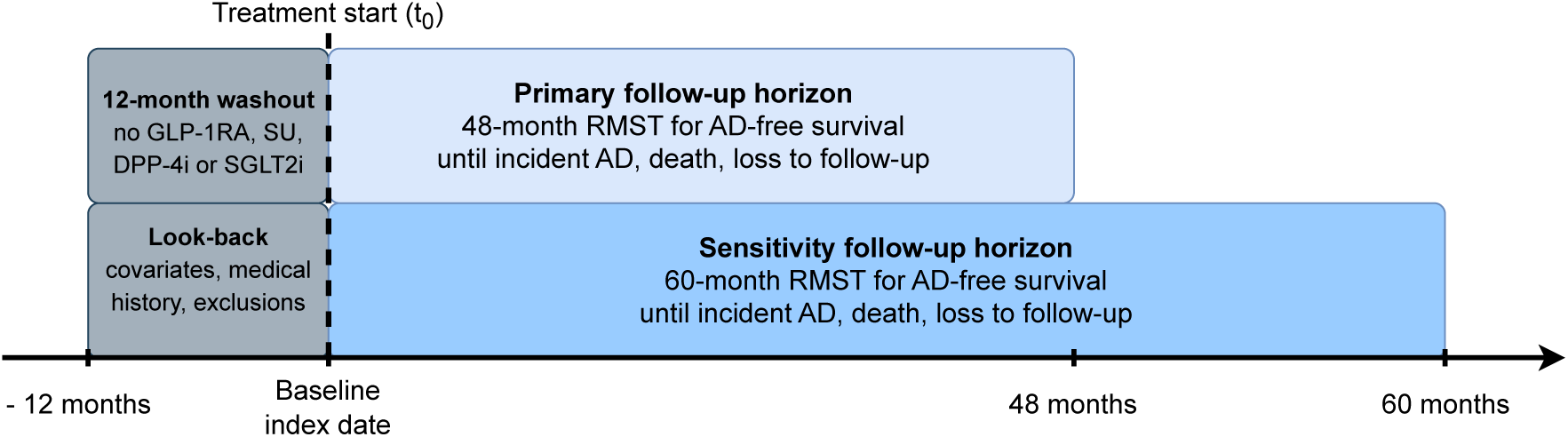
Target trial timing, look-back, washout, and follow-up horizons. Schematic overview of the temporal design used for the active-comparator, new-user target trial emulations. Baseline was defined as the date of first qualifying treatment initiation (*t*_0_). A fixed 12-month look-back period before baseline was used to assess baseline covariates, medical history, and exclusion criteria. The same 12-month pre-baseline window was used as a treatment-free washout period, requiring no recorded exposure to GLP-1 receptor agonists, SU, DPP-4 inhibitors, or SGLT2 inhibitors before index. Follow-up started at baseline and continued until incident Alzheimer’s disease, death, loss to follow-up, or the end of the prespecified horizon. The primary estimand was the difference in clinical AD-type dementia-free restricted mean survival time over 48 months, with an additional 60-month horizon used as a sensitivity analysis.

### Sensitivity analysis including cardiovascular and cerebrovascular disease

#### Rationale and analytic modification

Because exclusion of patients with cardiovascular or cerebrovascular disease may remove a subgroup with a high baseline risk and potentially high absolute benefit from GLP-1 receptor agonist treatment, and may select a healthier appearing treated cohort, we repeated the analyses without this exclusion. The sensitivity cohort therefore retained patients with baseline ischemic heart disease, heart failure, atrial fibrillation, aortic aneurysm or dissection, and cerebrovascular disease. The primary models were adjusted for these five component indicators. A composite indicator of cardiovascular or cerebrovascular history was used only for effect modification summaries and was not entered together with its component indicators, thereby avoiding deterministic collinearity. The new-user washout, baseline dementia exclusions, endpoint definitions, 48-month estimand, and the remainder of the modeling pipeline were unchanged.

#### Average treatment effects

Table S6 summarizes the 48-month RMST differences, expressed as GLP-1 receptor agonist initiation minus comparator initiation. Retaining patients with cardiovascular and cerebrovascular disease did not provide evidence of a statistically significant average benefit for any of the three active-comparator contrasts. The estimated average treatment effect was *−*0*·*06 months for DPP-4 inhibitors versus GLP-1 receptor agonists (95% CI, *−*0*·*25 to 0*·*14; *P* = 0*·*564), 0*·*03 months for SGLT2 inhibitors versus GLP-1 receptor agonists (95% CI, *−*0*·*21 to 0*·*27; *P* = 0*·*816), and 0*·*13 months for SU versus GLP-1 receptor agonists (95% CI, *−*0*·*01 to 0*·*28; *P* = 0*·*074).

**Supplementary Table S6:** Average treatment effects after inclusion of cardiovascular and cerebrovascular disease.

| Comparator vs. GLP-1 RA | Analysis $n$ | ATE, months | 95% CI, months | $P$ value |
| --- | --- | --- | --- | --- |
| DPP-4 inhibitor | 4,761 | $-0.057$ | $-0.252$ to $0.137$ | **0.564** |
| SGLT2 inhibitor | 4,699 | $0.029$ | $-0.212$ to $0.270$ | **0.816** |
| SU | 7,811 | $0.135$ | $-0.013$ to $0.282$ | **0.074** |

ATE denotes the augmented inverse probability-weighted average treatment effect on 48-month dementia-free RMST. Positive values favor GLP-1 receptor agonists.

#### Treatment effect heterogeneity

The distribution of predicted individualized treatment effects is shown in Supplementary Figure S16. For the SGLT2 inhibitor and SU contrasts, median predicted effects increased across predicted-benefit quartiles, with Q4–Q1 differences of approximately 0*·*93 and 0*·*82 months, respectively. DPP-4 inhibitor quartile medians remained tightly clustered near zero (Q4–Q1 difference, approximately 0*·*01 months). In the DPP-4 inhibitor comparison, quartile-specific AIPW estimates were unstable and did not follow the ordering of the predicted individualized effects; these subgroup estimates should therefore be interpreted cautiously. Formal linear trend tests were not statistically significant (DPP-4 inhibitor, *P* = 0*·*602; SGLT2 inhibitor, *P* = 0*·*275; SU, *P* = 0*·*377).

**Supplementary Figure S16:**
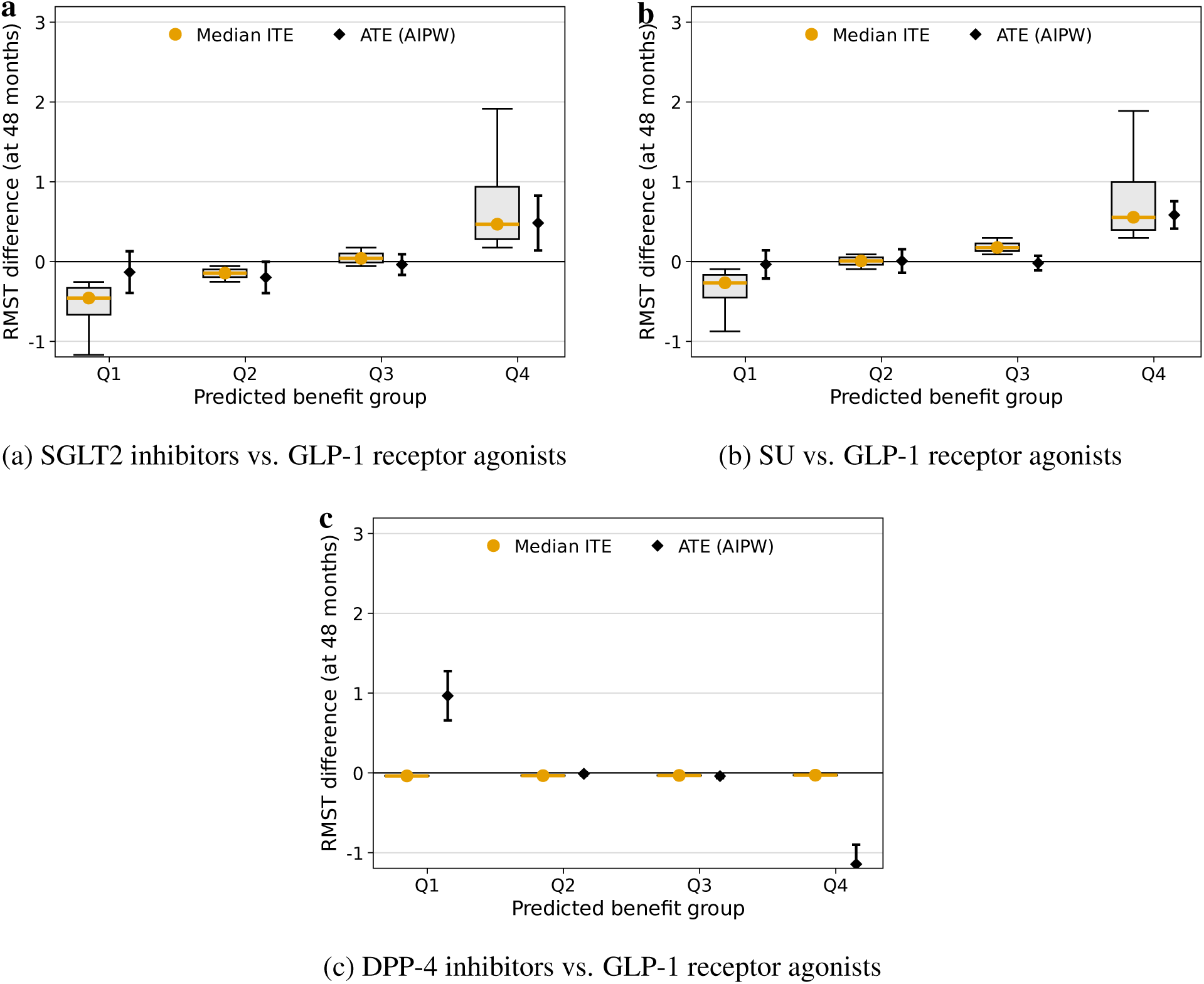
Predicted treatment effect heterogeneity after inclusion of cardiovascular and cerebrovascular disease. Predicted-benefit quartiles (Q1–Q4) are shown for **(a)** SGLT2 inhibitors, **(b)** SU, and **(c)** DPP-4 inhibitors, each compared with GLP-1 receptor agonists. Values are GLP-1 receptor agonist initiation minus comparator initiation on the 48-month dementia-free RMST scale. Orange points show median individualized treatment effects and black diamonds show AIPW average treatment effects. Small-cell counts are not displayed.

#### Concordance–discordance validation

Observed dementia-free time loss was similar between participants whose observed treatment was concordant and discordant with the model-predicted treatment benefit (Supplementary Figure S17). The corresponding concordance-minus-discordance RMST differences were small and not statistically significant: 0*·*02 months for DPP-4 inhibitors (95% CI, *−*0*·*18 to 0*·*20; *P* = 0*·*853), 0*·*11 months for SGLT2 inhibitors (95% CI, *−*0*·*12 to 0*·*37; *P* = 0*·*327), and 0*·*08 months for SU (95% CI, *−*0*·*08 to 0*·*25; *P* = 0*·*247; Supplementary Figure S18). These results do not support a clear improvement in observed 48-month dementia-free RMST when treatment assignment aligned with the predicted individualized benefit.

**Supplementary Figure S17:**
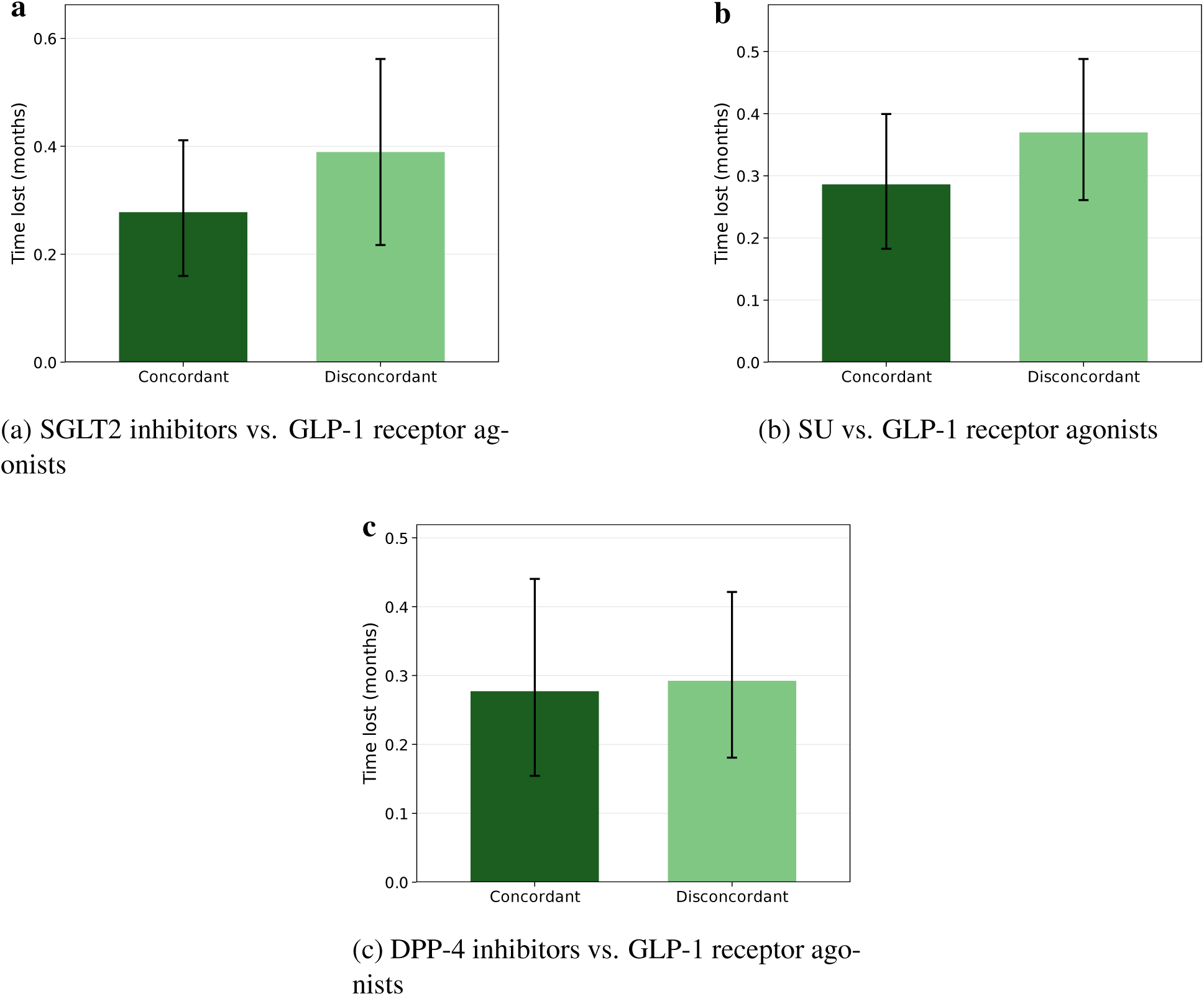
Dementia-free time loss by treatment concordance after inclusion of cardiovascular and cerebrovascular disease. Weighted mean dementia-free time loss through 48 months is shown for concordant and discordant treatment assignments for **(a)** SGLT2 inhibitors, **(b)** SU, and **(c)** DPP-4 inhibitors, each compared with GLP-1 receptor agonists. Error bars denote 95% confidence intervals. Only aggregate groups with at least 20 participants are displayed.

**Supplementary Figure S18:**
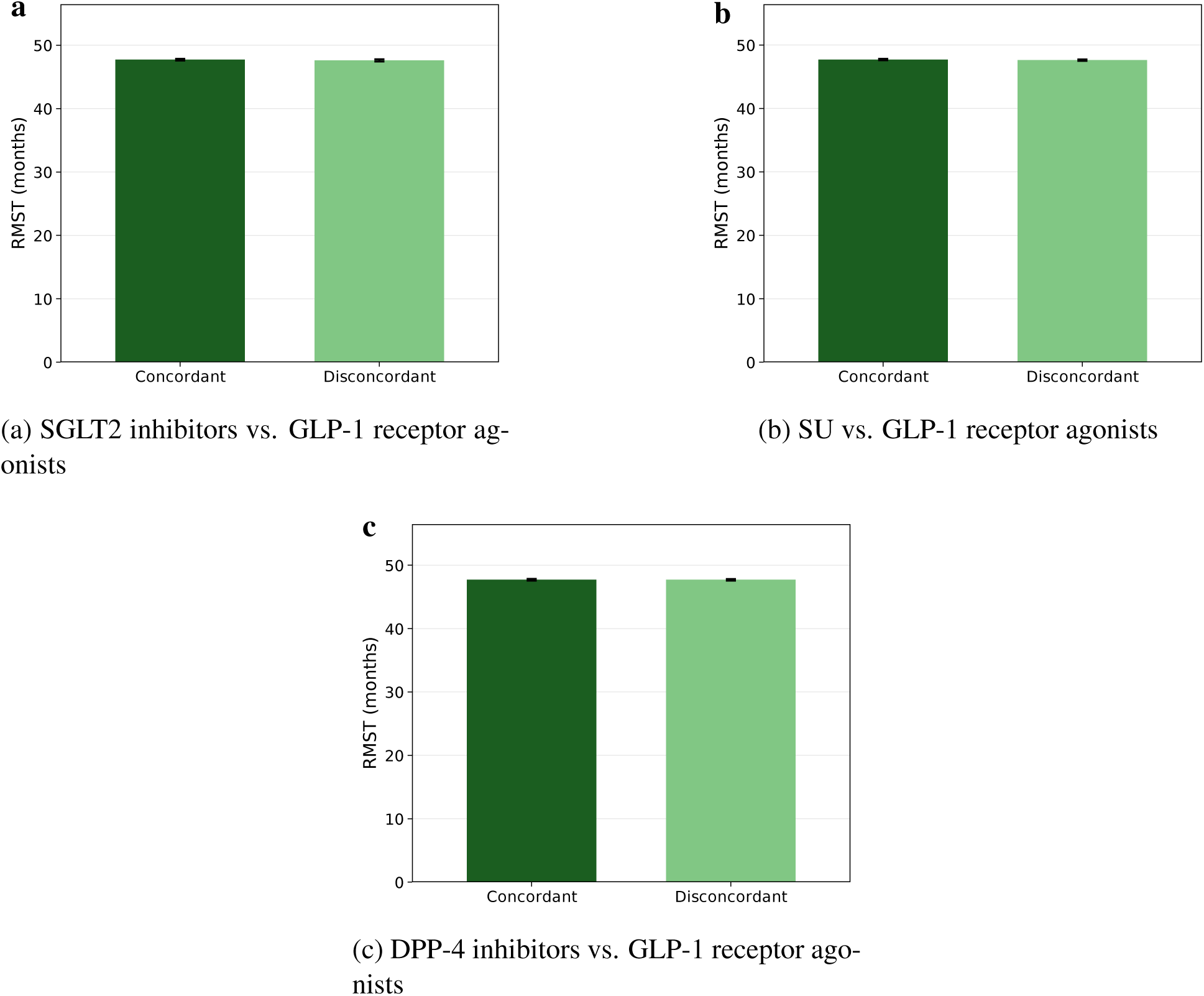
Dementia-free RMST by treatment concordance after inclusion of cardiovascular and cerebrovascular disease. Weighted 48-month dementia-free RMST is shown for concordant and discordant treatment assignments for **(a)** SGLT2 inhibitors, **(b)** SU, and **(c)** DPP-4 inhibitors, each compared with GLP-1 receptor agonists. Error bars denote 95% confidence intervals. Only aggregate groups with at least 20 participants are displayed.

## **A** Supplementary Discussion

### **A.1** Additional clinical context and interpretation of absolute effects

Trials in patients with established symptomatic AD address a further, clinically different question. In ELAD, liraglutide did not significantly improve the primary cerebral glucose-metabolism endpoint, although secondary cognitive and imaging signals were reported.^21^ More recently, the evoke and evoke+ phase 3 trials did not show that oral semaglutide slowed clinical progression in early symptomatic AD despite changes in some disease-related biomarkers.^22^ These trial results do not directly refute a possible association with delayed onset in people with diabetes who do not yet have clinically recorded dementia, because prevention and symptomatic treatment occur at different disease stages. Nevertheless, they temper mechanistic interpretation and indicate that the observational associations reported here should not be presented as established disease modification. Variation among clinical studies can also reflect differences in design, comparator, follow-up, outcome definition, and study population.^23^ The long preclinical development of AD means that the 48-month primary horizon may capture only a limited part of any treatment-related difference, even though the 60-month analysis produced broadly consistent patterns.

The absolute magnitude of the population-level association against SU was small. An RMST difference of 0*·*21 months over 48 months corresponds to only several additional days without a clinically recorded AD-type dementia event for the average patient. This estimate is more appropriately viewed as a population-level signal than as a clinically meaningful gain for an individual. Even the larger estimate in the highest predicted-benefit stratum remains modest and uncertain in relation to the long course of AD. Therefore, neither comparison supports selecting GLP-1 receptor agonists solely to reduce dementia risk. Treatment choice should continue to be guided primarily by glycemic control, weight management, cardiovascular and renal indications, heart failure, tolerability, contraindications, access, and patient preferences.^24, 25^

### **A.2** Treatment effect heterogeneity and exploratory subgroups

The heterogeneity analyses also require cautious interpretation. Causal survival forests are useful for detecting and ranking treatment effect variation without imposing a prespecified functional form, and the increasing stratum-level estimates indicate that the effect scores enriched groups with larger estimated average benefit. However, this does not imply that the individualized treatment effect is known precisely for any single patient. Only one potential outcome is observed for each individual, and the reported confidence intervals quantify average effects within model-defined strata rather than uncertainty around each patient’s ITE. Given the small number of dementia events, patient-specific treatment effect intervals would be expected to be wide. The C-for-benefit values of 0*·*578 in the SU comparison and 0*·*637 in the SGLT2 inhibitor comparison indicate modest ranking discrimination rather than accurate individual prediction. Consequently, the model should be understood as providing group-level prioritization for further study, not as a clinical treatment rule or a tool for choosing between GLP-1 receptor agonists and SGLT2 inhibitors for an individual patient.

The conventional subgroup analyses reinforce this caution. Patterns across age, BMI, and HbA1c were not consistent between comparisons and did not form a coherent biological gradient. For example, lower BMI was associated with a comparatively large positive estimate for GLP-1 receptor agonists versus SU, but the lowest BMI category favored SGLT2 inhibitors in the contemporary comparison. Likewise, the largest estimate across HbA1c categories occurred at 7–8% in the SU comparison, whereas the numerically largest estimate in the SGLT2 inhibitor comparison occurred below 7%. Age-related estimates also changed direction and were particularly imprecise in the oldest and smallest strata. Some variation is expected because effect modification is comparator-specific, but the observed patterns do not provide a sufficiently stable or biologically plausible basis for interpretation. They may reflect chance, sparse events, residual confounding, multiple exploratory comparisons, or interactions that were not reproducible across contrasts. We therefore do not interpret individual age, BMI, or HbA1c subgroup findings as evidence for specific treatment phenotypes. Their appropriate role is to generate hypotheses for prespecified testing in independent data.

### **A.3** Study strengths

Our study has several strengths. First, we integrated target trial emulation^1^ with causal machine learning^6^ to estimate clinically relevant active-comparator effects while exploring heterogeneity in time-to-event outcomes. Second, the active-comparator, new-user design aligned eligibility, treatment initiation, and follow-up and reduced biases related to prevalent use, immortal time, and comparisons with untreated individuals. Third, longitudinal data from the *All of Us Research Program* provided harmonized information on diagnoses, medications, laboratory measurements, and socioeconomic characteristics in a diverse real-world cohort. The inclusion of SGLT2 inhibitors allowed evaluation against a modern comparator used in overlapping clinical contexts. Finally, the analysis included cross-fitting, doubly robust estimation, overlap and calibration diagnostics, a negative-control outcome, longer horizon analyses, and an alternative T-learner. These analyses supported the presence of group-level ranking information, while not eliminating the uncertainty inherent in observational individualized-effect estimation.

### **A.4** Additional sources of residual confounding and outcome limitations

Cognitive reserve, baseline cognition, and educational quality were also incompletely measured. Although education was available as a covariate, it was missing for a large proportion of participants, especially among SU initiators, and EHR data do not capture lifelong cognitive reserve, literacy, occupational complexity, or subtle cognitive impairment before diagnosis. These factors could influence both medication selection and the probability or timing of a dementia diagnosis. Healthcare engagement represents a related source of potential healthy-user bias. Patients willing and able to initiate and continue an injectable medication may differ in adherence, diet, exercise, self-management, socioeconomic resources, caregiver support, and use of specialist care. Greater engagement could lower underlying dementia risk through pathways unrelated to the drug. At the same time, patients receiving newer therapies may have more frequent clinical contact, increasing opportunities to detect cognitive symptoms and potentially biasing the association toward earlier dementia recording. These mechanisms act in opposing directions and cannot be resolved fully with the available data.

The outcome itself was based on routinely recorded diagnoses and anti-dementia medication dispensing rather than biomarker confirmation, standardized neurological adjudication, or serial neuropsychological assessment. It may therefore include mixed, vascular, or other dementia syndromes and may miss undiagnosed disease. Differential coding, outside-system care, and incomplete medication capture may have caused outcome or exposure misclassification. The *All of Us* cohort is also volunteer-based, and treatment access and recording patterns may not generalize to all healthcare systems or to populations with larger frailty, lower healthcare access, or different racial and socioeconomic composition.

### **A.5** Additional methodological and mechanistic limitations

Methodological limitations further constrain interpretation. Treatment assignment followed an intention-to-treat-like strategy at initiation; discontinuation, adherence, switching, treatment intensification, and addition of other glucose-lowering drugs were not modeled as time-varying exposures. Such changes were common during follow-up and could attenuate or alter the estimated contrasts. The 48-month horizon is short relative to the preclinical course of AD, and event counts were limited, particularly in smaller subgroups and in the SGLT2 inhibitor comparison. The many subgroup estimates were exploratory and were not intended as confirmatory interaction tests. Negative-control and alternative-model analyses reduce concern about some forms of bias but cannot exclude unmeasured confounding or validate patient-specific causal effects. External validation is therefore essential, ideally in datasets with adjudicated outcomes, more complete measures of frailty and cognition, longer follow-up, and sufficient events to quantify uncertainty in treatment effect rankings.

The observational design does not establish the biological mechanism underlying the association. Clinical AD-type dementia has a multifactorial aetiology,^26^ and direct effects of GLP-1 receptor agonists on inflammatory, vascular, metabolic, or neuroprotective pathways remain biologically plausible.^27–29^ Indirect effects through cardiometabolic or weight-related pathways are also possible.^30, 31^ However, the absence of alignment between weight loss and the dementia-benefit strata suggests that short-term weight reduction is not the principal explanation for the estimated heterogeneity. Future studies should formally examine mediators using longitudinal metabolic, vascular, inflammatory, and neurodegenerative biomarkers rather than inferring mechanisms from baseline subgroup patterns. Replication of the benefit ranking in independent healthcare systems and prospective evaluation of prespecified treatment policies are needed before any clinical translation.

## STROBE Statement—checklist of items that should be included in reports of observational studies

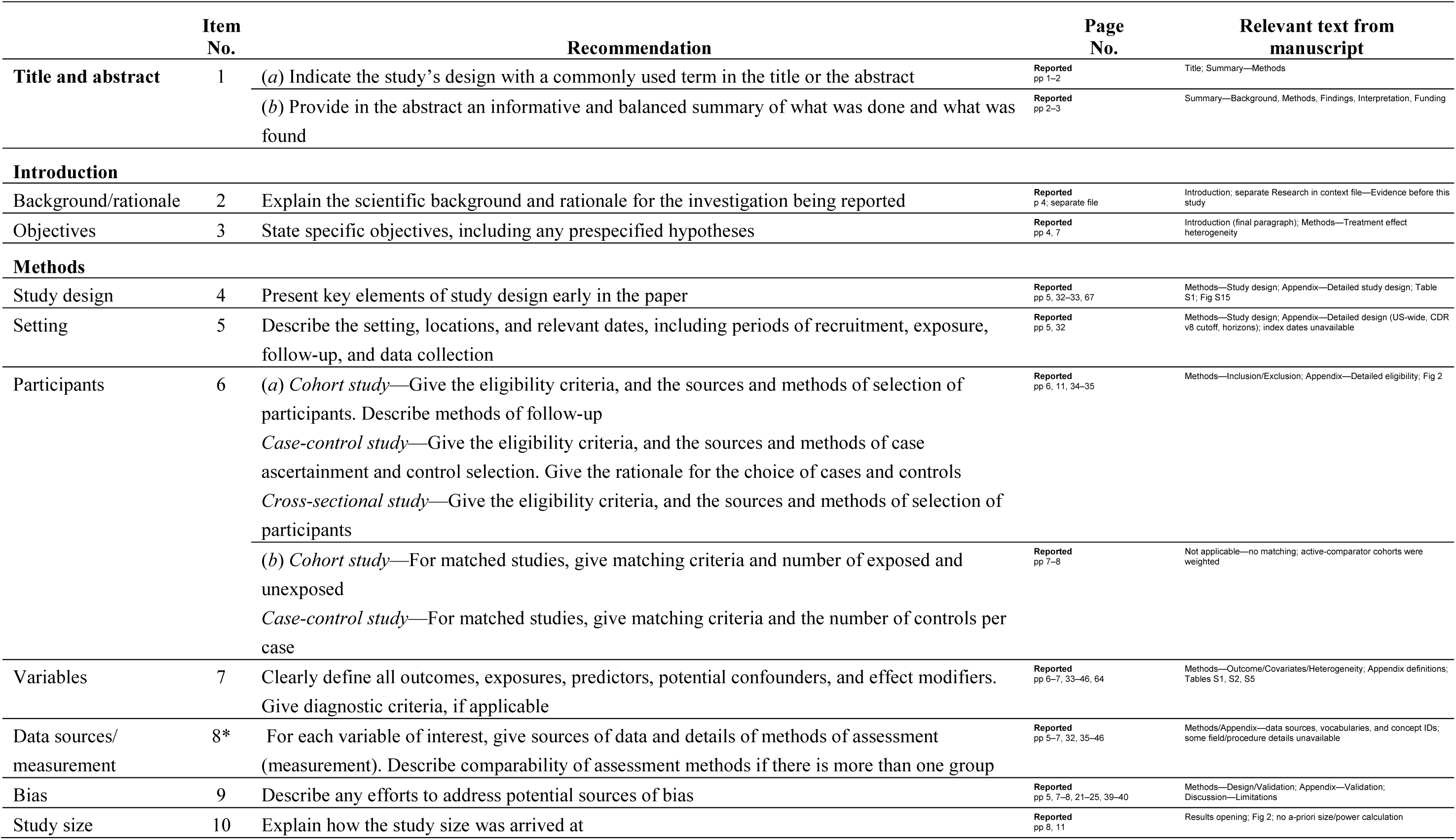
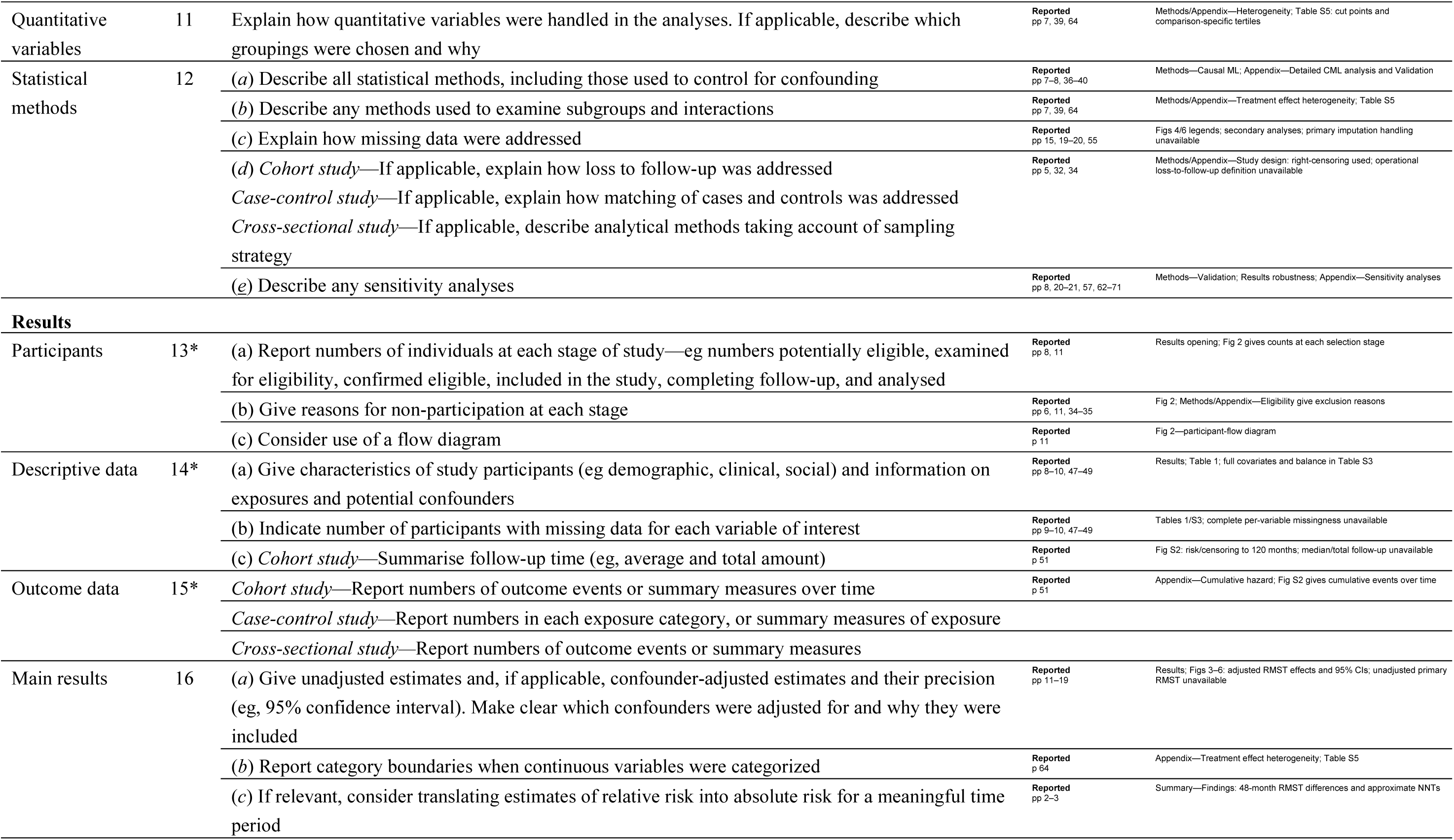
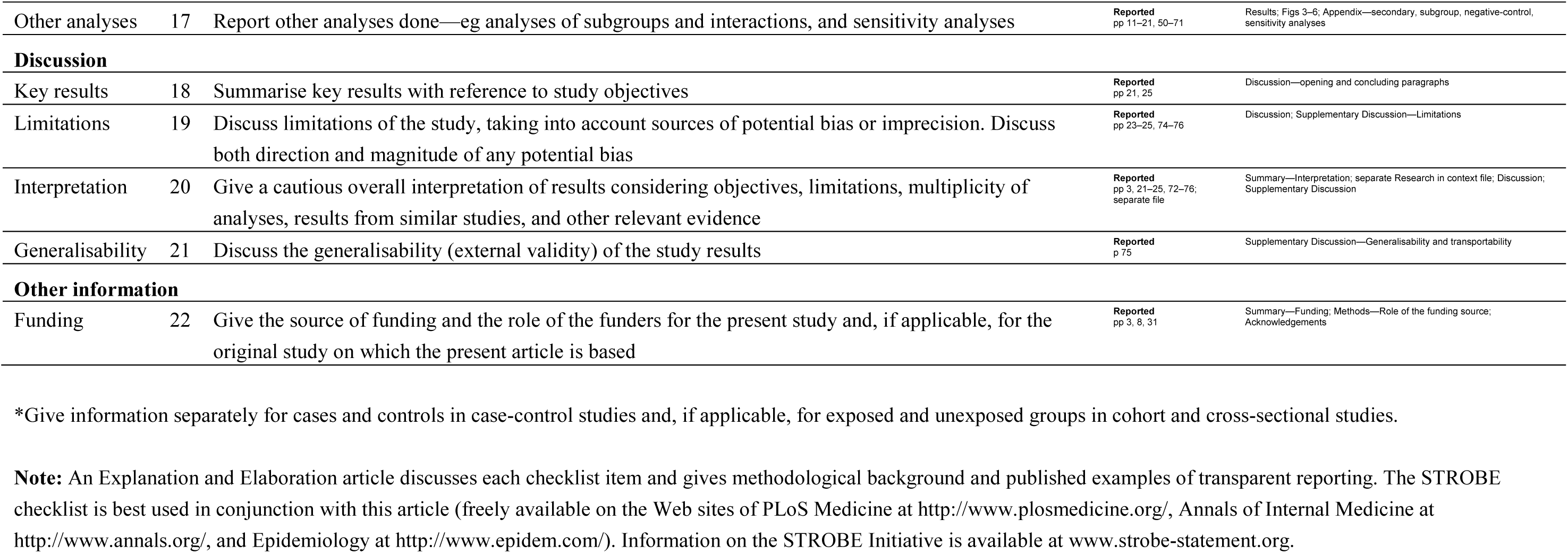

|  | Item No. | Recommendation | Page No. | Relevant text from manuscript |
| --- | --- | --- | --- | --- |
| Title and abstract | 1 | (a) Indicate the study’s design with a commonly used term in the title or the abstract | Reported pp 1–2 | Title; Summary—Methods |
|  |  | (b) Provide in the abstract an informative and balanced summary of what was done and what was found | Reported pp 2–3 | Summary—Background, Methods, Findings, Interpretation, Funding |
| Introduction |  |  |  |  |
| Background/rationale | 2 | Explain the scientific background and rationale for the investigation being reported | Reported p 4; separate file | Introduction; separate Research in context file—Evidence before this study |
| Objectives | 3 | State specific objectives, including any prespecified hypotheses | Reported pp 4, 7 | Introduction (final paragraph); Methods—Treatment effect heterogeneity |
| Methods |  |  |  |  |
| Study design | 4 | Present key elements of study design early in the paper | Reported pp 5, 32–33, 67 | Methods—Study design; Appendix—Detailed study design; Table S1; Fig S15 |
| Setting | 5 | Describe the setting, locations, and relevant dates, including periods of recruitment, exposure, follow-up, and data collection | Reported pp 5, 32 | Methods—Study design; Appendix—Detailed design (US-wide, CDR v8 cutoff, horizons); index dates unavailable |
| Participants | 6 | (a) Cohort study—Give the eligibility criteria, and the sources and methods of selection of participants. Describe methods of follow-up<br>Case-control study—Give the eligibility criteria, and the sources and methods of case ascertainment and control selection. Give the rationale for the choice of cases and controls<br>Cross-sectional study—Give the eligibility criteria, and the sources and methods of selection of participants | Reported pp 6, 11, 34–35 | Methods—Inclusion/Exclusion; Appendix—Detailed eligibility; Fig 2 |
|  |  | (b) Cohort study—For matched studies, give matching criteria and number of exposed and unexposed<br>Case-control study—For matched studies, give matching criteria and the number of controls per case | Reported pp 7–8 | Not applicable—no matching; active-comparator cohorts were weighted |
| Variables | 7 | Clearly define all outcomes, exposures, predictors, potential confounders, and effect modifiers. Give diagnostic criteria, if applicable | Reported pp 6–7, 33–46, 64 | Methods—Outcome/Covariates/Heterogeneity; Appendix definitions; Tables S1, S2, S5 |
| Data sources/measurement | 8* | For each variable of interest, give sources of data and details of methods of assessment (measurement). Describe comparability of assessment methods if there is more than one group | Reported pp 5–7, 32, 35–46 | Methods/Appendix—data sources, vocabularies, and concept IDs; some field/procedure details unavailable |
| Bias | 9 | Describe any efforts to address potential sources of bias | Reported pp 5, 7–8, 21–25, 39–40 | Methods—Design/Validation; Appendix—Validation; Discussion—Limitations |
| Study size | 10 | Explain how the study size was arrived at | Reported pp 8, 11 | Results opening; Fig 2; no a-priori size/power calculation |
| Quantitative variables | 11 | Explain how quantitative variables were handled in the analyses. If applicable, describe which groupings were chosen and why | Reported<br>pp 7, 39, 64 | Methods/Appendix—Heterogeneity; Table S5: cut points and comparison-specific tertiles |
| Statistical methods | 12 | (a) Describe all statistical methods, including those used to control for confounding | Reported<br>pp 7–8, 36–40 | Methods—Causal ML; Appendix—Detailed CML analysis and Validation |
|  |  | (b) Describe any methods used to examine subgroups and interactions | Reported<br>pp 7, 39, 64 | Methods/Appendix—Treatment effect heterogeneity; Table S5 |
|  |  | (c) Explain how missing data were addressed | Reported<br>pp 15, 19–20, 55 | Figs 4/6 legends; secondary analyses; primary imputation handling unavailable |
|  |  | (d) Cohort study—If applicable, explain how loss to follow-up was addressed<br>Case-control study—If applicable, explain how matching of cases and controls was addressed<br>Cross-sectional study—If applicable, describe analytical methods taking account of sampling strategy | Reported<br>pp 5, 32, 34 | Methods/Appendix—Study design: right-censoring used; operational loss-to-follow-up definition unavailable |
|  |  | (e) Describe any sensitivity analyses | Reported<br>pp 8, 20–21, 57, 62–71 | Methods—Validation; Results robustness; Appendix—Sensitivity analyses |
| Results |  |  |  |  |
| Participants | 13* | (a) Report numbers of individuals at each stage of study—eg numbers potentially eligible, examined for eligibility, confirmed eligible, included in the study, completing follow-up, and analysed | Reported<br>pp 8, 11 | Results opening; Fig 2 gives counts at each selection stage |
|  |  | (b) Give reasons for non-participation at each stage | Reported<br>pp 6, 11, 34–35 | Fig 2; Methods/Appendix—Eligibility give exclusion reasons |
|  |  | (c) Consider use of a flow diagram | Reported<br>p 11 | Fig 2—participant-flow diagram |
| Descriptive data | 14* | (a) Give characteristics of study participants (eg demographic, clinical, social) and information on exposures and potential confounders | Reported<br>pp 8–10, 47–49 | Results; Table 1; full covariates and balance in Table S3 |
|  |  | (b) Indicate number of participants with missing data for each variable of interest | Reported<br>pp 9–10, 47–49 | Tables 1/S3; complete per-variable missingness unavailable |
|  |  | (c) Cohort study—Summarise follow-up time (eg, average and total amount) | Reported<br>p 51 | Fig S2: risk/censoring to 120 months; median/total follow-up unavailable |
| Outcome data | 15* | Cohort study—Report numbers of outcome events or summary measures over time | Reported<br>p 51 | Appendix—Cumulative hazard; Fig S2 gives cumulative events over time |
|  |  | Case-control study—Report numbers in each exposure category, or summary measures of exposure |  |  |
|  |  | Cross-sectional study—Report numbers of outcome events or summary measures |  |  |
| Main results | 16 | (a) Give unadjusted estimates and, if applicable, confounder-adjusted estimates and their precision (eg, 95% confidence interval). Make clear which confounders were adjusted for and why they were included | Reported<br>pp 11–19 | Results; Figs 3–6: adjusted RMST effects and 95% CIs; unadjusted primary RMST unavailable |
|  |  | (b) Report category boundaries when continuous variables were categorized | Reported<br>p 64 | Appendix—Treatment effect heterogeneity; Table S5 |
|  |  | (c) If relevant, consider translating estimates of relative risk into absolute risk for a meaningful time period | Reported<br>pp 2–3 | Summary—Findings: 48-month RMST differences and approximate NNTs |
| Other analyses | 17 | Report other analyses done—eg analyses of subgroups and interactions, and sensitivity analyses | **Reported**<br>pp 11–21, 50–71 | Results; Figs 3–6; Appendix—secondary, subgroup, negative-control, sensitivity analyses |
| **Discussion** |  |  |  |  |
| Key results | 18 | Summarise key results with reference to study objectives | **Reported**<br>pp 21, 25 | Discussion—opening and concluding paragraphs |
| Limitations | 19 | Discuss limitations of the study, taking into account sources of potential bias or imprecision. Discuss both direction and magnitude of any potential bias | **Reported**<br>pp 23–25, 74–76 | Discussion; Supplementary Discussion—Limitations |
| Interpretation | 20 | Give a cautious overall interpretation of results considering objectives, limitations, multiplicity of analyses, results from similar studies, and other relevant evidence | **Reported**<br>pp 3, 21–25, 72–76; separate file | Summary—Interpretation; separate Research in context file; Discussion; Supplementary Discussion |
| Generalisability | 21 | Discuss the generalisability (external validity) of the study results | **Reported**<br>p 75 | Supplementary Discussion—Generalisability and transportability |
| **Other information** |  |  |  |  |
| Funding | 22 | Give the source of funding and the role of the funders for the present study and, if applicable, for the original study on which the present article is based | **Reported**<br>pp 3, 8, 31 | Summary—Funding; Methods—Role of the funding source; Acknowledgements |

\*Give information separately for cases and controls in case-control studies and, if applicable, for exposed and unexposed groups in cohort and cross-sectional studies.
**Note:** An Explanation and Elaboration article discusses each checklist item and gives methodological background and published examples of transparent reporting. The STROBE checklist is best used in conjunction with this article (freely available on the Web sites of PLoS Medicine at http://www.plosmedicine.org/, Annals of Internal Medicine at http://www.annals.org/, and Epidemiology at http://www.epidem.com/). Information on the STROBE Initiative is available at www.strobe-statement.org.

## The RECORD statement – checklist of items, extended from the STROBE statement, that should be reported in observational studies using routinely collected health data

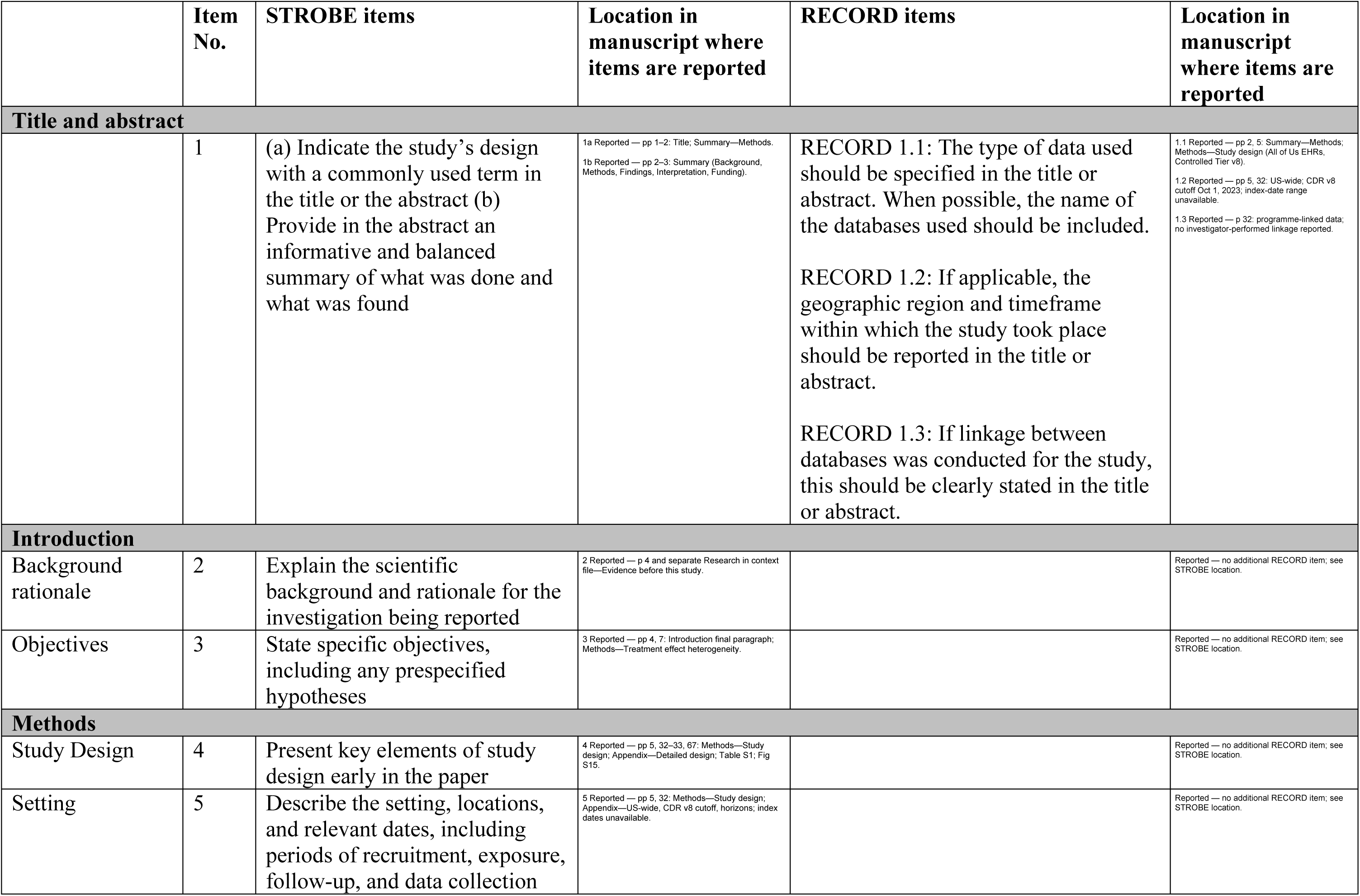
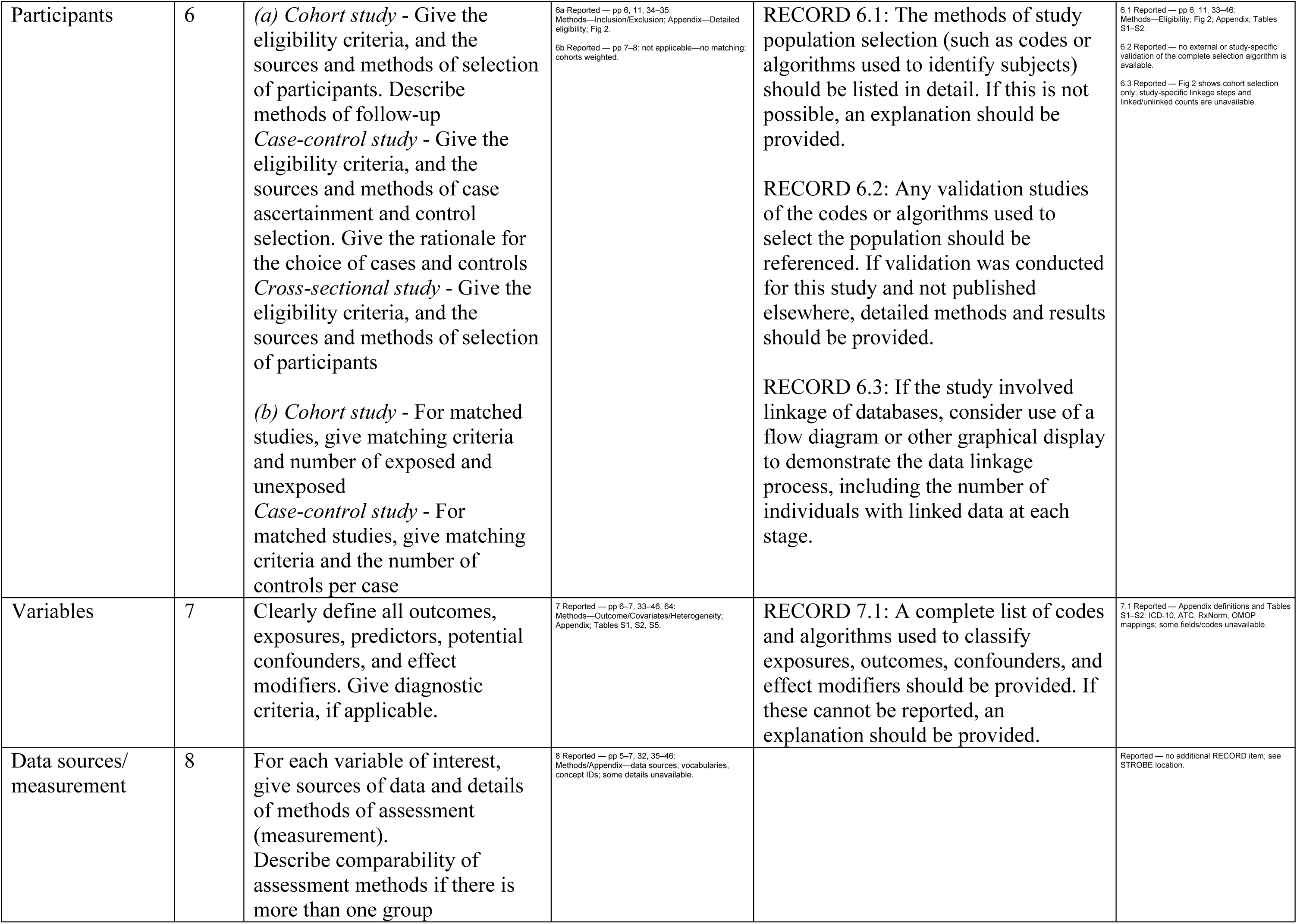
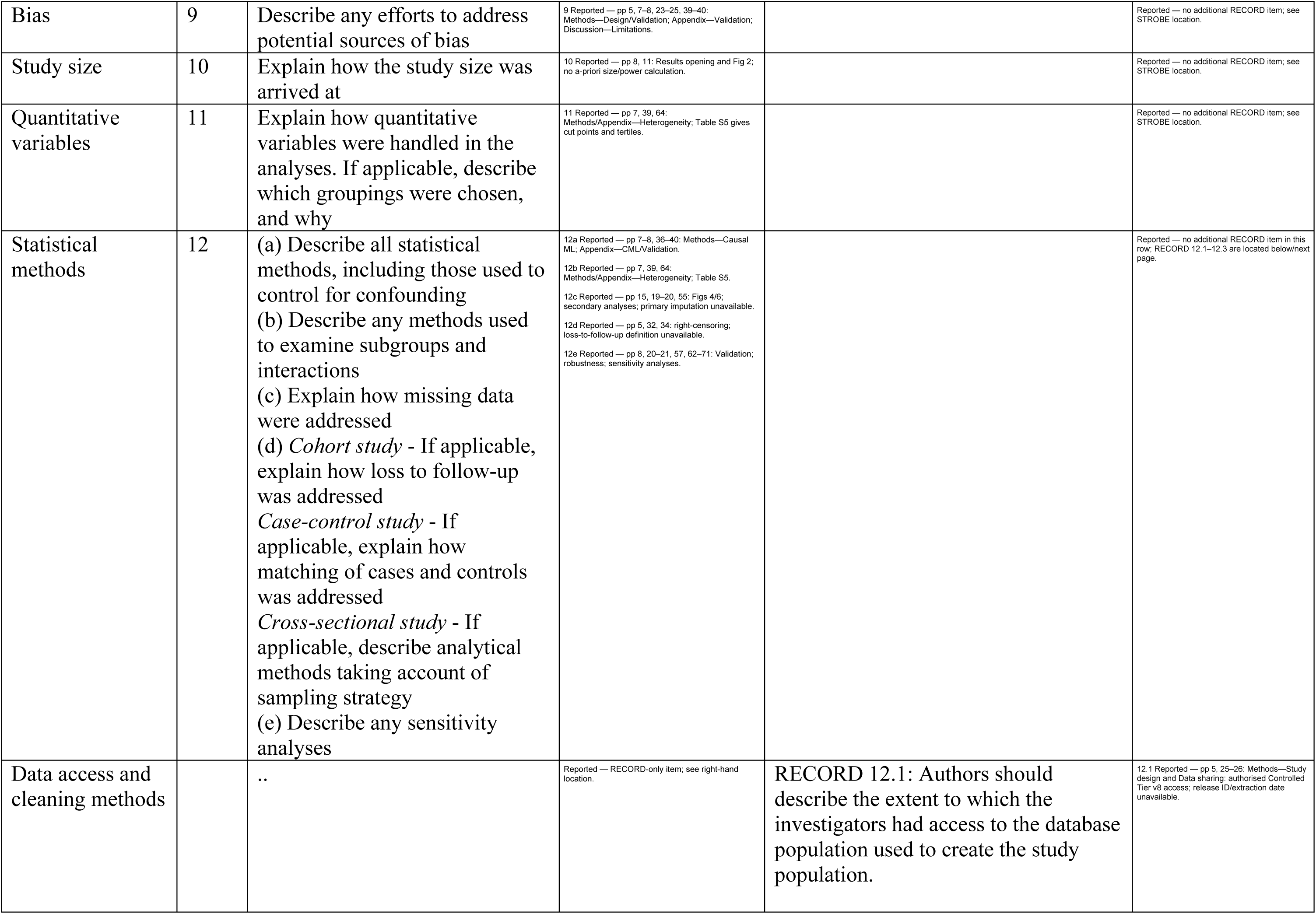
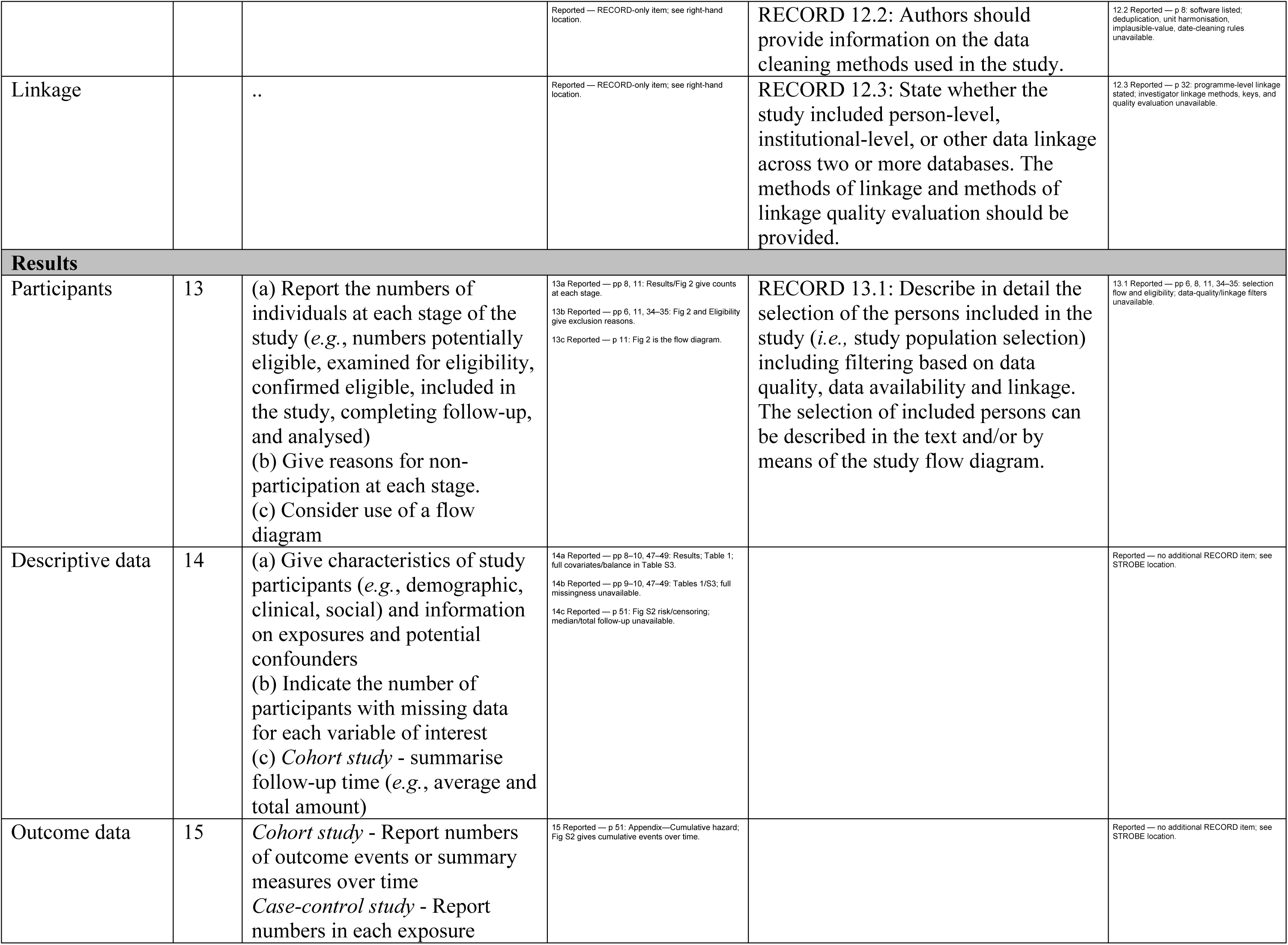
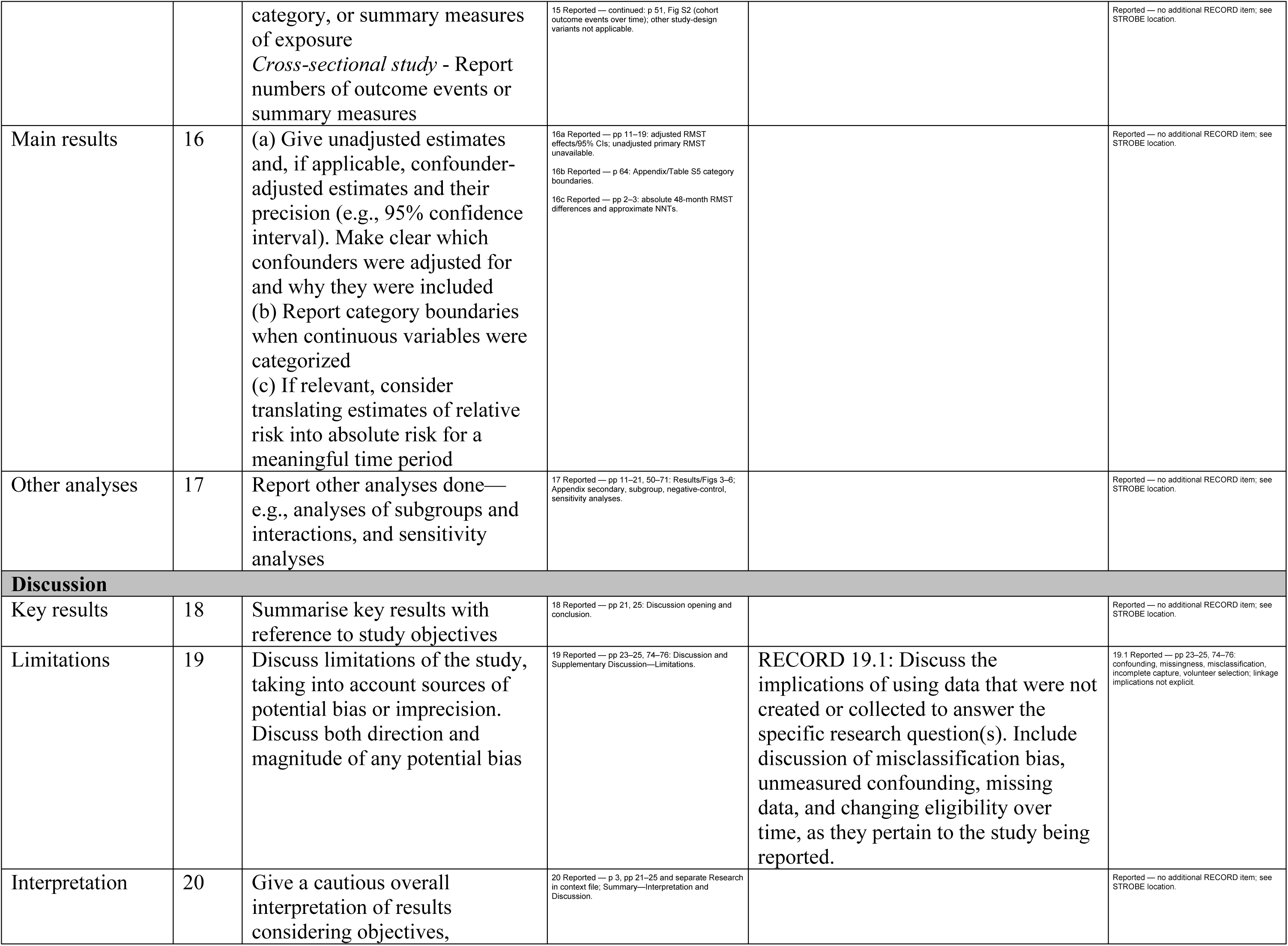
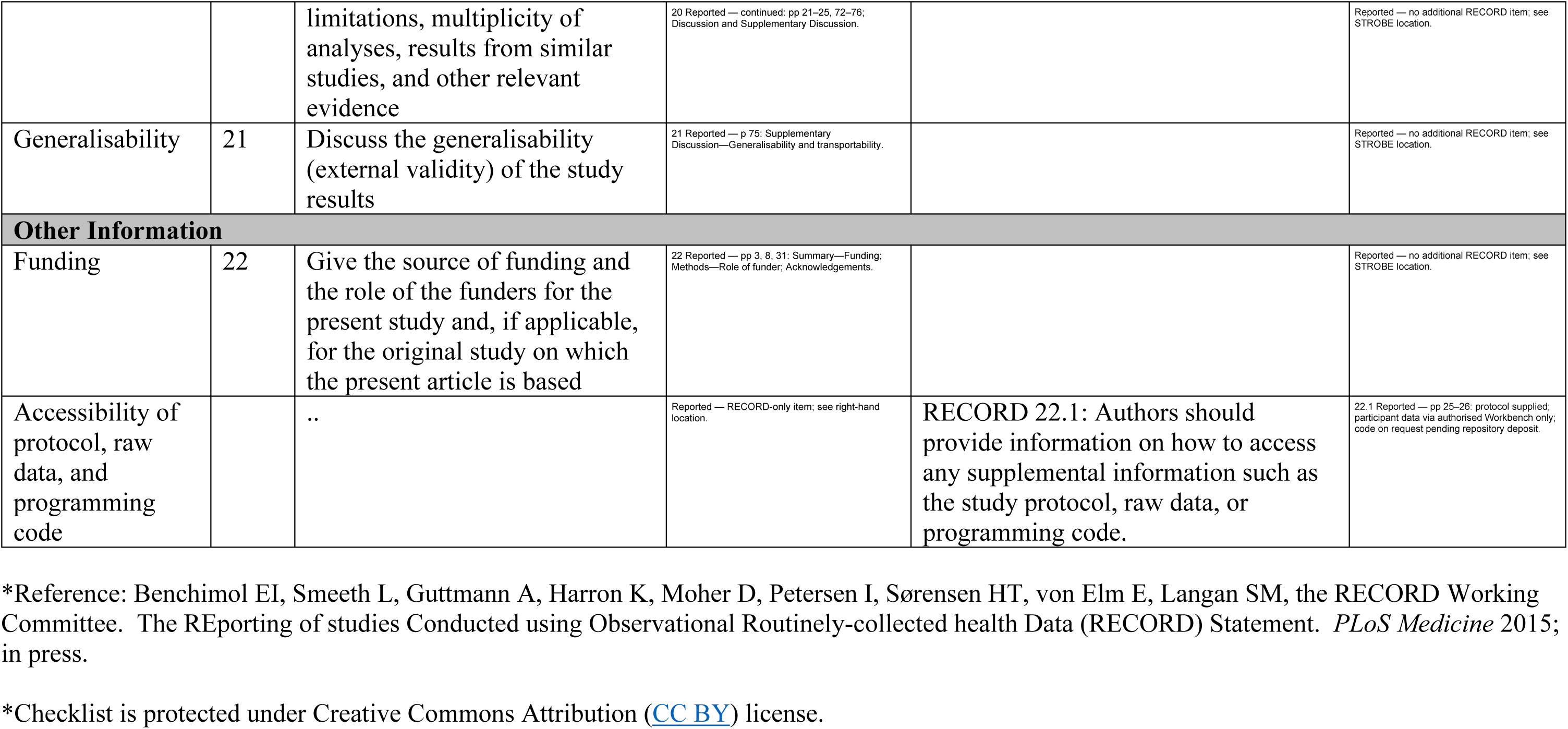

## References

[1] Livingston G, Huntley J, Sommerlad A, et al. Dementia Prevention, Intervention, and Care: 2020 Report of the Lancet Commission. Lancet 2020; 396: 413–46.

[2] GBD 2019 Dementia Forecasting Collaborators. Estimation of the Global Prevalence of Dementia in 2019 and Forecasted Prevalence in 2050: An Analysis for the Global Burden of Disease Study 2019. Lancet Public Health 2022; 7: e105–25.

[3] McKhann GM, Knopman DS, Chertkow H, et al. The Diagnosis of Dementia Due to Alzheimer’s Disease: Recommendations from the National Institute on Aging-Alzheimer’s Association Workgroups on Diagnostic Guidelines for Alzheimer’s Disease. Alzheimers Dement 2011; 7: 263–9.

[4] Hampel H, Vergallo A, Perry G, Lista S, Alzheimer Precision Medicine Initiative (APMI). The Alzheimer Precision Medicine Initiative. J Alzheimers Dis 2019; 68: 1–24.

[5] Yao H, Zhang A, Li D, et al. Comparative Effectiveness of GLP-1 Receptor Agonists on Glycaemic Control, Body Weight, and Lipid Profile for Type 2 Diabetes: Systematic Review and Network Meta-Analysis. BMJ 2024; 384: e076410.

[6] De Giorgi R, Ghenciulescu A, Yotter C, Taquet M, Koychev I. Glucagon-like Peptide-1 Receptor Agonists for Major Neurocognitive Disorders. J Neurol Neurosurg Psychiatry 2025; 96: 870–83.

[7] Sabbagh MN, Cummings JL, Ballard C, et al. Repurposing Glucagon-like Peptide-1 Receptor Agonists for the Treatment of Neurodegenerative Disorders. Nat Aging 2026; 6: 56–67.

[8] Drucker DJ. The Expanding Landscape of GLP-1 Medicines. Nat Med 2026; 32: 47–57.

[9] Zheng Z, Zong Y, Ma Y, et al. Glucagon-like Peptide-1 Receptor: Mechanisms and Advances in Therapy. Signal Transduct Target Ther 2024; 9: 234.

[10] Moiz A, Filion KB, Tsoukas MA, Yu OHY, Peters TM, Eisenberg MJ. The Expanding Role of GLP-1 Receptor Agonists: A Narrative Review of Current Evidence and Future Directions. EClinicalMedicine 2025; 86: 103363.

[11] Lin HT, Tsai YF, Liao PL, Wei JCC. Neurodegeneration and Stroke After Semaglutide and Tirzepatide in Patients With Diabetes and Obesity. JAMA Netw Open 2025; 8: e2521016.

[12] Hölscher C. Novel Dual GLP-1/GIP Receptor Agonists Show Neuroprotective Effects in Alzheimer’s and Parkinson’s Disease Models. Neuropharmacology 2018; 136: 251–9.

[13] Edison P, Femminella GD, Ritchie C, et al. Liraglutide in Mild to Moderate Alzheimer’s Disease: A Phase 2b Clinical Trial. Nat Med 2026; 32: 353–61.

[14] Cummings JL, Atri A, Sano M, et al. Efficacy and Safety of Oral Semaglutide 14 Mg (Flexible Dose) in Early-Stage Symptomatic Alzheimer’s Disease (Evoke and Evoke+): Two Phase 3, Randomised, Placebo-Controlled Trials. Lancet 2026; 407: 2167–79.

[15] Seminer A, Mulihano A, O’Brien C, et al. Cardioprotective Glucose-Lowering Agents and Dementia Risk: A Systematic Review and Meta-Analysis. JAMA Neurol 2025; 82: 450–60.

[16] Feuerriegel S, Frauen D, Melnychuk V, et al. Causal Machine Learning for Predicting Treatment Outcomes. Nat Med 2024; 30: 958–68.

[17] Desai RJ, Glynn RJ, Solomon SD, Claggett B, Wang SV, Vaduganathan M. Individualized Treatment Effect Prediction with Machine Learning—Salient Considerations. NEJM Evid 2024; 3: EVIDoa2300041.

[18] Ramirez AH, Sulieman L, Schlueter DJ, et al. The All of Us Research Program: Data Quality, Utility, and Diversity. Patterns (N Y) 2022; 3: 100570.

[19] Hernán MA, Robins JM. Using Big Data to Emulate a Target Trial When a Randomized Trial Is Not Available. Am J Epidemiol 2016; 183: 758–64.

[20] Tang B, Sjölander A, Wastesson JW, et al. Comparative Effectiveness of Glucagon-like Peptide-1 Agonists, Dipeptidyl Peptidase-4 Inhibitors, and Sulfonylureas on the Risk of Dementia in Older Individuals with Type 2 Diabetes in Sweden: An Emulated Trial Study. EClinicalMedicine 2024; 73: 102689.

[21] Cui Y, Kosorok MR, Sverdrup E, Wager S, Zhu R. Estimating Heterogeneous Treatment Effects with Right-Censored Data via Causal Survival Forests. J R Stat Soc Series B Stat Methodol 2023; 85: 179–211.

[22] Athey S, Tibshirani J, Wager S. Generalized Random Forests. Ann Stat 2019; 47: 1148–78.

[23] Chernozhukov V, Chetverikov D, Demirer M, et al. Double/Debiased Machine Learning for Treatment and Structural Parameters. Econom J 2018; 21: C1–C68.

[24] Wang G, Heagerty PJ, Dahabreh IJ. Using Effect Scores to Characterize Heterogeneity of Treatment Effects. JAMA 2024; 331: 1225–6.

[25] Seitz KP, Spicer AB, Casey JD, et al. Individualized Treatment Effects of Bougie versus Stylet for Tracheal Intubation in Critical Illness. Am J Respir Crit Care Med 2023; 207: 1602–11.

[26] Künzel SR, Sekhon JS, Bickel PJ, Yu B. Metalearners for Estimating Heterogeneous Treatment Effects Using Machine Learning. Proc Natl Acad Sci U S A 2019; 116: 4156–65.

[27] Lipsitch M, Tchetgen Tchetgen E, Cohen T. Negative Controls: A Tool for Detecting Confounding and Bias in Observational Studies. Epidemiology 2010; 21: 383–8.

[28] Chow AW, Benninger MS, Brook I, et al. IDSA Clinical Practice Guideline for Acute Bacterial Rhinosinusitis in Children and Adults. Clin Infect Dis 2012; 54: e72–e112.

[29] Nørgaard CH, Friedrich S, Hansen CT, et al. Treatment with Glucagon-like Peptide-1 Receptor Agonists and Incidence of Dementia: Data from Pooled Double-blind Randomized Controlled Trials and Nationwide Disease and Prescription Registers. Alzheimers Dement (N Y) 2022; 8: e12268.

[30] Sun M, Wang X, Lu Z, et al. Comparative Effectiveness of SGLT2 Inhibitors and GLP-1 Receptor Agonists in Preventing Alzheimer’s Disease, Vascular Dementia, and Other Dementia Types among Patients with Type 2 Diabetes. Diabetes Metab 2025; 51: 101623.

## Supplementary References

[1] Hernán MA, Robins JM. Using Big Data to Emulate a Target Trial When a Randomized Trial Is Not Available. Am J Epidemiol 2016; 183: 758–64.

[2] Ramirez AH, Sulieman L, Schlueter DJ, et al. The All of Us Research Program: Data Quality, Utility, and Diversity. Patterns (N Y) 2022; 3: 100570.

[3] Keogh RH, Gran JM, Seaman SR, Davies G, Vansteelandt S. Causal Inference in Survival Analysis Using Longitudinal Observational Data: Sequential Trials and Marginal Structural Models. Stat Med 2023; 42: 2191–225.

[4] Tang B, Sjölander A, Wastesson JW, et al. Comparative Effectiveness of Glucagon-like Peptide-1 Agonists, Dipeptidyl Peptidase-4 Inhibitors, and Sulfonylureas on the Risk of Dementia in Older Individuals with Type 2 Diabetes in Sweden: An Emulated Trial Study. EClinicalMedicine 2024; 73: 102689.

[5] Lin L, Sperrin M, Jenkins DA, Martin GP, Peek N. A Scoping Review of Causal Methods Enabling Predictions under Hypothetical Interventions. Diagn Progn Res 2021; 5: 3.

[6] Feuerriegel S, Frauen D, Melnychuk V, et al. Causal Machine Learning for Predicting Treatment Outcomes. Nat Med 2024; 30: 958–68.

[7] Dang LE, Gruber S, Lee H, et al. A Causal Roadmap for Generating High-Quality RealWorld Evidence. J Clin Transl Sci 2023; 7: e212.

[8] Cui Y, Kosorok MR, Sverdrup E, Wager S, Zhu R. Estimating Heterogeneous Treatment Effects with Right-Censored Data via Causal Survival Forests. J R Stat Soc Series B Stat Methodol 2023; 85: 179–211.

[9] Breiman L. Random Forests. Mach Learn 2001; 45: 5–32.

[10] Athey S, Tibshirani J, Wager S. Generalized Random Forests. Ann Stat 2019; 47: 1148–78.

[11] Austin PC. An Introduction to Propensity Score Methods for Reducing the Effects of Confounding in Observational Studies. Multivariate Behav Res 2011; 46: 399–424.

[12] Chernozhukov V, Chetverikov D, Demirer M, et al. Double/Debiased Machine Learning for Treatment and Structural Parameters. Econom J 2018; 21: C1–C68.

[13] Wang G, Heagerty PJ, Dahabreh IJ. Using Effect Scores to Characterize Heterogeneity of Treatment Effects. JAMA 2024; 331: 1225–6.

[14] Bang H, Robins JM. Doubly Robust Estimation in Missing Data and Causal Inference Models. Biometrics 2005; 61: 962–73.

[15] Seitz KP, Spicer AB, Casey JD, et al. Individualized Treatment Effects of Bougie versus Stylet for Tracheal Intubation in Critical Illness. Am J Respir Crit Care Med 2023; 207: 1602–11.

[16] Lipsitch M, Tchetgen Tchetgen E, Cohen T. Negative Controls: A Tool for Detecting Confounding and Bias in Observational Studies. Epidemiology 2010; 21: 383–8.

[17] Chow AW, Benninger MS, Brook I, et al. IDSA Clinical Practice Guideline for Acute Bacterial Rhinosinusitis in Children and Adults. Clin Infect Dis 2012; 54: e72–e112.

[18] Künzel SR, Sekhon JS, Bickel PJ, Yu B. Metalearners for Estimating Heterogeneous Treatment Effects Using Machine Learning. Proc Natl Acad Sci U S A 2019; 116: 4156–65.

[19] Tchetgen Tchetgen E. The Control Outcome Calibration Approach for Causal Inference With Unobserved Confounding. Am J Epidemiol 2014; 179: 633–40.

[20] Lundberg SM, Lee SI. A Unified Approach to Interpreting Model Predictions. In: Proceedings of the 31st International Conference on Neural Information Processing Systems. NIPS’17. Red Hook, NY, USA: Curran Associates Inc.; 2017. p. 4768–77.

[21] Edison P, Femminella GD, Ritchie C, et al. Liraglutide in Mild to Moderate Alzheimer’s Disease: A Phase 2b Clinical Trial. Nat Med 2026; 32: 353–61.

[22] Cummings JL, Atri A, Sano M, et al. Efficacy and Safety of Oral Semaglutide 14 Mg (Flexible Dose) in Early-Stage Symptomatic Alzheimer’s Disease (Evoke and Evoke+): Two Phase 3, Randomised, Placebo-Controlled Trials. Lancet 2026; 407: 2167–79.

[23] Cheng HW, Yang SF, Liao PL, Lee CH, Jong GP. Impact of Glucagon-Like Peptide-1 Receptor Agonists on the Dementia Incidence in Patients With Type 2 Diabetes Mellitus: A Population-Based Longitudinal Cohort Study. Diabetes Metab Res Rev 2025; 41: e70058.

[24] Davies MJ, Aroda VR, Collins BS, et al. Management of Hyperglycemia in Type 2 Diabetes, 2022. A Consensus Report by the American Diabetes Association (ADA) and the European Association for the Study of Diabetes (EASD). Diabetes Care 2022; 45: 2753–86.

[25] Nauck MA, Quast DR, Wefers J, Meier JJ. GLP-1 Receptor Agonists in the Treatment of Type 2 Diabetes State-of-the-Art. Mol Metab 2021; 46: 101102.

[26] Carreiras MC, Mendes E, Perry MJ, Francisco AP, Marco-Contelles J. The Multifactorial Nature of Alzheimer’s Disease for Developing Potential Therapeutics. Curr Top Med Chem 2013; 13: 1745–70.

[27] Zheng Z, Zong Y, Ma Y, et al. Glucagon-like Peptide-1 Receptor: Mechanisms and Advances in Therapy. Signal Transduct Target Ther 2024; 9: 234.

[28] Moiz A, Filion KB, Tsoukas MA, Yu OHY, Peters TM, Eisenberg MJ. The Expanding Role of GLP-1 Receptor Agonists: A Narrative Review of Current Evidence and Future Directions. EClinicalMedicine 2025; 86: 103363.

[29] Drucker DJ. The Expanding Landscape of GLP-1 Medicines. Nat Med 2026; 32: 47–57.

[30] Wilding JPH, Batterham RL, Calanna S, et al. Once-Weekly Semaglutide in Adults with Overweight or Obesity. N Engl J Med 2021; 384: 989–1002.

[31] Yao H, Zhang A, Li D, et al. Comparative Effectiveness of GLP-1 Receptor Agonists on Glycaemic Control, Body Weight, and Lipid Profile for Type 2 Diabetes: Systematic Review and Network Meta-Analysis. BMJ 2024; 384: e076410.

